# A regression-based approach to modelling recurrent outbreaks: a case study of dengue fever in Nepal (2006–2022)

**DOI:** 10.1101/2025.05.21.25327909

**Authors:** Jessie Jane Khaki, Bipin Kumar Acharya, Kouichi Morita, Basu Dev Pandey, Emanuele Giorgi

**Affiliations:** Centre for Health Informatics, Computing and Statistics, Lancaster University, Lancaster, United Kingdom; Malawi-Liverpool-Wellcome Programme (MLW), Blantyre, Malawi; School of Global and Public Health, Kamuzu University of Health Sciences, Blantyre, Malawi; Planetary Health Research Center, Kathmandu, Nepal; Nepal Open University, Lalitpur, Nepal; Nepal Geographical Society, Kathmandu, Nepal; DEJIMA Infectious Disease Research Alliances, Nagasaki University, Japan; Department of Virology, Institute of Tropical Medicine, Nagasaki University, Japan; Everest International Clinic and Research Center, Kathmandu, Nepal

**Keywords:** Dengue, Epidemic, Infectious disease, Outbreak, Statistical modelling, Nepal

## Abstract

Understanding the temporal and spatial dynamics of dengue fever outbreaks is essential for effective public health planning. In this study, we develop a novel, data-driven approach to characterise dengue outbreaks in Nepal from 2006 to 2022 by extending the standard Negative Binomial regression model to incorporate multiple outbreak intensity functions (OIFs). Each OIF is defined by three interpretable parameters: the peak timing, the scale regulating the duration of the outbreak, and a magnitude parameter that quantifies the contribution of the OIF to the overall epidemic. Unlike mechanistic transmission models, this framework makes minimal assumptions and offers greater flexibility when working with aggregated surveillance data. The application of this modelling approach to annual dengue case counts from all 77 districts of Nepal, revealed substantial heterogeneity in outbreak timing and intensity. Most districts experienced three major OIFs, with the third, typically in 2022, accounting for the largest share of the disease burden. The modelling approach is generalisable and can be applied to routinely reported data for other infectious diseases. We also outline how the framework could be extended for outbreak detection if higher-resolution temporal data were available.

## 1. Introduction

Dengue is the most rapidly growing mosquito-borne disease in the world and can be caused by four viruses (DENV): DENV-1, DENV-2, DENV-3, and DENV-4 (Yang et al., 2021; Colón-González et al., 2021). The number of dengue infections has been steadily increasing in the last 30 years (Zeng et al., 2021a). There are estimated to be more than 100 million dengue cases yearly, approximately 20,000 of which result in death (Zeng et al., 2021b; Yang et al., 2021). Recent reports show that the death rate from dengue has increased by almost 70%, from 0.31 per 100,000 population in 1990 to 0.53 per 100,000 population in 2017 (Zeng et al., 2021a). Disability adjusted life years (DALYs) due to dengue have also increased more than a hundred-fold (Zeng et al., 2021a; Gathsaurie et al., 2023). Furthermore, recent studies estimate that the potential number of people at risk of dengue could increase by an additional 4.7 billion individuals by 2070 due to climate change (Colón-González et al., 2021; Messina et al., 2019). Due to its high burden level, the World Health Organization (WHO) has declared dengue one of the top ten threats to global health and upgraded it to highest threat level 3 (, WHO). Dengue is endemic in more than 100 countries, with the highest cases occurring in tropical and subtropical regions such as the Caribbeans, South Asia, Southeast Asia, and Latin America (Yang et al., 2021; Gathsaurie et al., 2023). Approximately 70% of the burden of dengue is estimated to be in Asia (Bhatt et al., 2013).

Urbanization, climate change, and co-circulation of multiple DENV serotypes are hypothesized to be part of the variables driving the dengue epidemic in Nepal and elsewhere (Pandey and Costello, 2019; Gathsaurie et al., 2023). Previous studies have shown that the density of Aedes aegypti mosquitoes, which transmit dengue viruses (DENV 1-4), is highly influenced by environmental and climatic variables such as rainfall, temperature, and precipitation (Kong et al., 2019; Pandey and Costello, 2019; Phanitchat et al., 2019). An increase in climatic variables such as temperature and precipitation is associated with an increase in the incidence of dengue (Sun et al., 2017; Phanitchat et al., 2019; Abdulsalam et al., 2022; Dhimal et al., 2014). However, other studies in Asian countries such as Thailand found that the relationship between dengue and climatic variables such as precipitation and rainfall is complex (Campbell et al., 2013). For instance, although precipitation can create a breeding ground for mosquitoes, excessive rainfall can wash away their breeding sites and thus reduce dengue incidence (Phanitchat et al., 2019; Li et al., 1985; Rozilawati et al., 2007). In addition to varying by time, the incidence of dengue has been shown to vary geographically (Kong et al., 2019; Sun et al., 2017; do Carmo et al., 2020). Furthermore, studies in other countries such as China have shown notable variations in the peaks and intensity of dengue outbreaks (Chen et al., 2022).

In Nepal, the first case of dengue was recorded in 2004 (Acharya et al., 2016; Pandey et al., 2004). Since then, the number of dengue cases in Nepal has increased rapidly from around 32 cases in 9 districts in 2006 to more than 50,000 cases in 2022, with all four DENV serotypes co-circulating in the country (Pandey and Costello, 2019; Rijal et al., 2021; Pandey et al., 2013; Dumre et al., 2017; Prajapati et al., 2020; Ngwe Tun et al., 2021). Although dengue has become endemic in Nepal, cyclical outbreaks of dengue cases are observed every two to three years (Rijal et al., 2021; Ngwe Tun et al., 2021; Acharya et al., 2020). Nepal’s first national dengue prevention, control, and management guidelines were developed in 2008 and revised in 2011 and 2019 (, WHO). Despite the successful implementation of dengue control measures in Nepal, dengue remains a public health concern.

Describing the dynamics of disease outbreaks is essential for investigating the underlying factors that drive them, enhancing the precision of outbreak predictions, and devising effective strategies to manage and mitigate the outbreaks (Viboud et al., 2016). Two key characteristics of disease outbreaks are the peak in the number of infected cases and the duration of the outbreak, which are especially important for formulating timely response measures, allocating resources efficiently, and implementing appropriate control strategies. In this paper, we defined the “ duration of an outbreak” as the length of time from the onset of the outbreak to the point when the number of new cases returns to baseline or significantly decreases to levels that are not of public health concern. In other words, the duration of the outbreak is the period that includes the rise, peak and decline of the outbreak, encompassing all phases of the epidemic curve. The statistical problem we address in this paper is how to accurately model these aspects using a data-driven approach, particularly when data availability is limited due to temporal and spatial aggregation.

Existing modelling methods used for describing these aspects of disease outbreaks have mainly focused on the use of compartmental models such as the Susceptible-Exposed-Infectious-Removed (SEIR) and Susceptible-Infectious-Quarantined-Recovered (SIQR) (Tovissodé et al., 2020; Cooper et al., 2015; Chen et al., 2022). However, these models have inherent limitations, as they do not allow the inclusion of spatial risk factors unless the fitting is done by stratifying for the covariates of interest (Franco et al., 2022), and cannot easily accommodate the occurrence of multiple outbreaks. Other studies have employed epidemic-endemic models such as the modified Poisson and Negative Binomial to describe outbreaks of diseases such as COVID-19 and measles Semakula et al. (2023); Parpia et al. (2020). Prior research on characterizing dengue outbreaks has focused on the rate at which individuals susceptible to dengue become infected, a parameter also referred to as the force of infection (Man et al., 2023; Imai et al., 2015). Whilst this parameter affects the peak and duration of an outbreak, these two characteristics are not explicitly modelled and explained in epidemic-endemic models. Moreover, existing epidemic models do not allow for multiple outbreaks to be estimated in a single model.

In this study, we use a data-driven approach to extend the standard class of Negative Binomial regression models to estimate the peaks and scale parameters of dengue fever outbreak intensity functions (OIFs) at the district level in Nepal using yearly reported cases. Our proposed modelling framework allows for the estimation of multiple OIFs through the specification of an outbreak function, for which we consider three different specifications. In our modelling framework, we interpret the outbreak function as the unexpected increase in reported cases that cannot be explained by the covariates used in the regression. The Negative Binomial models are validated using the Chi-square goodness of fit test (Roback and Legler, 2021).

The structure of the paper is as follows. In Section 2, we describe the data and outline the modelling framework for characterising the dengue outbreak in Nepal. In Section 3, we illustrate the results of the descriptive analysis. We also illustrate the timing of the peak and scale parameters for at least three OIFs within each district. The discussion and conclusions of the study are presented in Section 5.

## 2. Methods and materials

### 2.1. Study site and Data collection

Nepal, a country of approximately 147,200 km^2^ (Acharya et al., 2018), borders India and China and is located at a latitude of 28.39° and a longitude of 84.12°. The elevation in Nepal ranges from 60 meters (*m*) to 8,848 m above sea level (Acharya et al., 2018).

Topographically, Nepal is divided into 11 main ecological zones, with the most prominent being the Terai region (67 m to 300 m above sea level), the Siwalik Hills (700 m to 1,500 m), the Mahabharat region (1,500 m to 2,700 m), and the Himalayan zone (above 4,000 m) (Bhuju et al., 2007). The climate in Nepal, therefore, varies significantly due to the wide variations in altitude (Acharya et al., 2018; Nayava, 1980; Bhuju et al., 2007). Approximately 80% of Nepal’s precipitation occurs from June to September in the summer monsoon season. The average annual rainfall in Nepal also varies widely and ranges from 295 millimetres (mm) to 3,345 mm, depending on the ecological zone (Bhuju et al., 2007). Furthermore, the average temperature decreases by 6° C for every 1,000 meters increase in altitude (Bhuju et al., 2007).

Nepal is administratively divided into 7 provinces, 77 districts, and 753 municipalities. We retrieved district-level annual dengue fever case data reported between 2006 and 2022 from the Epidemiology and Diseases Control Division (EDCD), Department of Health services, Ministry of Health and Population, Nepal responsible for the preparedness and response of the outbreak in Nepal (https://www.edcd.gov.np/section/dengue-control-program). The EDCD is also responsible for the prevention, surveillance, and control of communicable and non-communicable diseases in Nepal. Details on how data are collated from the facility level (such as district hospitals and health centers) to a central location at the EDCD have been published elsewhere (Rijal et al., 2021; Organization et al., 2019). Briefly, the official EDCD reports publish the district and year of every dengue case confirmed through laboratory tests. The annual number of dengue cases per district was defined as the sum of confirmed cases within a particular district. Figure 1 shows the location of the study.

**Figure 1:**
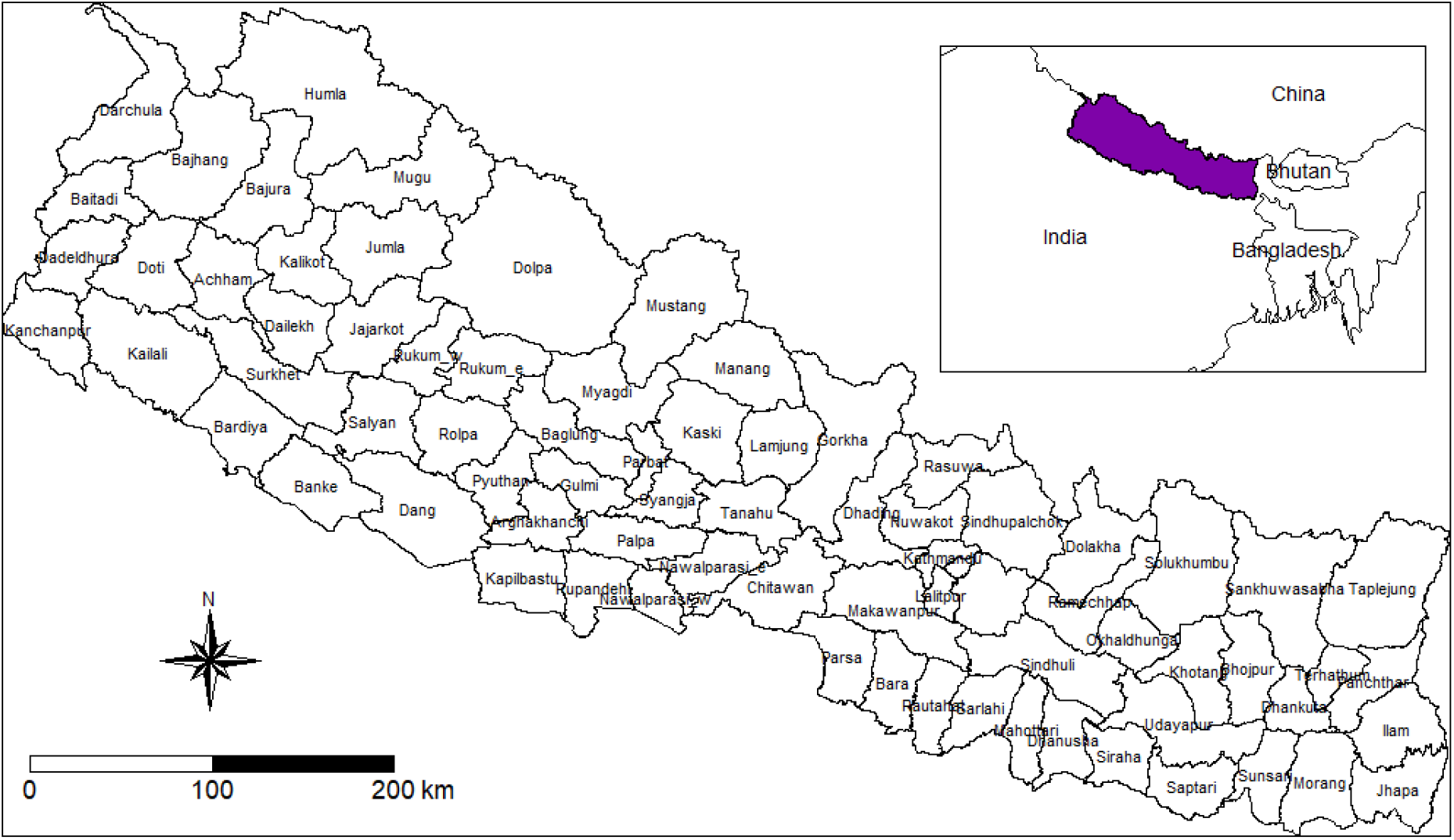
Map of Nepal showing the location of Nepal and the boundaries (black lines) of the 77 districts.

### 2.2. Spatio-temporal covariates, covariate processing and population data

We extracted a list of spatio-temporal environmental variables for which a significant association with the risk of dengue fever has been previously reported (Kong et al., 2019; Pandey and Costello, 2019; Sun et al., 2017; Phanitchat et al., 2019; Acharya et al., 2025). Table 1 outlines the list of environmental variables, their sources, and their temporal resolutions.

**Table 1:** List of environmental covariates and their sources.

| Covariate | Temporal Resolution | Source of Data |
| --- | --- | --- |
| Minimum, mean & maximum Precipitation | 2006-2022 | TerraClimate (Abatzoglou et al., 2018) |
| Minimum & maximum temperature | 2006-2022 | TerraClimate (Abatzoglou et al., 2018) |
| Potential evapotranspiration (PET) | 2006-2022 | TerraClimate (Abatzoglou et al., 2018) |
| Aridity index | 2006-2022 | Mean precipitation ÷ PET |

Following previous modelling studies, we explored the relationships between the covariates and the incidence of dengue fever using scatter plots to assess the strength of association (Giorgi et al., 2021; Utazi et al., 2020; Wariri et al., 2023). We combined the covariates using Principal Component Analysis (PCA) to develop an index of environmental exposure. The PCA was carried out to reduce the number of covariates in our final model to avoid overfitting due to the small sample size per district. The supplementary information presents the exploratory analysis and the PCA results. The variables were standardized prior to carrying out the PCA. The first component, which accounts for 48% of the variation in the PCA, was selected as our environmental exposure index (Figure 4).

We also obtained the yearly population counts per district from 2006 to 2020 from the WorldPop website (Tatem, 2017). The 2021 and 2022 population counts were derived by scaling the 2020 population using the Nepal population growth rate (0.92 per cent) reported in the 2021 Nepal National Population and Housing Census (Nepal National Statistics Office, 2021).

### 2.3. Statistical modelling

Let *Y*_*t*_ denote the yearly dengue case count at the district level in year *t* (t = 1 (2006), 2,3, …,17 (2022)). To account for the overdispersion, we then assume that *Y*_*t*_ follows a Negative Binomial (NB) distribution, i.e. *Y*_*t*_ *∼* NB(*λ*_*t*_, *α*), where *λ*_*t*_ denotes the annual dengue incidence, and *α* is the dispersion parameter. Let *θ* denote the vector of unknown model parameters; the likelihood function is then defined as (Zwilling, 2013):

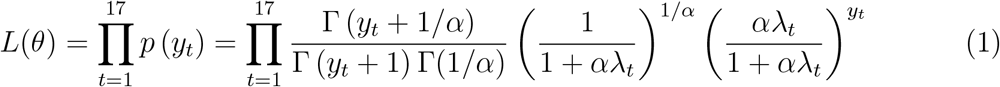

where *λ*_*t*_ is the mean number of dengue cases at time *t* and was defined as follows in Table 2. To characterize the outbreaks in Nepal, and assuming that there were a total of *K* outbreak intensity functions (OIFs) within each district, we extend the Negative Binomial regression model (equation 1) as follows. We initially assumed *K* = 3 main OIFs per district, based on expert recommendations and preliminary evaluation of annual incidence trends. Based on the results of the model validation (see following sections for details), we then incrementally increased this number until a satisfactory model fit was achieved.

**Table 2:**
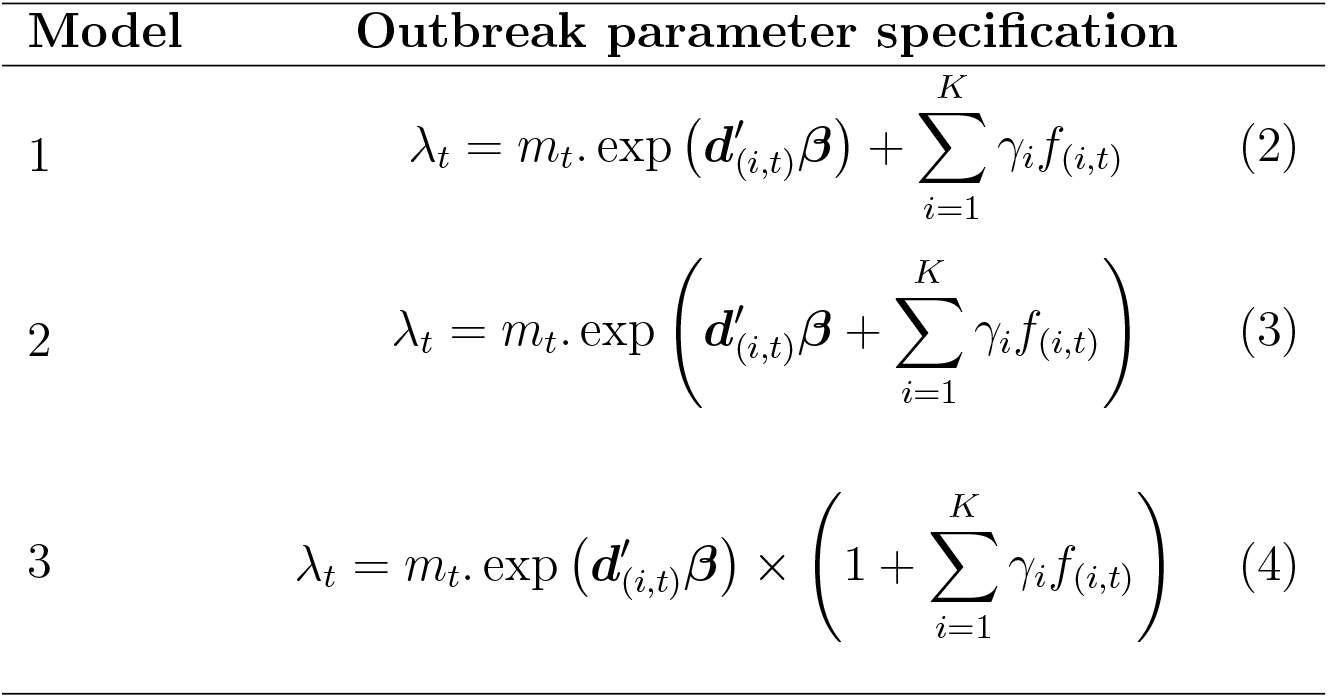
Specification of parameters used in characterizing the dengue outbreak in Nepal.

In equations 2, 3, and 4, the population count (*m*_*t*_) is used as an offset in the model, ***d***_(*i*,*t*)_ = (1, *d*_(*i*,*t*)_) is the vector containing the intercept and the environmental exposure index, and ***β*** = (*β*_0_, *β*_1_)^*′*^ is the coefficient vector associated with the intercept and environmental exposure index covariate. Additionally, ***γ*** = (*γ*_1_, *γ*_2_, *γ*_3_)^*′*^ is a coefficient vector for the *K* OIFs, and *f*_*t*_ is a function used to quantify the *intensity* of the outbreak in the district being considered for the analysis. Henceforth, we refer to *f*_*t*_ as the outbreak intensity function (OIF). In this setting, we used the squared exponential function (equation 5) to define each OIF for each district at time *t*:

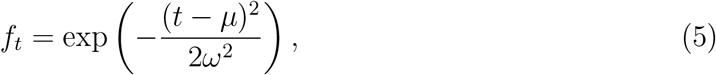

where *µ* and *ω* denote the peak and the scale parameter of the OIF.

In the three models considered in Table 2, the *γ* parameters reflect the contribution of the corresponding OIF to the mean dengue incidence, *λ*_*t*_. In Model 1 (equation 2), the effect of *γ*_*i*_ *f*_(*i*,*t*)_ is additive, which means that the OIF quantifies the excess number of cases that are not due to covariate effects. In Models 2 (equation 3) and 3 (equation 4), the effect is multiplicative. In Model 2, *γ*_*i*_*f*_(*i*,*t*)_ is incorporated into the linear predictor and its effect can be interpreted in the same way as the other covariates, i.e. by using exp*{γ*_*i*_*}* to quantify the multiplicative increase in the number of cases due to the OIF. In Model 3 (equation 4), the term 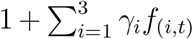 scales the overall mean dengue incidence. Here, *γ*_*i*_*f*_(*i*,*t*)_ acts as a proportion of the baseline incidence *m*_*t*_ exp(*d*^*′*^*β*), adjusting the number of cases relative to the baseline level, where the higher values of *γ*_*i*_*f*_(*i*,*t*)_ represent proportionally larger OIFs.

We fitted the Negative Binomial outbreak model to each of the districts separately due to the variability in the incidence of dengue fever within each district. Confidence intervals for parameters {*θ* = (*β*_1_, *β*_2_, ***γ, µ, ω***)} estimated from the Negative Binomial models were constructed using a bootstrap procedure following the steps below (Efron, 2000):

- Construct the first bootstrap sample by selecting a random sample of 17 (N) observations with replacement from our original sample.
- Fit the model to the first bootstrap sample and store the model coefficients.
- Repeat steps 1 and 2 10,000 times, drawing new random samples of size N with replacement each time to create subsequent bootstrap samples.
- Compute the confidence intervals using the Bias-Corrected and Accelerated (BCa) method, which utilizes the distribution of estimates obtained from the bootstrap samples as the sampling distribution (Efron and Tibshirani, 1994; Efron, 1987).

We assessed our models’ goodness of fit (gof) using the Chi-square gof test (Roback and Legler, 2021). The test, which has the null hypothesis that there is no significant difference between the observed and the expected dengue counts, is given as:

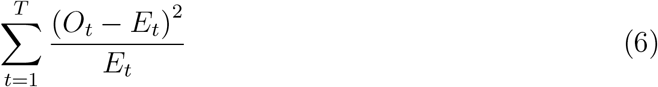

where *O*_*t*_ are the cases of dengue fever observed at time *t* and *E*_*t*_ are the predicted dengue counts from the model.

## 3. Results

### 3.1. Descriptive analysis

A total of 78,819 dengue cases were reported over the 17 years in all 77 districts. Figure 2 shows the total number of dengue cases in each district over 17 years (2006 to 2022). As can be seen, the highest total number of cases was observed in the central areas of Nepal. In particular, the districts with the observed dengue cases of at least 2,000 over the study period were Kathmandu (15,940), Lalitpur (10,220), Chitawan (8,845), Makawanpur (8,237), Bhaktapur (6,515), Rupandehi (3,695), Kaski (3,654), Dang (2,585), and Jhapa (2,008).

**Figure 2:**
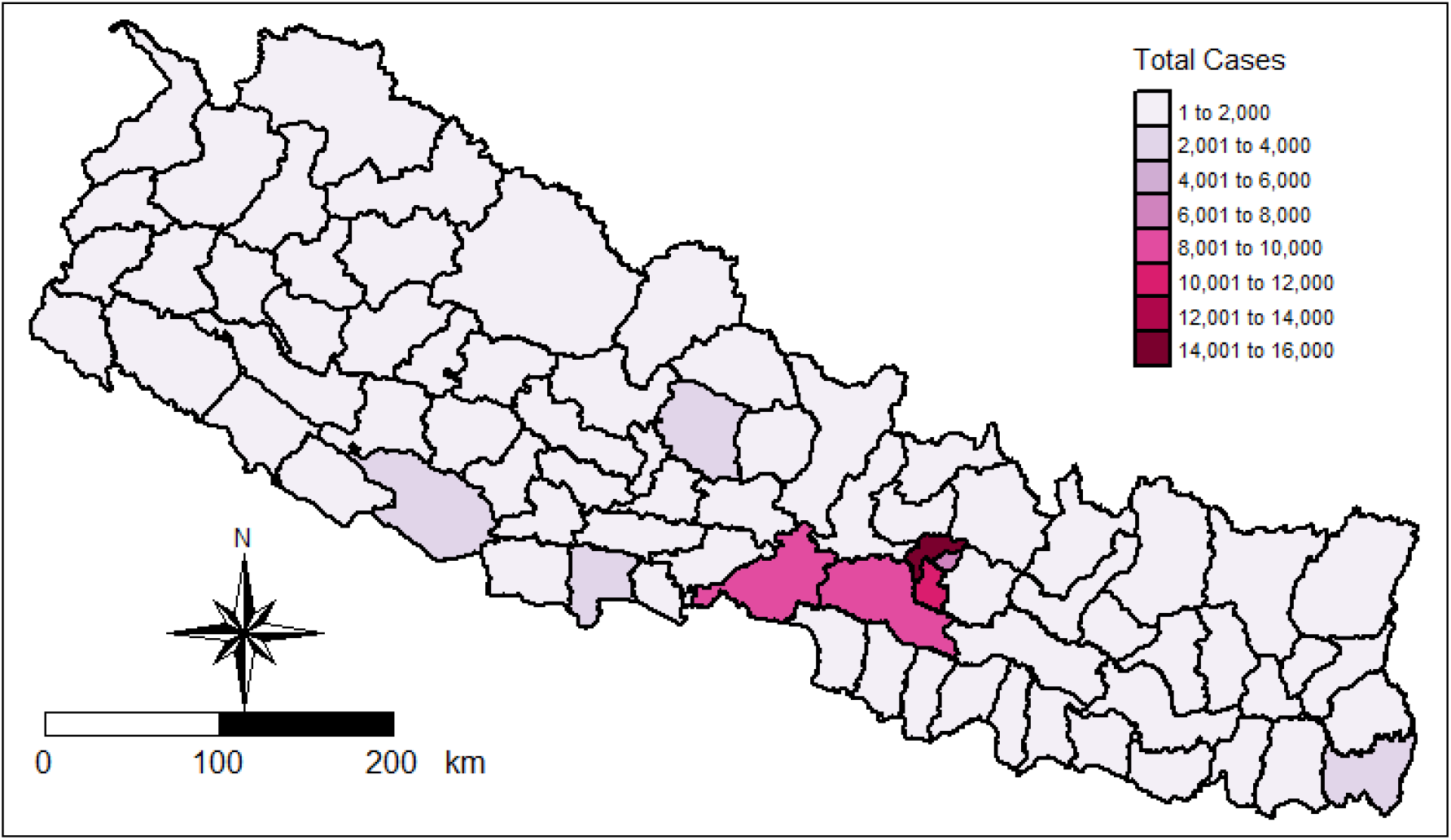
A map illustrating the total number of cases observed in each district over the study period.

In general, there was a rise in dengue fever incidence per 100,000 population in each district over time, as illustrated in Figure 3. A discernible pattern emerged, with notable peaks in 2010, 2013, 2016, 2019, and 2022. Notably, the year 2022 exhibited the highest incidence in most districts. Nevertheless, Figure 3 also highlights that the districts did not experience peaks in the same years. Our modelling framework accounts for the variations in peak occurrences by explicitly incorporating outbreak parameters such as peak and scale parameter, in each district.

**Figure 3:**
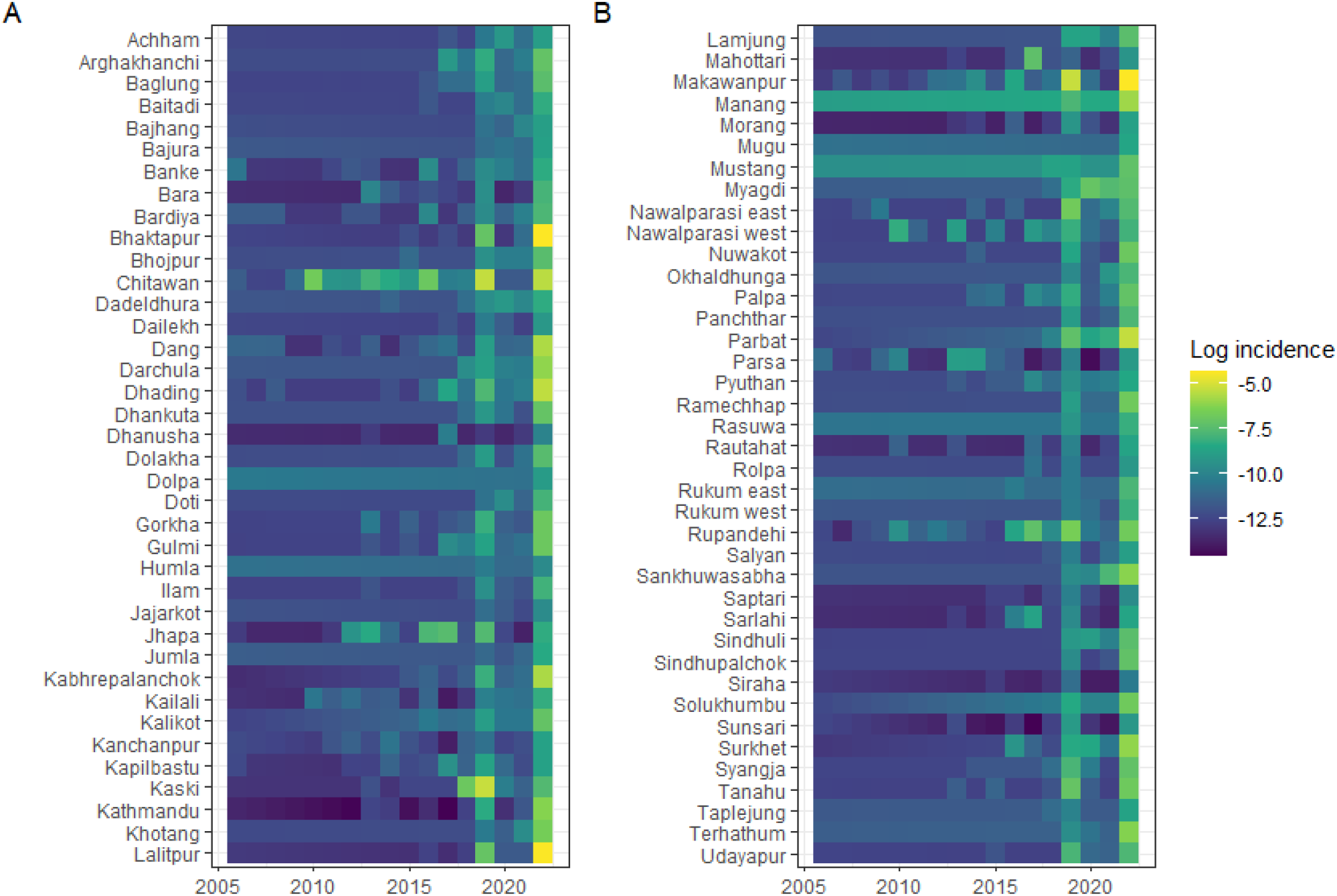
Heat maps illustrating the observed log-transformed cases of dengue fever per 100,000 population (log((count of dengue cases + 1) / population)) for each district from 2006 to 2022. The transformation adds 1 to the case counts before taking the logarithm to handle zero values.

### 3.2. Principal Components Analysis (PCA) results

An index of environmental exposure was derived from the above covariates (Table 1) using Principal Component Analysis (PCA). Figure 4 shows the variation percentage explained by the PCA components. Our environmental exposure score was based on the first principal component, which explained 48% of the variation in our data, supporting its use as a summary exposure score (Howe et al., 2012; Hjelm et al., 2017).

**Figure 4:**
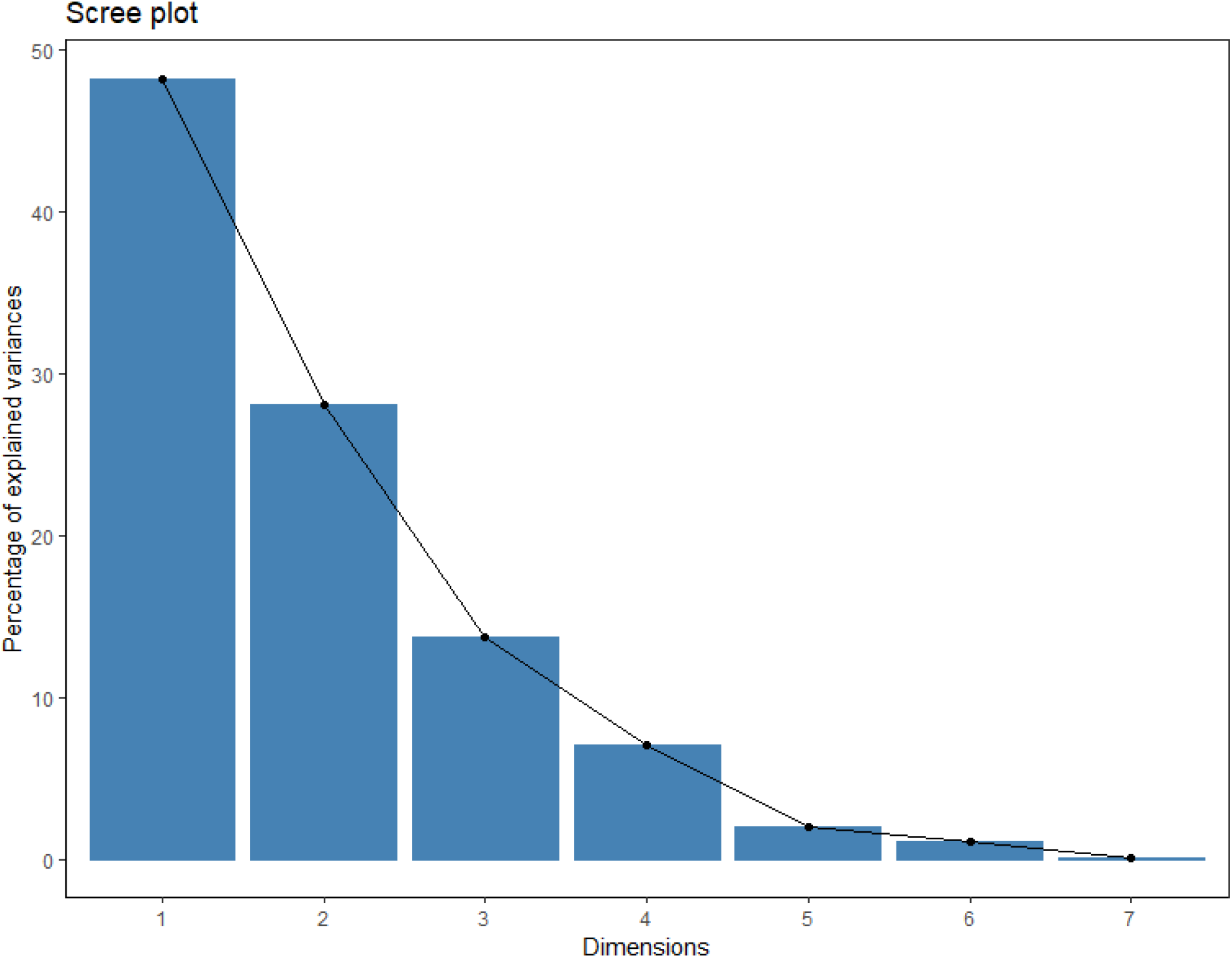
Eigen values illustrating the variance percentage explained by each component.

Figure B.10 illustrates the loadings for Principal Component 1 (PC1). The most significant contributions come from mean precipitation (-0.48), the aridity index (-0.43), and maximum precipitation (-0.40). The weights, therefore, indicate that the first principal component primarily captures variations related to aridity index and precipitation. By taking into account the sign of the loadings, we find that an increase in the environmental exposure index (PC1) is associated with a reduction in precipitation and increase in aridity (see Figure in appendix)

### 3.3. Model selection

We fitted the models in equations 2, 3 and 4 with and without an environmental exposure index covariate and compared the models using the Akaike Information Criteria (AIC). Among the three models, those with the covariate provided the lowest AIC. Furthermore, equation 3 with the environmental exposure index as a covariate had the lowest AIC for all 6 models. Next, the results presented are based on this model only (see equation 3).

### 3.4. Model validation

The model validation results are presented in the appendix. In this analysis, 87% (n=67) of the district models demonstrated a satisfactory fit to the observed data. However, the initial model, which assumed three outbreak intensity functions (OIFs) per district, did not perform well for ten districts, each of which experienced more than three distinct outbreak events. To address this, we incrementally increased the number of modelled OIFs. Incorporating four OIFs led to satisfactory fits for Banke, Kailali, Makawanpur and Rautahat. For Jhapa, Kapilbastu, Nawalparasi West, and Parsa models with five OIFs produced good fits. Finally, Chitawan and Rupandehi required that six outbreak intensity functions be added in order to achieve satisfactory model validation. Detailed model results for these ten districts are provided in the Appendix.

## 4. Association between the environmental exposure index and dengue fever incidence

The appendix provides estimates and confidence intervals of the environmental exposure index. In general, an increase in the environmental exposure index was associated with an increase in the incidence of dengue in 34 districts and a decrease in 43 districts. However, the results for 82% (n=63) districts were not statistically significant as the confidence intervals for the coefficients span the null hypothesis value of zero.

### 4.1. Characterizing the dengue outbreak intensity functions in Nepal

Figure 5 shows the outbreak years in each district after adjusting for the effect of the environmental exposure index. Out of the 67 districts with 3 outbreak intensity functions (OIFs), most districts experienced their first OIF (*µ*_1_) in 2017 (16%), the second OIF (*µ*_2_) in 2019 (65%) and the third OIF (*µ*_3_) in 2022 (95%).

**Figure 5:**
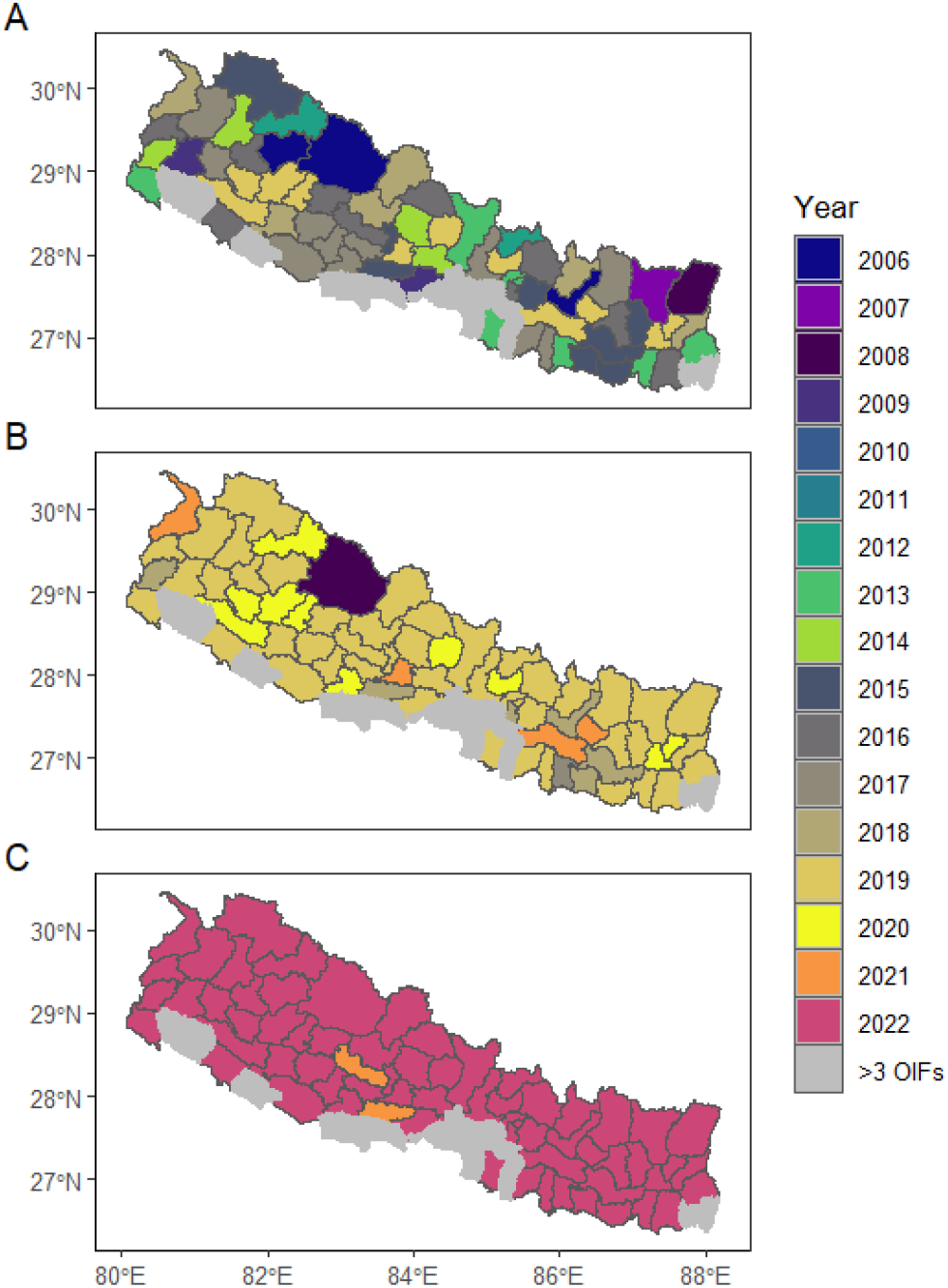
Maps showing the years that the districts had their first (A), second (B) and third outbreak intensity functions (C).

The estimate of the scale parameter (the duration) of the 3 OIFs also varied within each district. Figure 6 illustrates the scale parameter of three distinct OIFs across the 67 Nepalese districts with 3 OIFs. For the first outbreak, 87% of the 67 districts experienced it for less than a year, while 13% had it for a year or more. The second outbreak affected 69% of the districts for less than a year and 31% for more than a year. For the third outbreak, 79% of the districts experienced it for less than a year, with 12% affected for more than 2 years.

**Figure 6:**
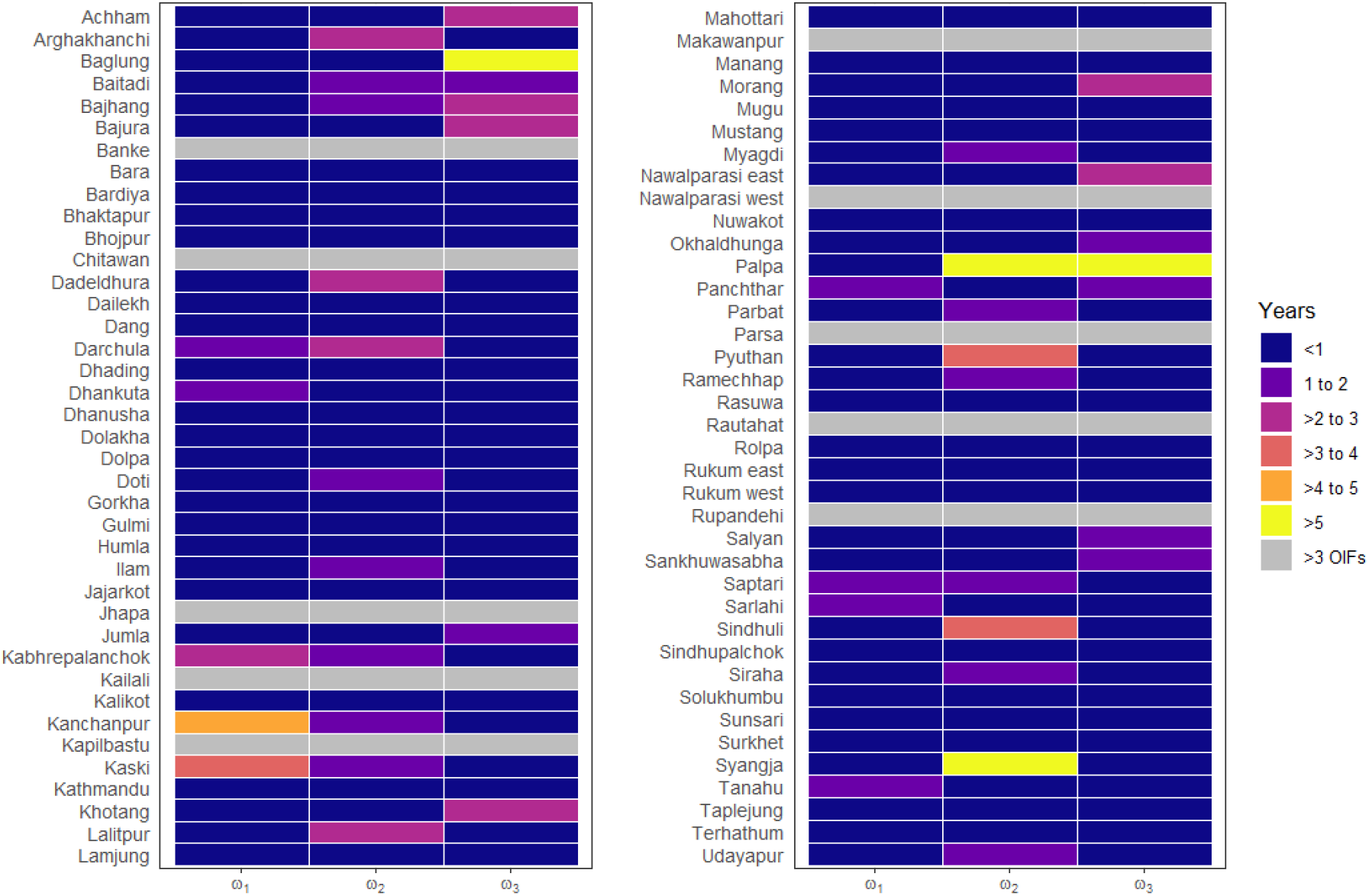
Heat maps showing the scale parameter (*ω*’s) of each outbreak intensity function in each district.

The largest coefficient for 96% (n=64) of the 67 districts with 3 OIFs was associated with the third OIF (*γ*_3_), followed by the second (*γ*_2_), and then the first (*γ*_1_). Figure 7 illustrates the approximate contribution of each OIF to the overall dengue epidemic in Nepal. Overall, the two highest contributions to the dengue epidemic in the 72 districts were from the second (15%, n = 10) and third OIFs (81%, n = 54). However, this result should be cautiously interpreted, as we did not constrain the outbreak coefficients (*γ*’s in equation 3) in our study.

**Figure 7:**
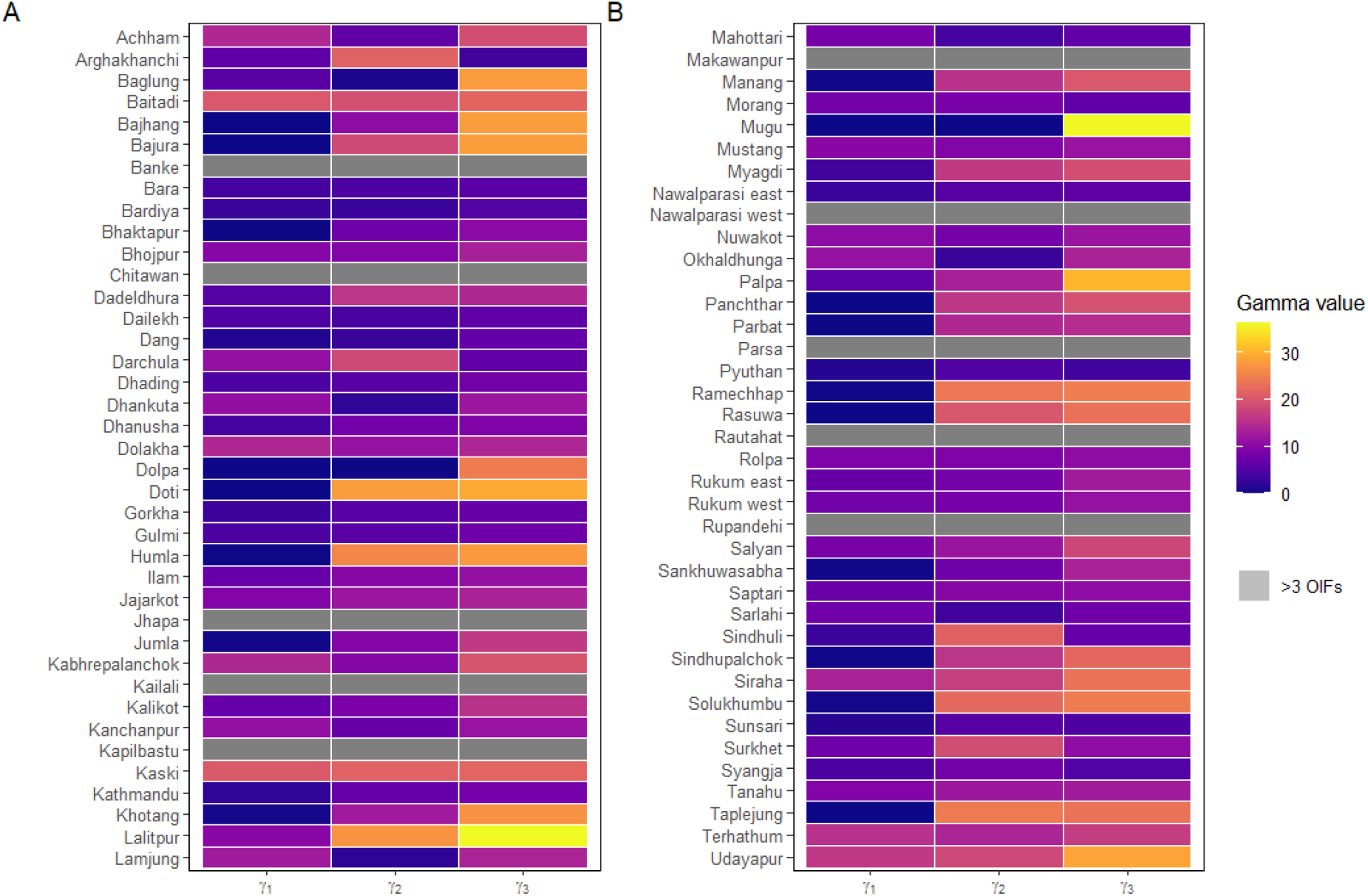
Heat maps showing the estimated coefficients (*γ*’s) for each outbreak intensity function.

The outbreak parameters (*µ, ω* and *γ*) in our models reveal distinct patterns of dengue incidence in Nepal’s districts. Districts such as Okhaldunga, Sindhuli and Syangja experienced more recent OIFs as characterized by higher values of *µ*_1_ and *µ*_2_ associated with later time functions *f*_(*i*,*t*)_. Furthermore, higher values of the outbreak coefficients (*γ*) in these districts indicate that the dengue epidemic has intensified in recent years. Conversely, districts such as Kailali and Nawalparasi West experienced more longstanding OIFs, as evidenced by the high values of the scale parameter of the outbreak intensity function (*ω*) across multiple OIFs, suggesting a prolonged epidemic with sustained cases over time. These results imply a heterogeneity of dengue dynamics across Nepal, with some districts facing newer OIFs while others have endured more extensive and persistent epidemics over the study period.

As noted above, ten districts experienced more than three dengue OIFs during the study period. Figure 8 presents the estimated timing of the OIFs’ peak for up to six OIFs (*µ*_1_ to *µ*_6_) in these districts between 2006 and 2022. Each colored cell represents the year of an identified outbreak, while gray cells (“ N/A”) indicate that no additional OIFs were estimated. The figure shows that most OIFs occurred between 2017 and 2022, although a few districts experienced earlier events in 2006 and 2010. Notably, nine of the ten districts experienced their most recent outbreak in 2022.

**Figure 8:**
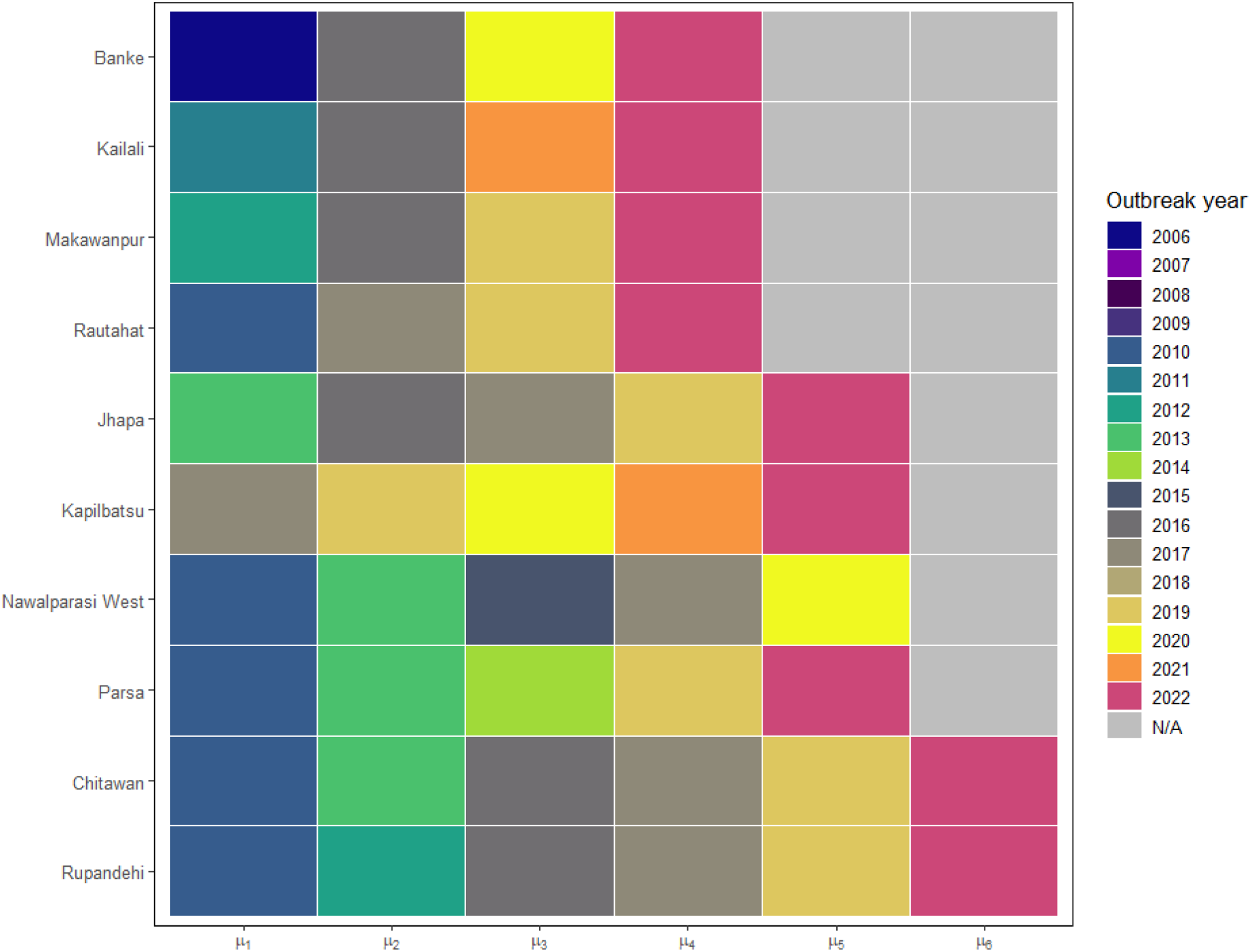
Heat map showing the years that districts with more than three outbreak intensity functions (OIFs) experienced each OIF (up to six OIFs)

## 5. Discussion and Conclusion

In this study, we analyzed dengue outbreak intensity functions (OIFs) in Nepal and extended the Negative Binomial regression model to allow the inclusion of an OIF with parameters that directly express the time of the outbreak peak and scale parameter of the OIF. To our knowledge, this is the first study to characterize dengue in Nepal using a model-based approach. Additionally, the proposed modelling framework effectively accommodates the occurrence of multiple OIFs within a district. While models such as HHH4 and HHH4ZI are valuable for decomposing outbreak data into endemic and epidemic components (Meyer et al., 2016; Lu and Meyer, 2023), they do not focus on estimating the scale of multiple OIFs. Similarly, other approaches identify clusters in space, but does not estimate the timing of the peak (Anderson et al., 2014). Additionally, other previous methods require data to be aggregated into spatio-temporal blocks (e.g., at 1 week, 3 weeks, and 5 weeks) to investigate outbreaks within those blocks (Rincón, 2020). In contrast, our model does not require such aggregation and is capable of estimating both the scale and timing of individual OIFs’ peaks. Although it operates effectively with annual data, as shown in this study, the model would indeed benefit from more temporally granular data, such as monthly or weekly dengue counts, which were not available in this analysis. The proposed model also offers a further understanding of dengue outbreak dynamics by estimating both the scale parameter of each OIF and approximating the contribution of each OIF to the overall dengue epidemic.

Our findings indicate that the relationship between dengue incidence and the environmental exposure index varies across districts. This spatial heterogeneity aligns with previous studies that have reported both positive and negative associations between environmental factors—such as precipitation—and dengue incidence in different regions (Abdulsalam et al., 2022; Campbell et al., 2013; Phanitchat et al., 2019; Li et al., 1985; Rozilawati et al., 2007). In some districts, we estimated a scale parameter of the outbreak intensity function (OIF) approaching five years, suggesting sustained dengue transmission over extended periods. This persistence underscores the importance of strengthened surveillance systems and district-specific intervention strategies to disrupt ongoing transmission and mitigate the risk of future outbreaks.

Another key finding is the occurrence of multiple OIFs over time within a single district. Similar to the environmental exposure index, the peak and scale parameters of each OIF varied across districts. An important extension of this work would be to introduce covariates that could account for the heterogeneity in the peak and scale parameters of each OIF across districts. Incorporating district-level variables, such as nighttime light data as a proxy for urbanization, could help explain the observed differences in outbreak dynamics (Li et al., 2021). These covariates would enable us to model how factors such as urbanization, human mobility and climate conditions influence the timing and size of OIFs. However, we did not pursue this approach here due to the limited amount of data available, which constrains our ability to estimate the effects of these variables. With information at higher temporal resolution, future research could expand on the work carried out in this study by incorporating additional data to better understand the drivers of outbreak variation across regions.

If temporally disaggregated data, such as weekly case counts, were available, we could improve outbreak detection by identifying exceedances of the OIF *f*_*t*_ above a predefined threshold *c*. Specifically, an outbreak could be declared when *f*_*t*_ *> c*, and the predictive probability of an outbreak at time *t* could be expressed as 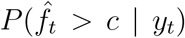, where 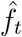 is the maximum likelihood estimator of the OIF. For instance, in the best-fitting model presented in our results, setting the threshold at *c* = log(2) implies that an outbreak is defined as a period during which the expected number of cases is at least twice the baseline level (i.e., when *f*_*t*_ = 0). However, with only annual data available, our ability to identify precise onset and end points of outbreaks is limited. Hence, we must restricted our description of outbreak dynamics to broad features such as peak timing, magnitude, and duration. Given more granular temporal data, another potential extension of the model proposed in this study would be to allow for asymmetric OIFs, such as those based on the skew-Normal distribution (Azzalini, 1985) which increase the flexibility of the model.

This study has several limitations. First, our reliance on passive surveillance data from the Ministry of Health introduces the likelihood of underreporting and misclassification of symptomatic dengue cases, a common issue in dengue surveillance systems (Pandey and Costello, 2019; da Fonseca and Fonseca, 2002; Gupta et al., 2018). Second, the analysis was limited by the unavailability of certain key risk factors - most notably, urbanization levels - which are known to influence the density of Aedes aegypti mosquitoes and, consequently, dengue transmission (Kolimenakis et al., 2021; Zeng et al., 2021a). Third, we were unable to assess the impact of lagged environmental variables, such as delayed effects of precipitation or temperature, due to the lack of temporally granular data (e.g., monthly observations). Future research should incorporate such lagged effects, as they have been shown to influence dengue dynamics in other contexts (Sugeno et al., 2023; Li et al., 2023). Finally, we did not detect evidence of residual spatial correlation in the current analysis (see Appendix); however, a natural extension of our modelling framework would be to incorporate spatially structured random effects, such as those used in the Besag–York–Mollié (BYM) model (Besag et al., 1991), to increase the predictive power of the model. However, this approach may require more detailed spatial information at the sub-district or ward level so as to better capture dependencies at smaller spatial scales.

In conclusion, we present a novel modelling framework that captures multiple OIFs over time in each district of Nepal. Applied to dengue, this model offers improvements over prior approaches, as it allows to use a data-driven approach to characterize timing of the OIFs’ peaks, scale parameters, and intensity without imposing assumptions about disease transmission dynamics, as is typically necessary in mechanistic compartmental models. The results could support the Nepalese government in identifying districts with synchronized outbreaks and developing targeted interventions. Moreover, the framework is adaptable to other infectious diseases, providing a tool for reconstructing historical outbreak dynamics and informing future public health responses.

## CRediT authorship contribution statement

JJ Khaki: Methodology, Data resources, Software, Analysis, Writing - Original Draft, Writing – review & editing. BK Acharya: Data resources, Writing – review & editing. BD Pandey: Conceptualization, Data resources, Writing – review & editing. K Morita: Conceptualization, Writing – review & editing. E Giorgi: Conceptualization, Methodology, Data resources, Software, Analysis, Supervision, Writing – review & editing. All authors have contributed to reviewing and editing of the manuscript.

## Data availability

All the sources for the data used in this study have been cited in the main manuscript. The R code used to run the models in this study can be accessed on *Github*.

## Appendix A. Exploratory analysis and variable processing

Figure A.9 shows scatter plots of the log of dengue incidence against several spatial covariates. The plot shows that most of the covariates had a linear relationship with the incidence of dengue.

**Figure A.9:**
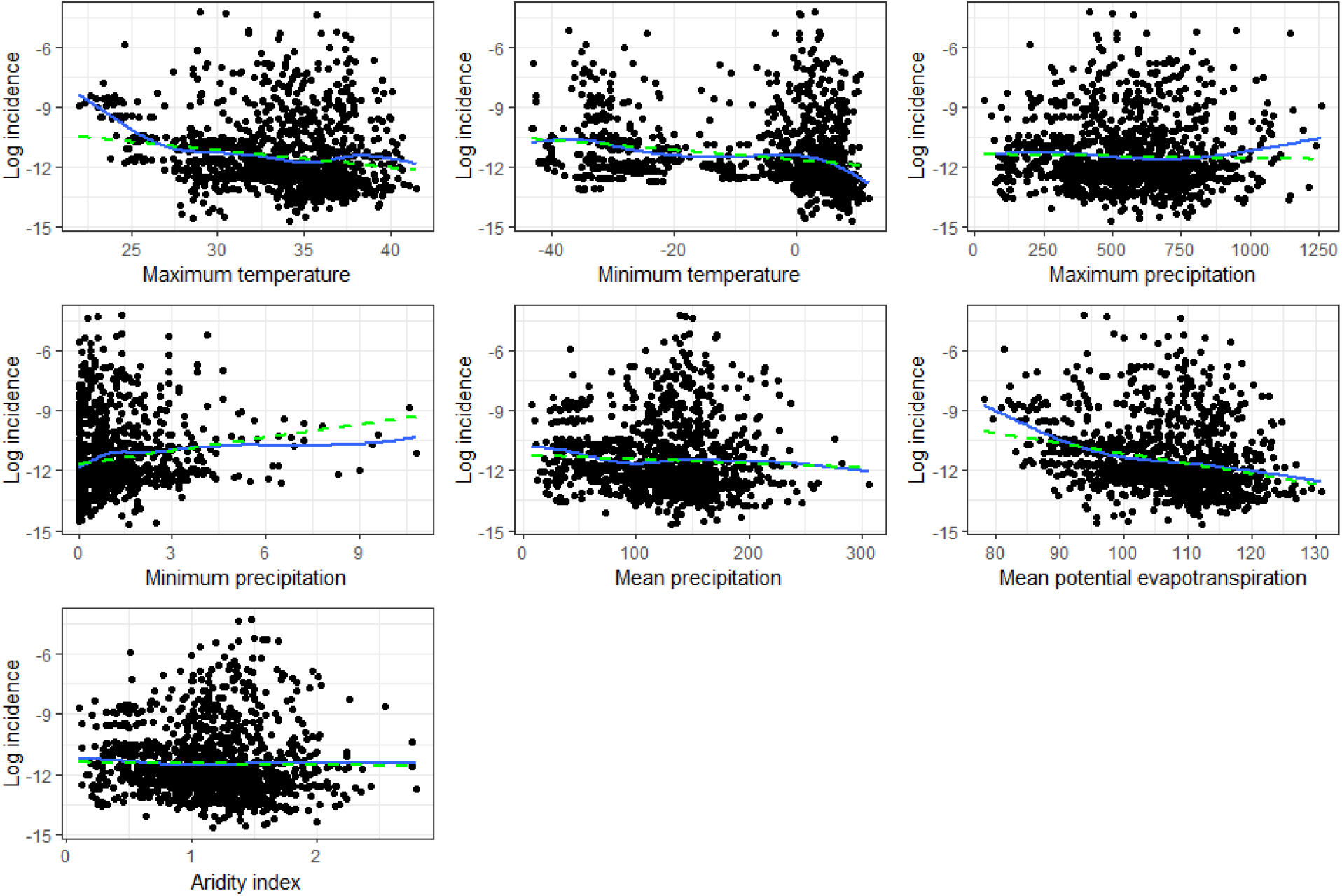
Scatter plots of the log incidence against maximum temperature, minimum temperature, maximum precipitation, minimum precipitation, mean precipitation, mean evapotranspiration, and aridity index. The dashed green lines are regression lines from a linear model, whilst the blue solid lines are natural splines from a generalized additive model.

## Appendix B. Principal components analysis further results

This section presents additional results from the Principal Component Analysis.

**Figure B.10:**
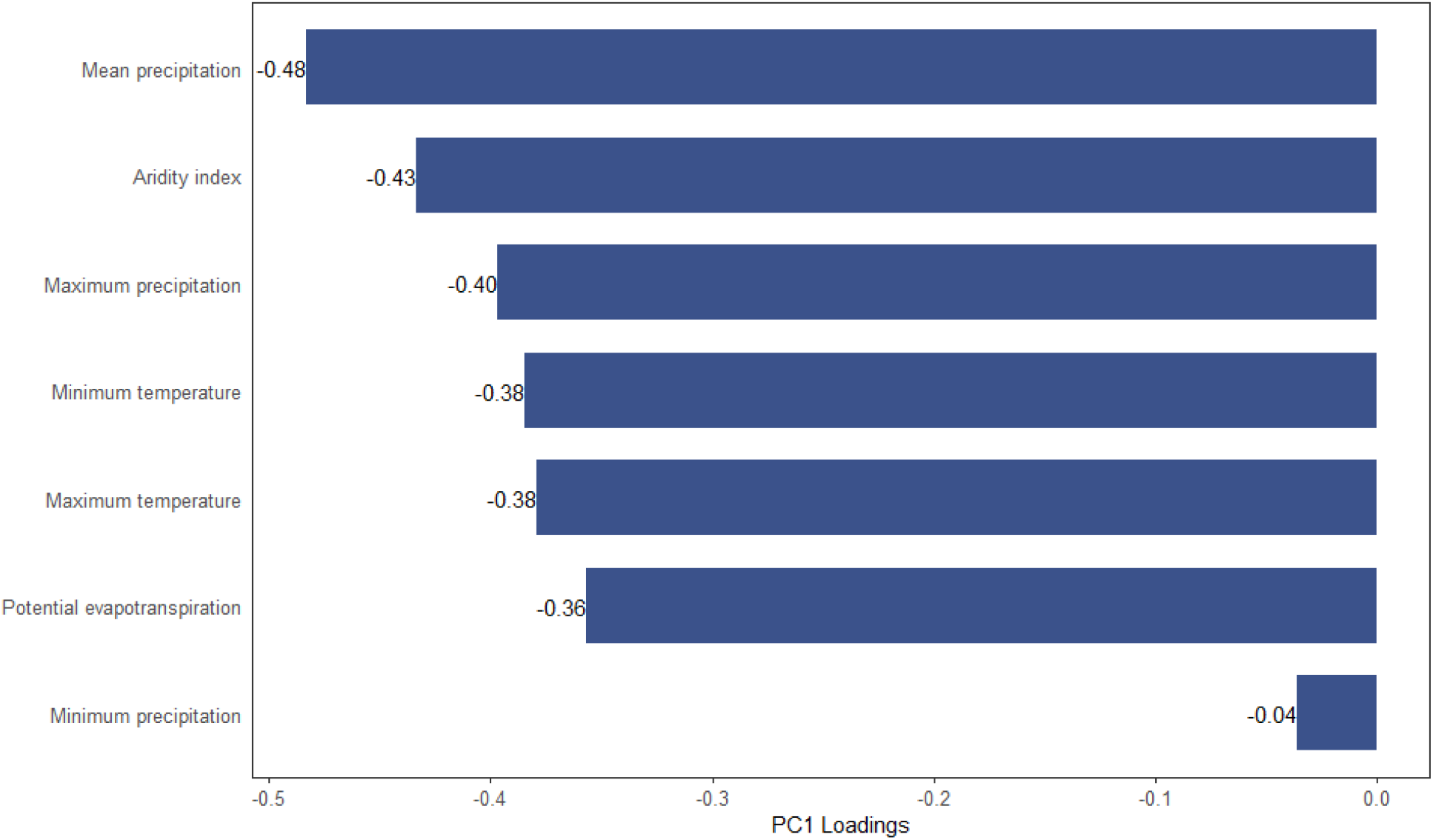
Loadings of the environmental exposure index (PC1)

**Figure B.11:**
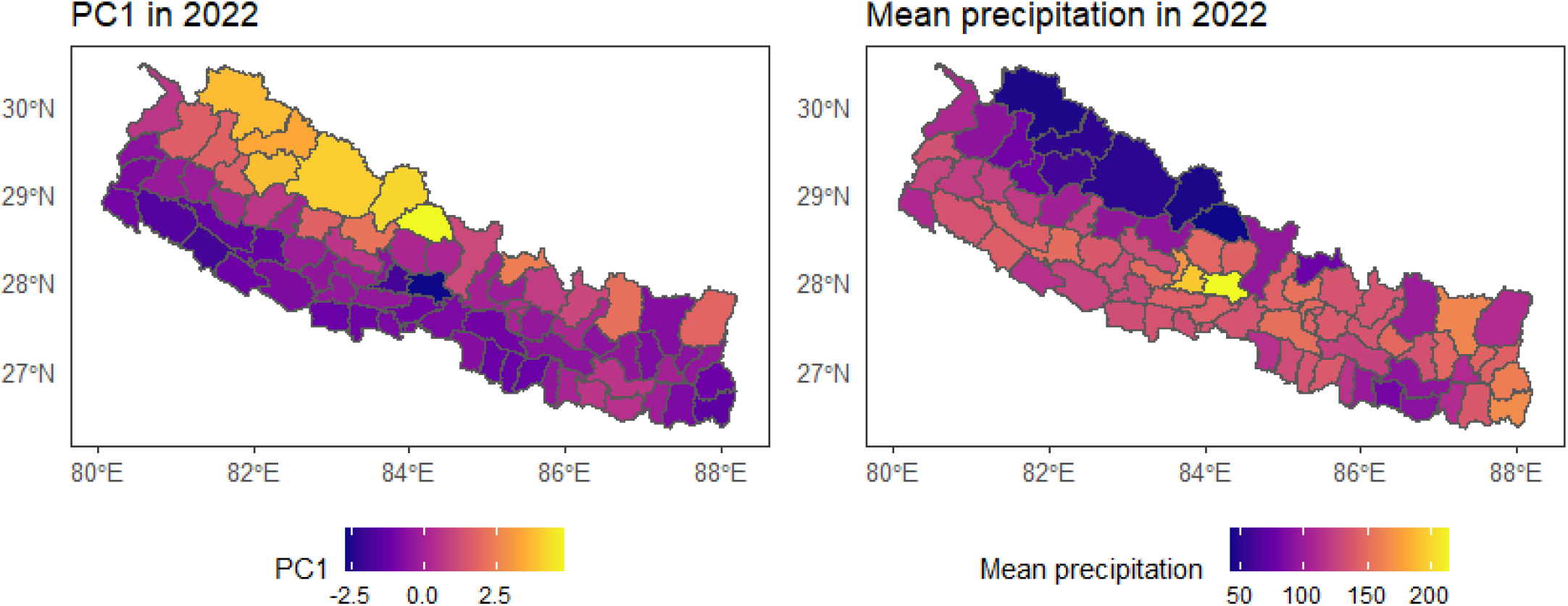
Maps of the environmental exposure index (PC1) and the mean precipitation

## Appendix C. Assessing the presence of residual spatial autocorrelation

We extracted the geographic centroids of all 77 districts in Nepal and used them as spatial locations for analysis. For each year from 2006 to 2022, we fitted a negative binomial regression model that adjusted for population size (as an offset) and included the environmental exposure index as a covariate. Pearson residuals from each model were extracted and used to assess residual spatial autocorrelation. Using district centroids as spatial coordinates, we computed empirical variograms of residuals for each year and generated corresponding simulation envelopes using 10,000 Monte Carlo permutations (Diggle and Giorgi, 2019). In all 17 years, the observed variogram (Figure C.12 and Figure C.13) remained entirely within the simulation envelope, indicating that there was no significant evidence of spatial autocorrelation in the model residuals across Nepal.

**Figure C.12:**
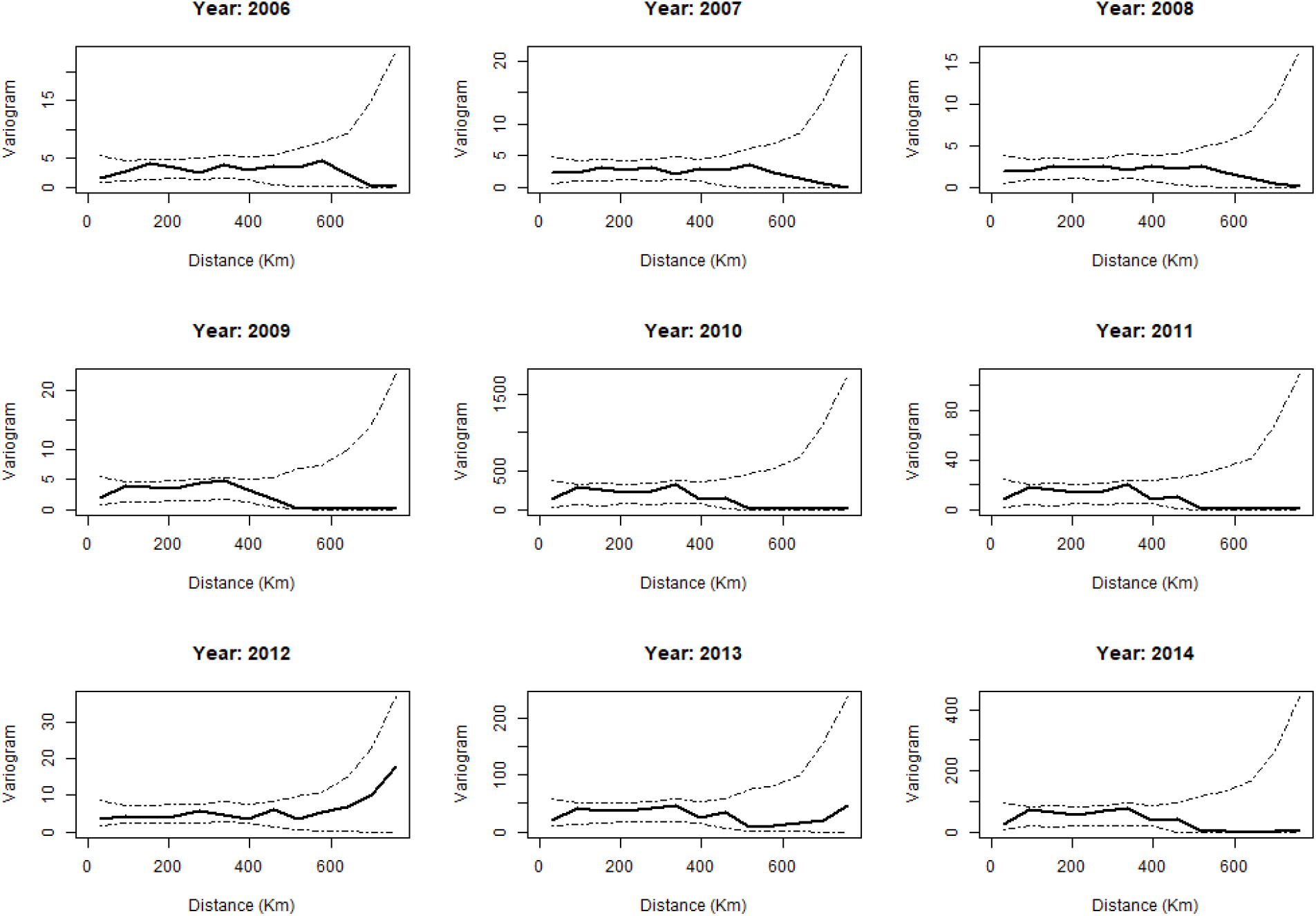
Empirical variograms of model residuals from 2006 to 2014, with 95% tolerance envelopes generated under the null hypothesis of no spatial autocorrelation.

**Figure C.13:**
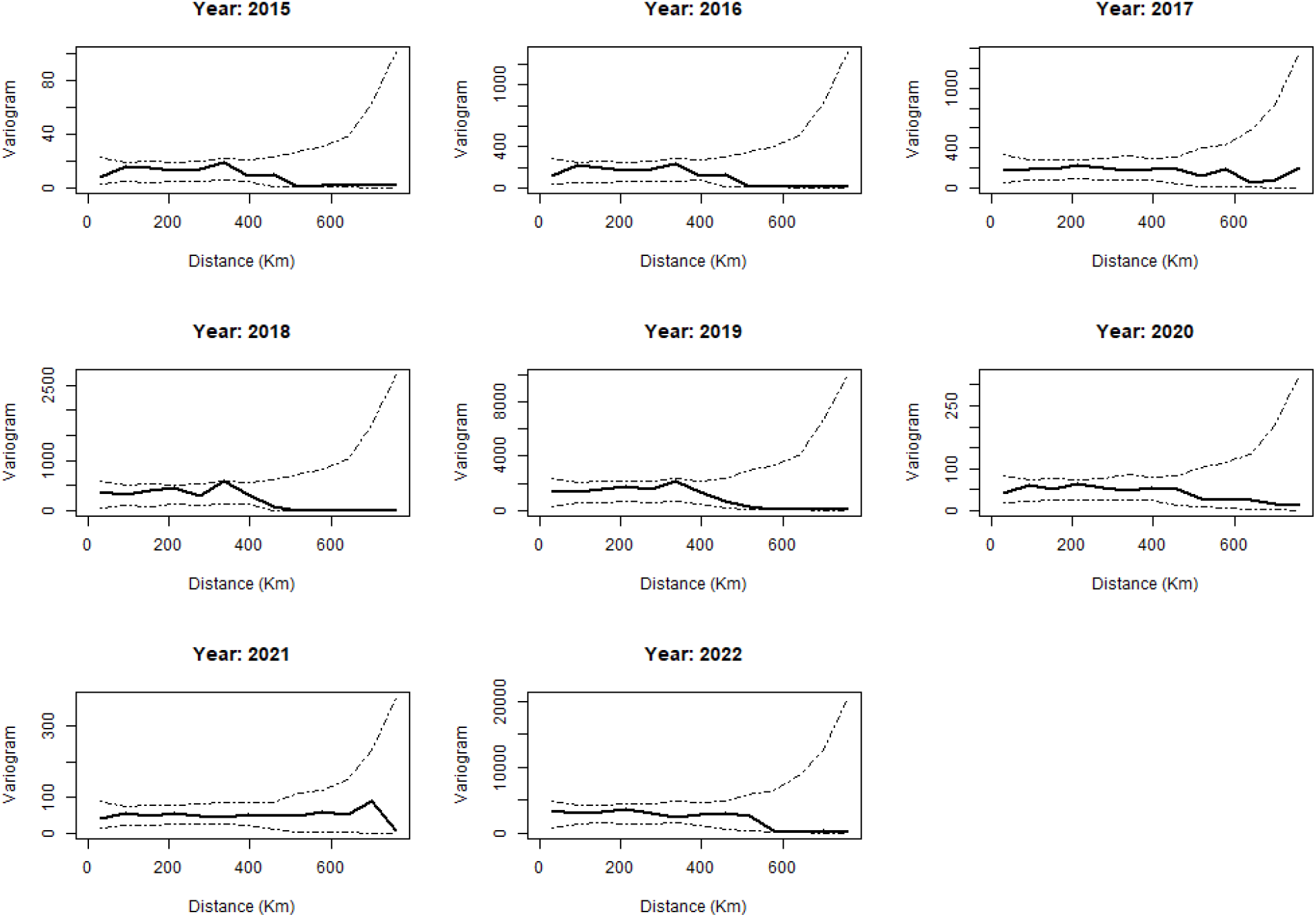
Empirical variograms of model residuals from 2015 to 2022, with 95% tolerance envelopes generated under the null hypothesis of no spatial autocorrelation.

Figures C.14, C.15, and C.16 illustrate the predicted timing of the peak (*µ*) of the three outbreak intensity functions (OIFs) and their confidence intervals (CIs).

**Figure C.14:**
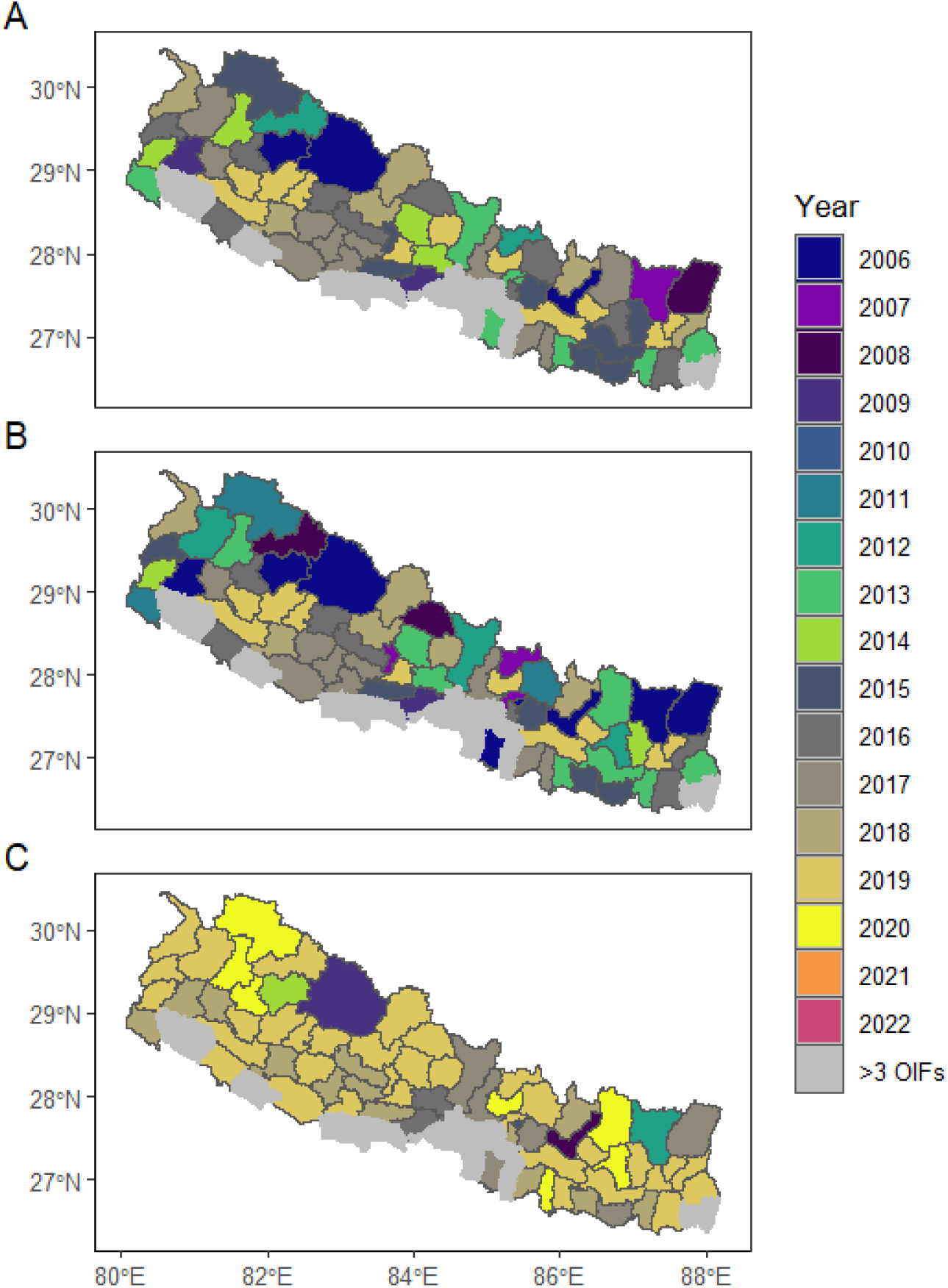
Maps showing the timing of the first outbreak peak (*µ*_1_, A), and the lower (B) and upper (C) bounds of the 95% CIs. Grey areas represent districts that experienced more than three OIFs.

**Figure C.15:**
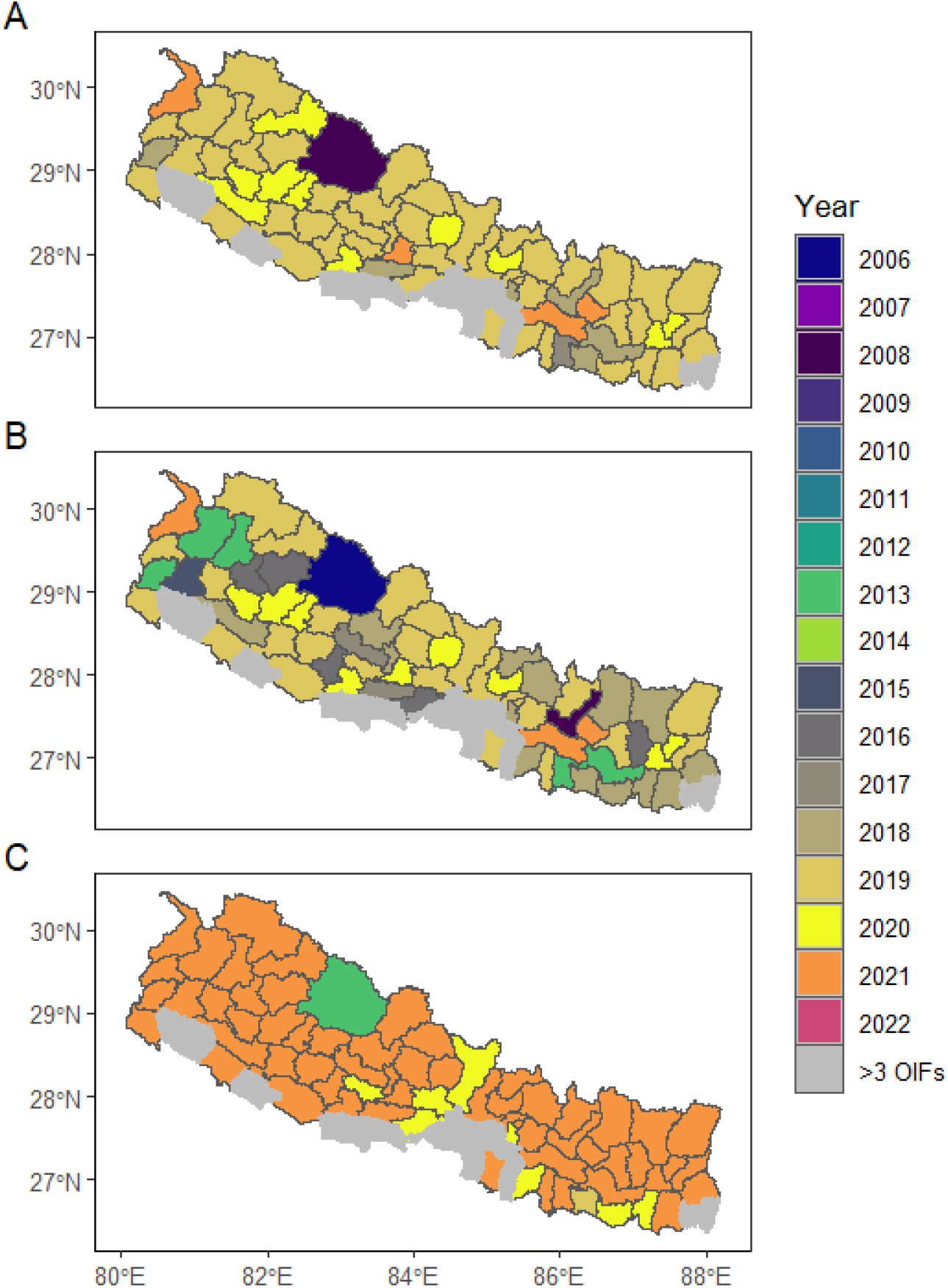
Maps showing the timing of the second outbreak peak (*µ*_2_, A), and the lower (B) and upper (C) bounds of the 95% CIs. Grey areas tiles represent districts that experienced more than three OIFs.

**Figure C.16:**
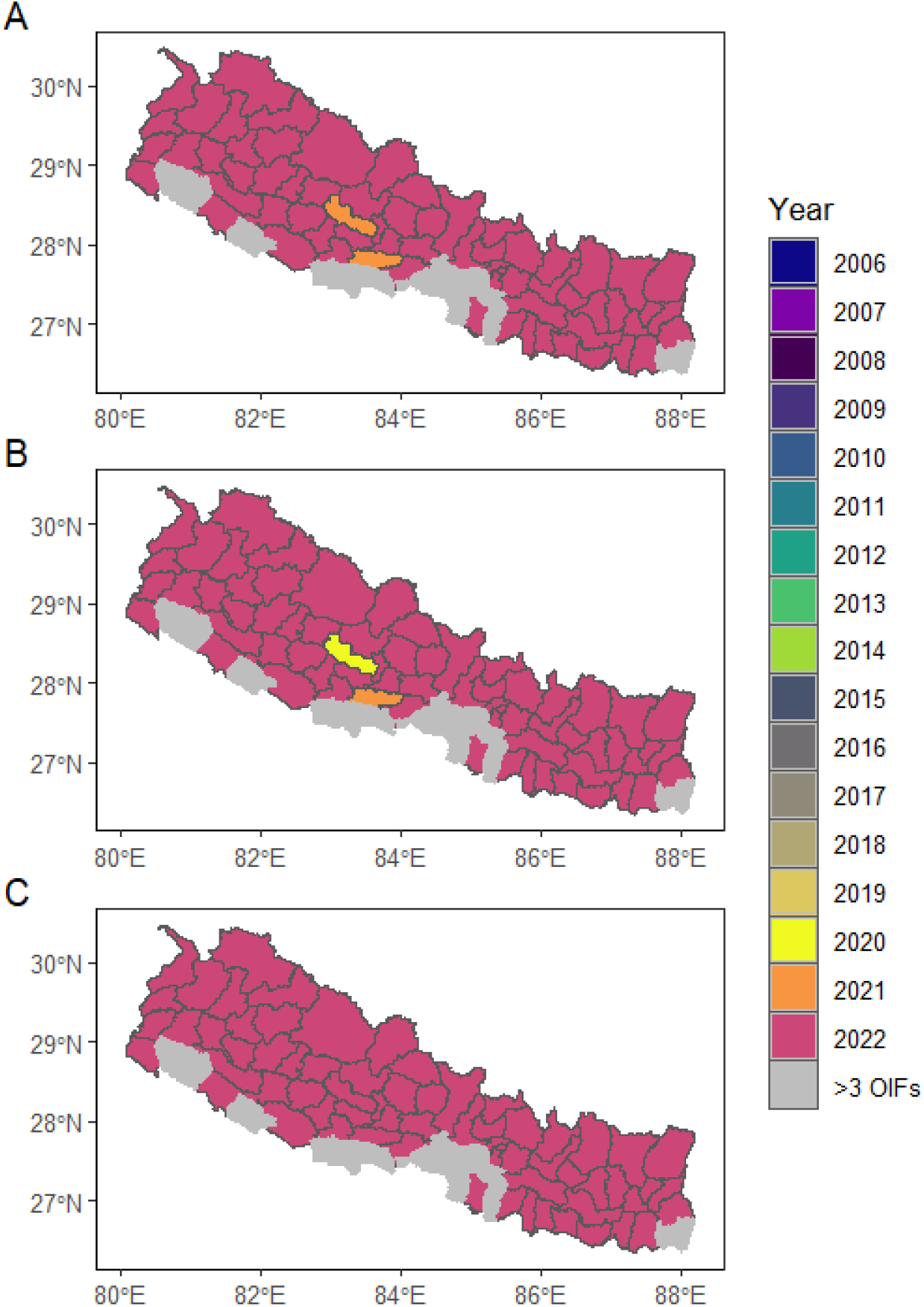
Maps showing the timing of the third outbreak peak (*µ*_3_, A), and the lower (B) and upper (C) bounds of the 95% CIs. Grey areas represent districts that experienced more than three OIFs.

Figures C.17, C.18, and C.19 illustrate the predicted scale parameters (*ω*) of the three outbreak intensity functions (OIFs) and their confidence intervals (CIs).

**Figure C.17:**
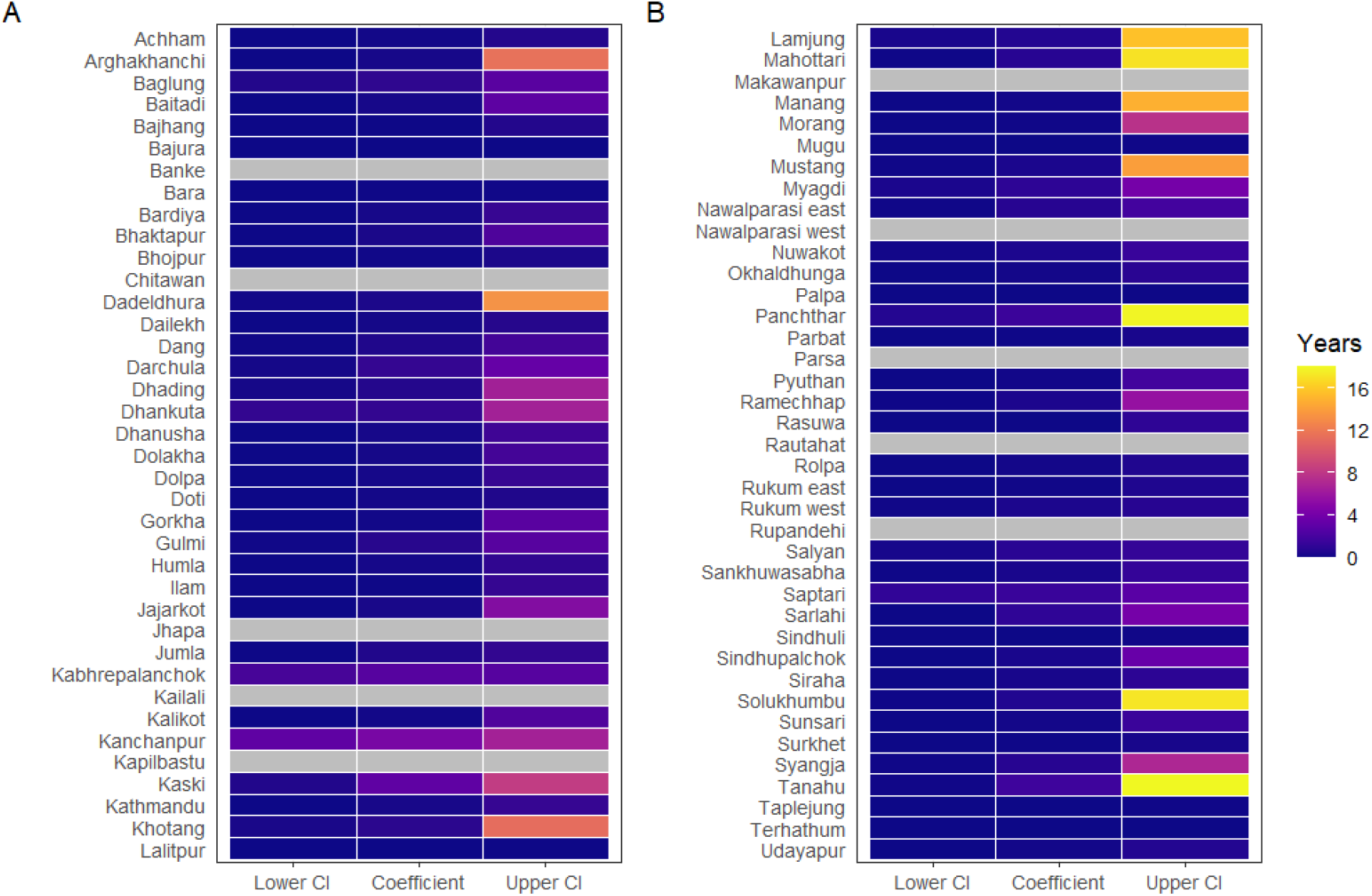
Heat maps showing the scale parameter of the first OIF (*ω*_1_), and the lower and upper CIs. Grey tiles represent districts that experienced more than three OIFs.

**Figure C.18:**
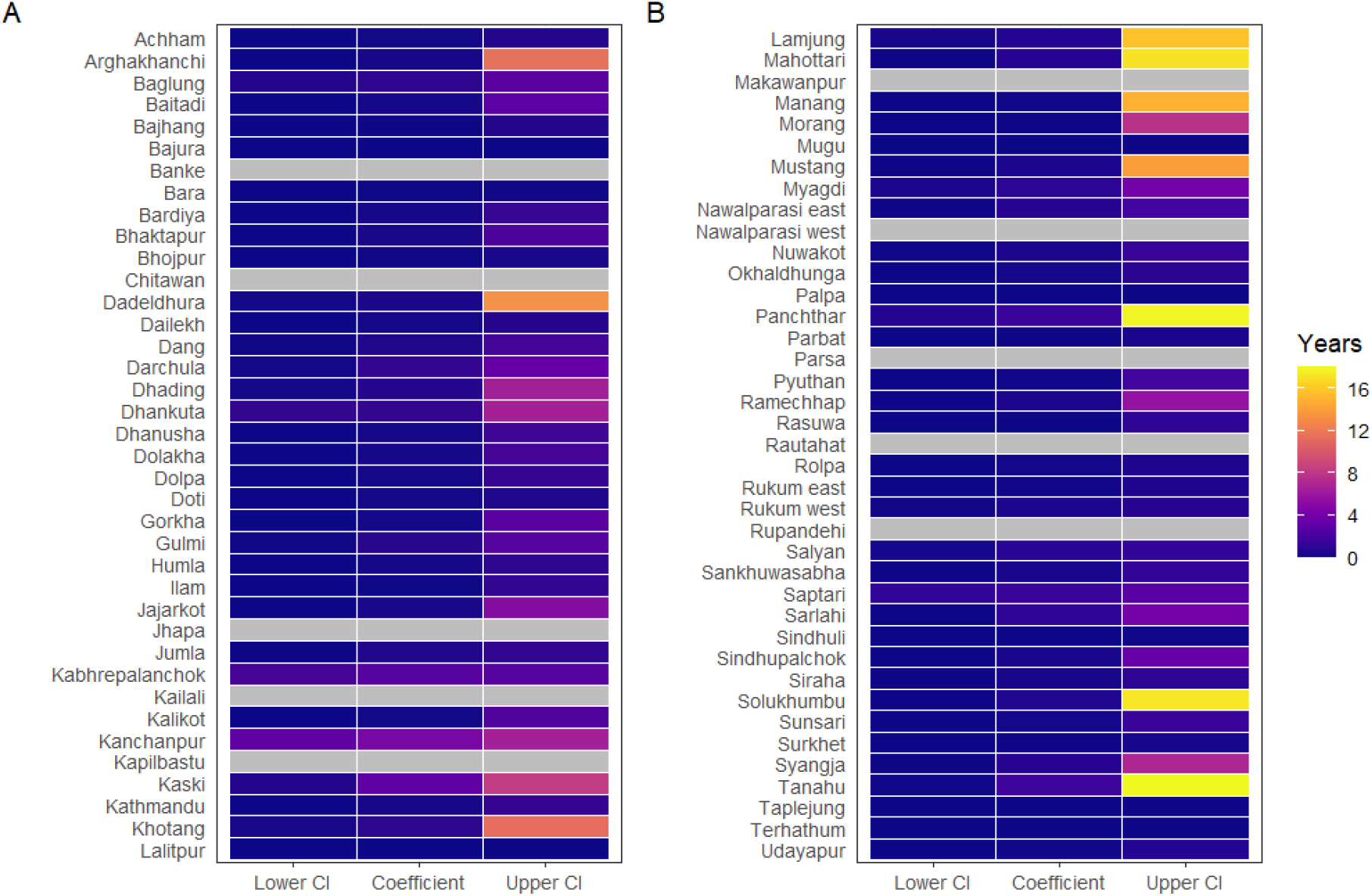
Heat maps showing the scale parameter of the second OIF (*ω*_2_), and the lower and upper CIs. Grey tiles represent districts that experienced more than three OIFs.

**Figure C.19:**
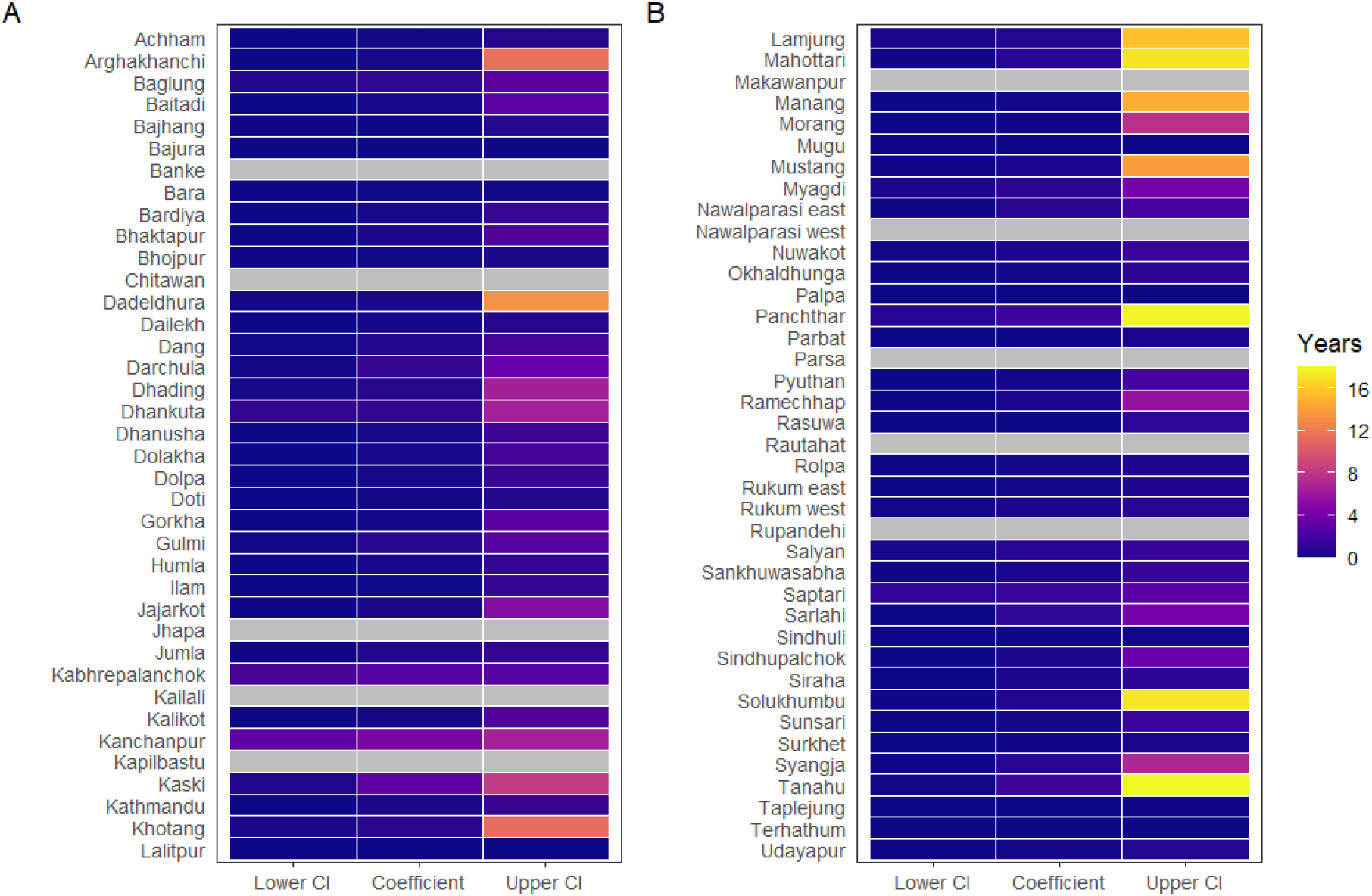
Heat maps showing the scale parameter of the third OIF (*ω*_3_), and the lower and upper CIs. Grey tiles represent districts that experienced more than three OIFs.

Figures C.20, C.21, and C.22 illustrate the estimated coefficients (*γ*) of the three outbreak intensity functions (OIFs) and their confidence intervals (CIs).

**Figure C.20:**
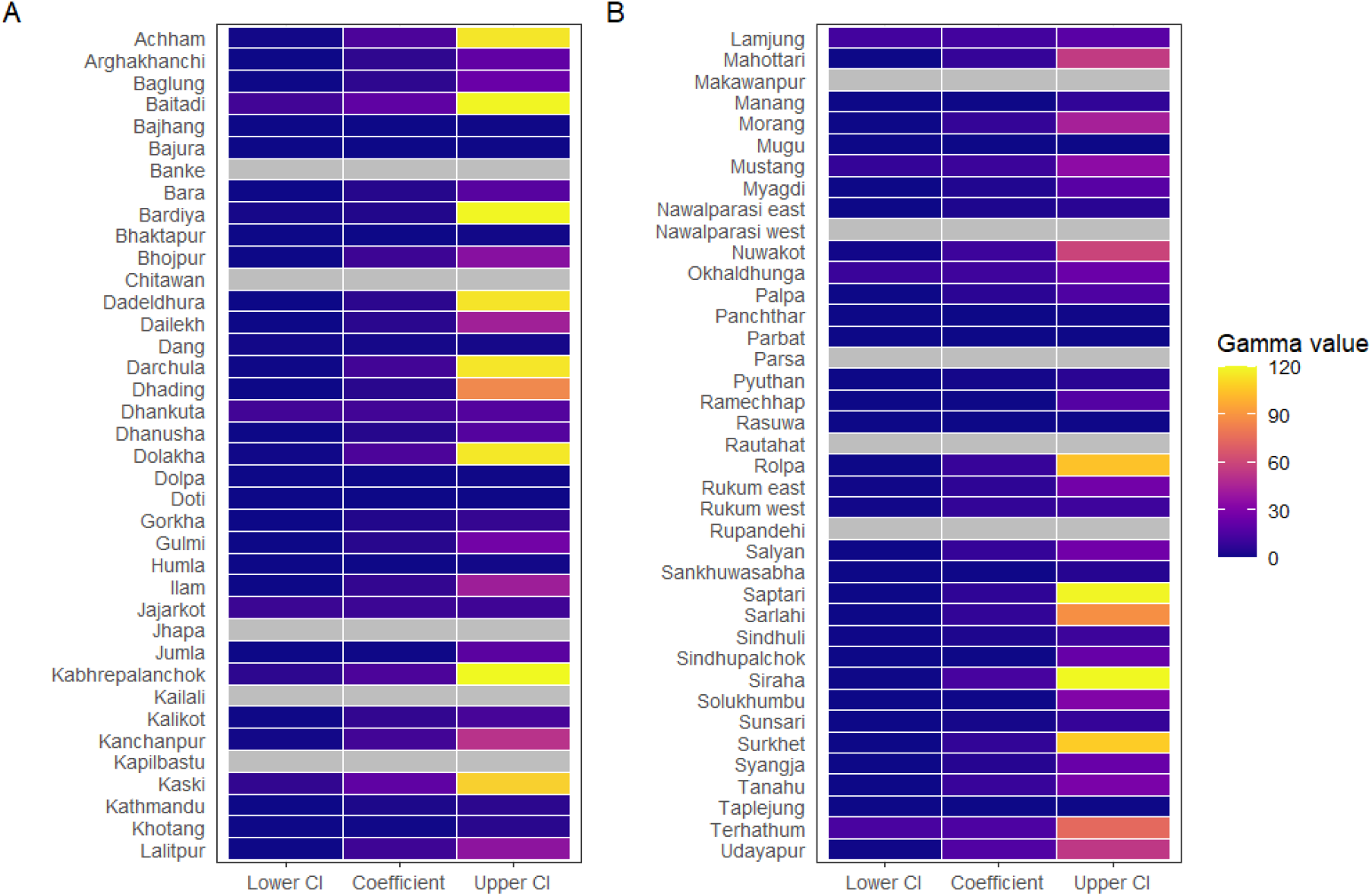
Heat maps showing the estimated coefficient of the first OIF (*γ*_1_), and the lower and upper CIs. Grey tiles represent districts that experienced more than three OIFs.

**Figure C.21:**
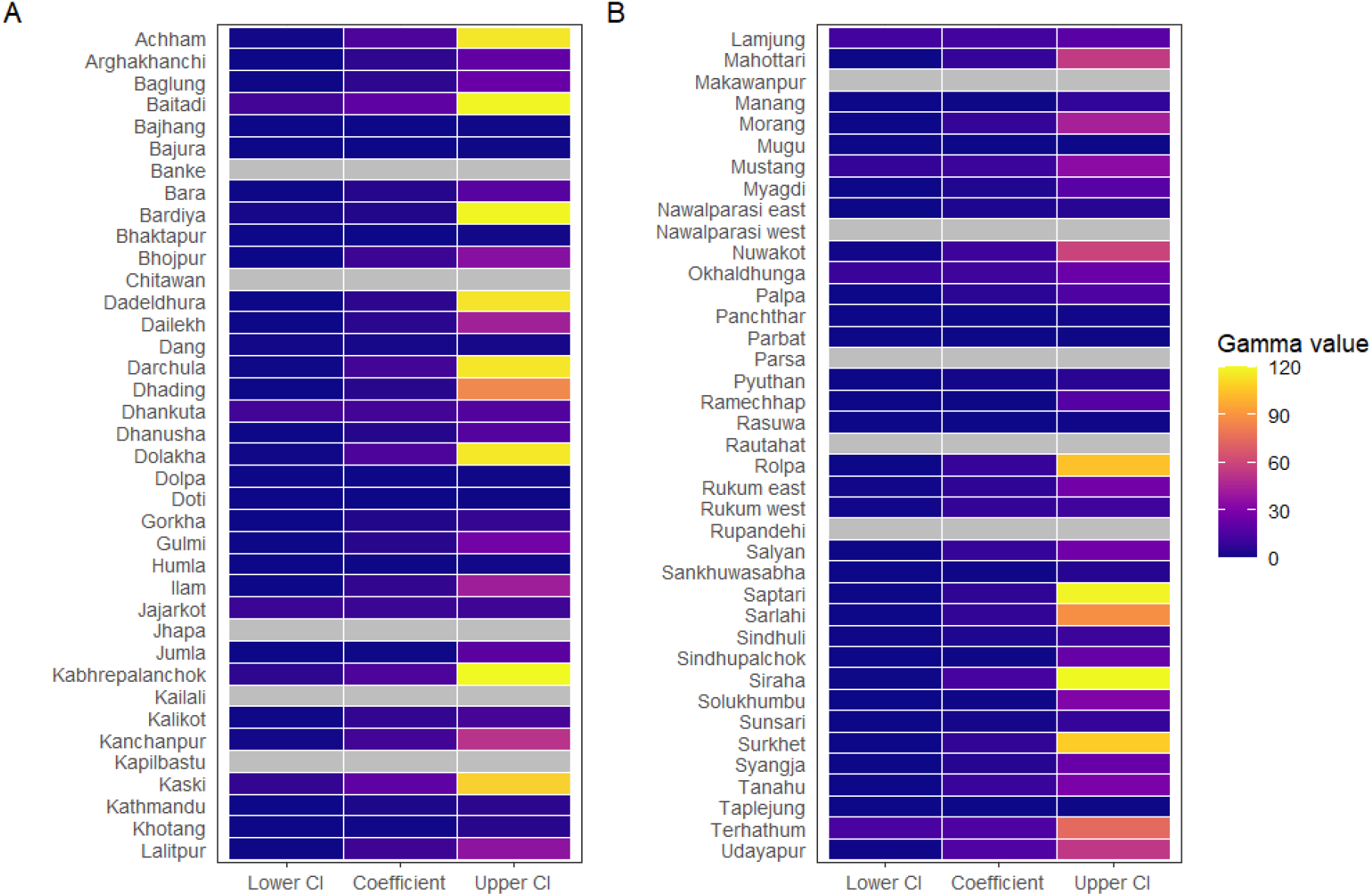
Heat maps showing the estimated coefficient of the second OIF (*γ*_2_), and the lower and upper CIs. Grey tiles represent districts that experienced more than three OIFs.

**Figure C.22:**
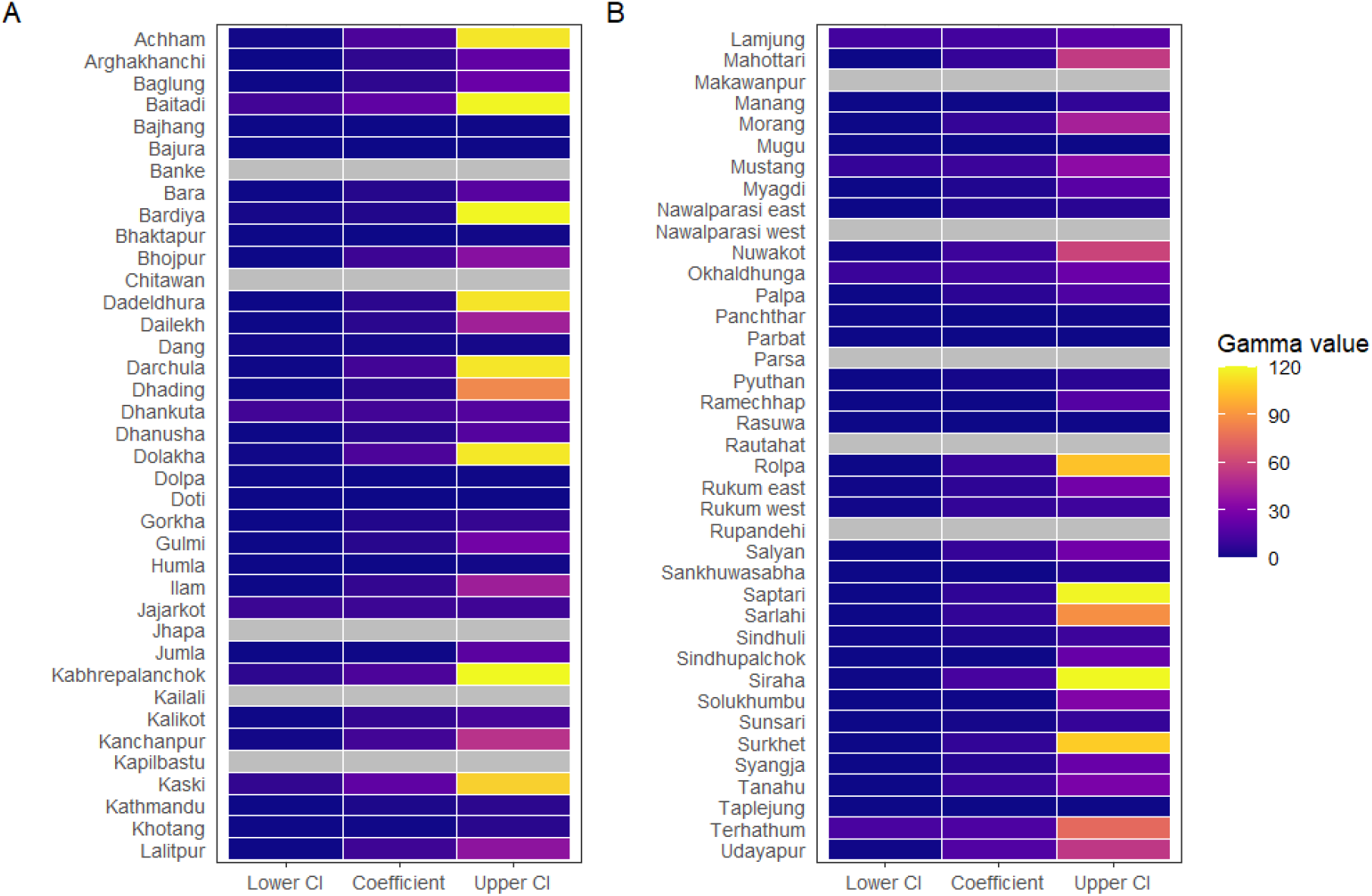
Heat maps showing the estimated coefficient of the third OIF (*γ*_3_), and the lower and upper CIs. Grey tiles represent districts that experienced more than three OIFs.

### Additional results for districts with more than 3 outbreak intensity functions

**Figure C.23:**
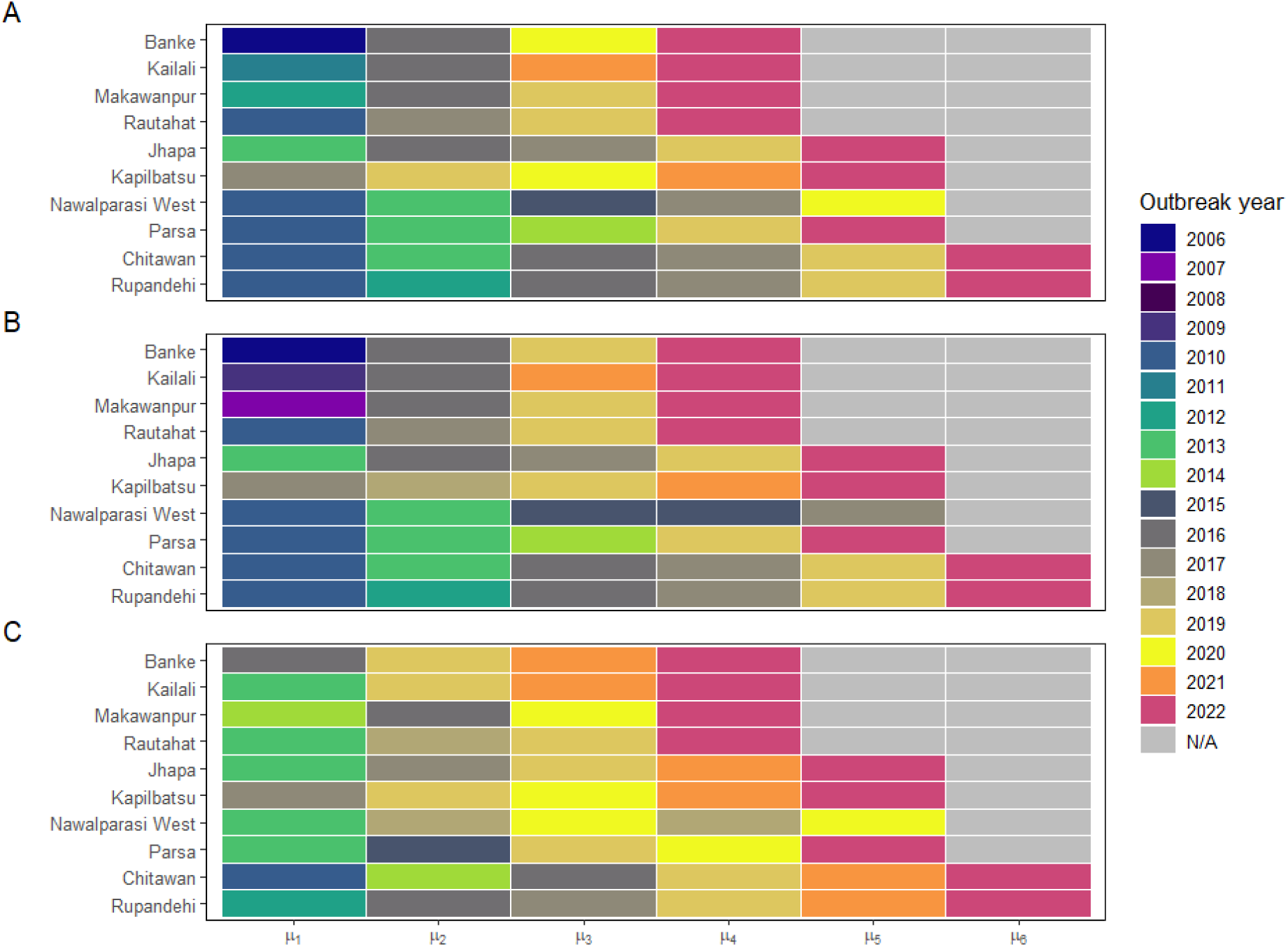
Heat maps showing the timing of outbreak intensity function peaks (A), and the lower (B) and upper (C) bounds of the 95% CIs.

**Figure C.24:**
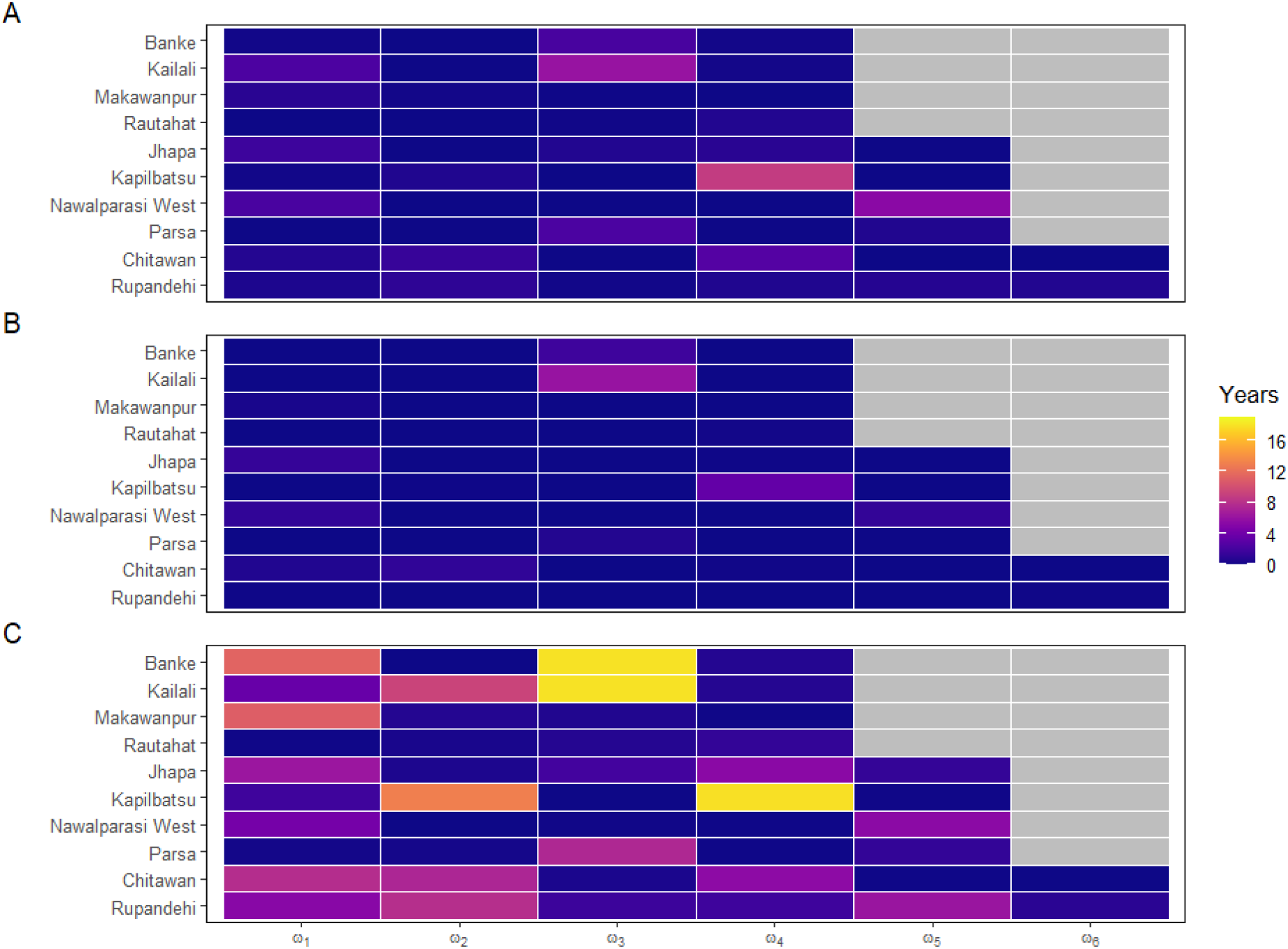
Heat maps showing the scale parameters of OIF (A), and the lower (B) and upper (C) CIs.

**Figure C.25:**
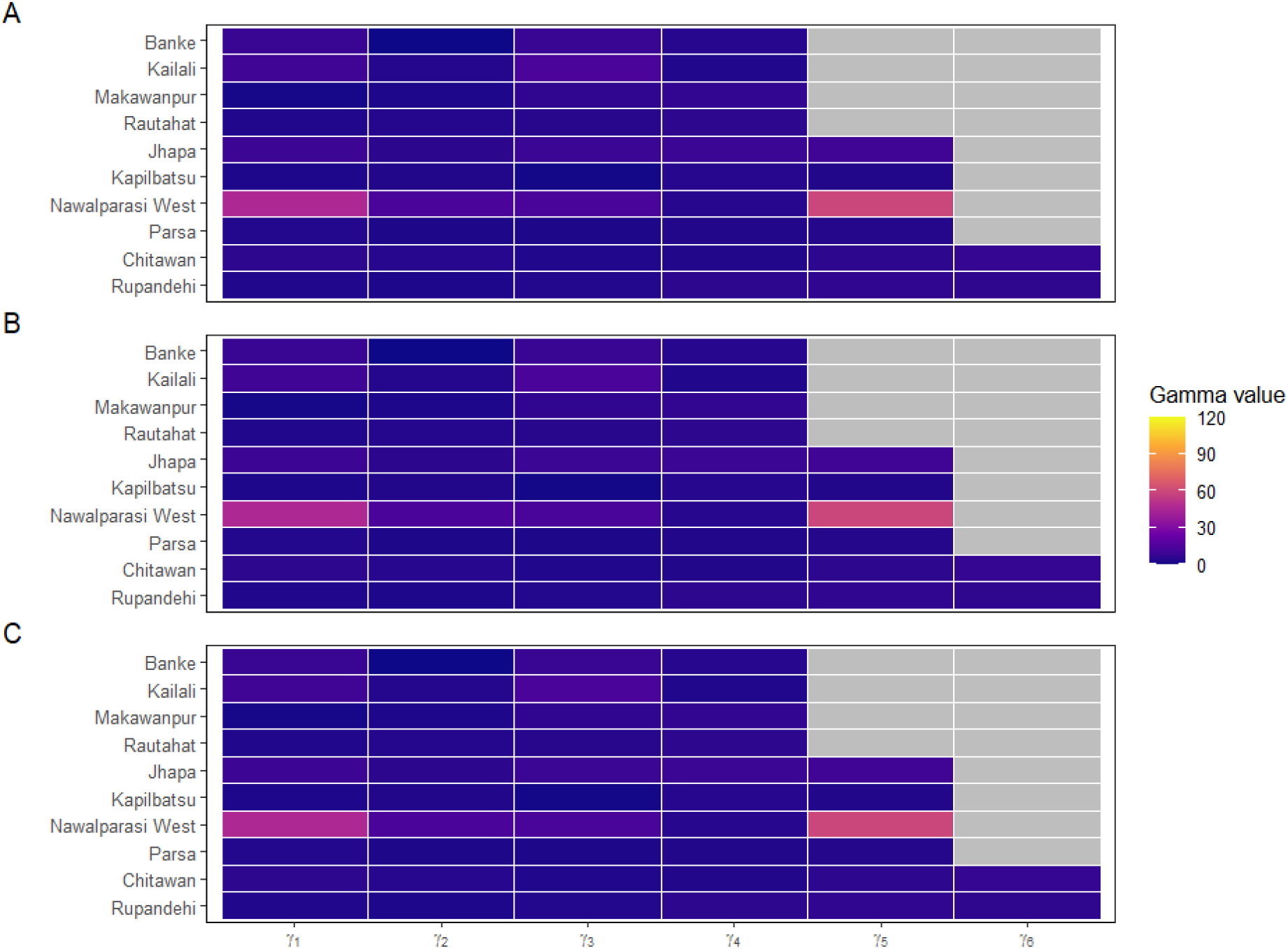
Heat maps showing the estimated coefficient (A) of OIF, and the lower(B) and upper (C) CIs.

## Appendix D. Model validation

We carried out a Chi-square (*χ*^2^) goodness of fit test for each of the models fitted to each district. The goodness of fit test was carried out under the null hypothesis that there was no statistically significant difference between the expected and predicted dengue counts. Each of the tests had 16 degrees of freedom (df = n-1 = 17-1) Table D.3 shows the Chi-square statistic and p-values for each district.

**Table D.3:**
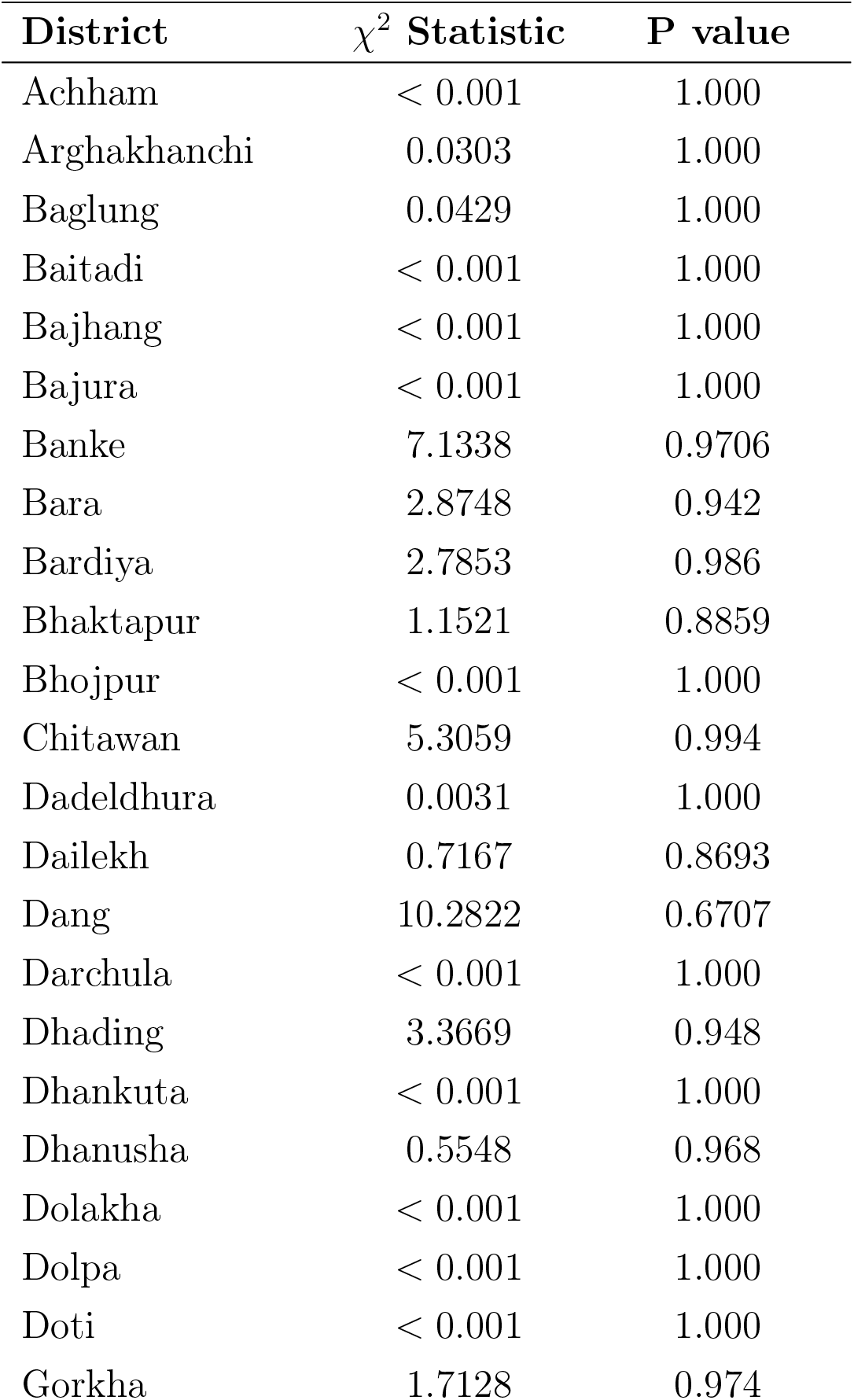
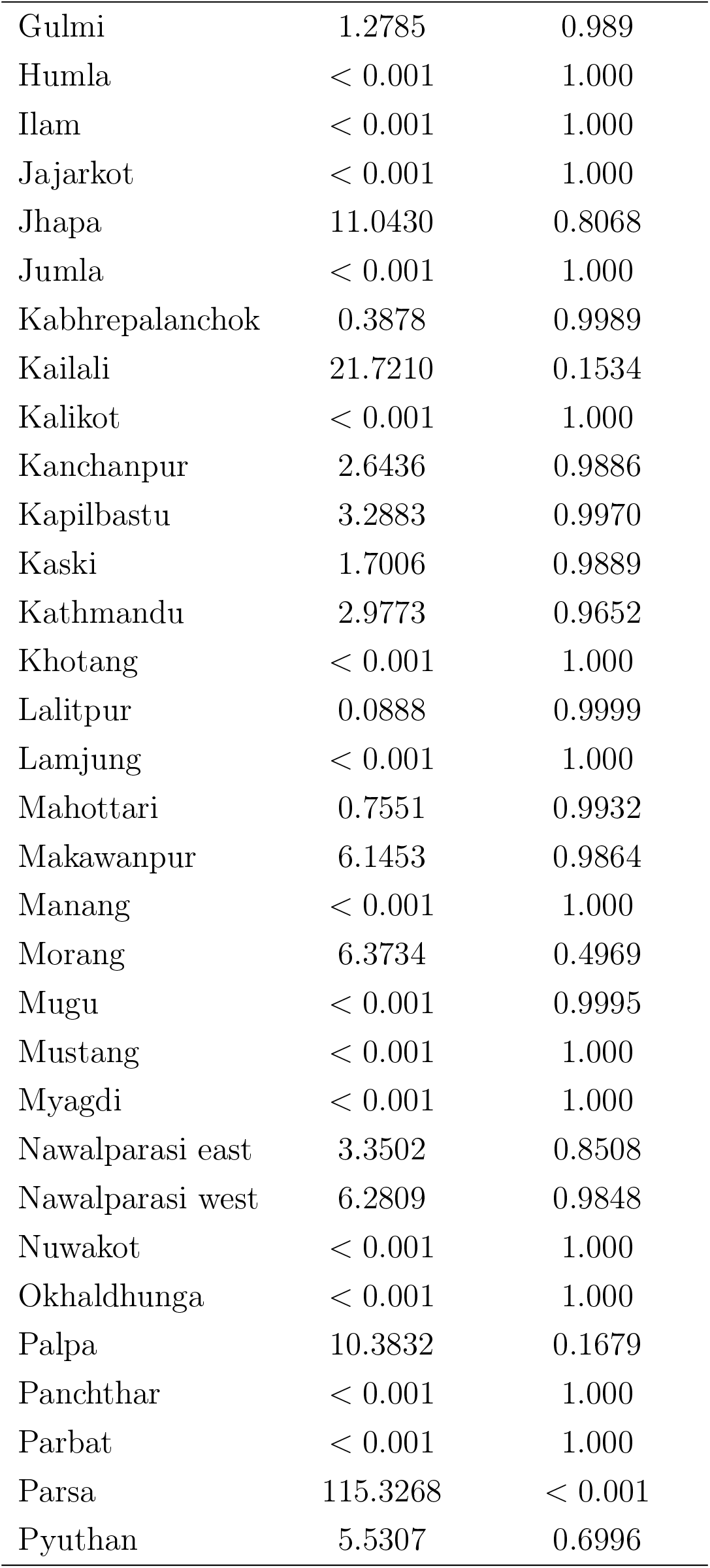
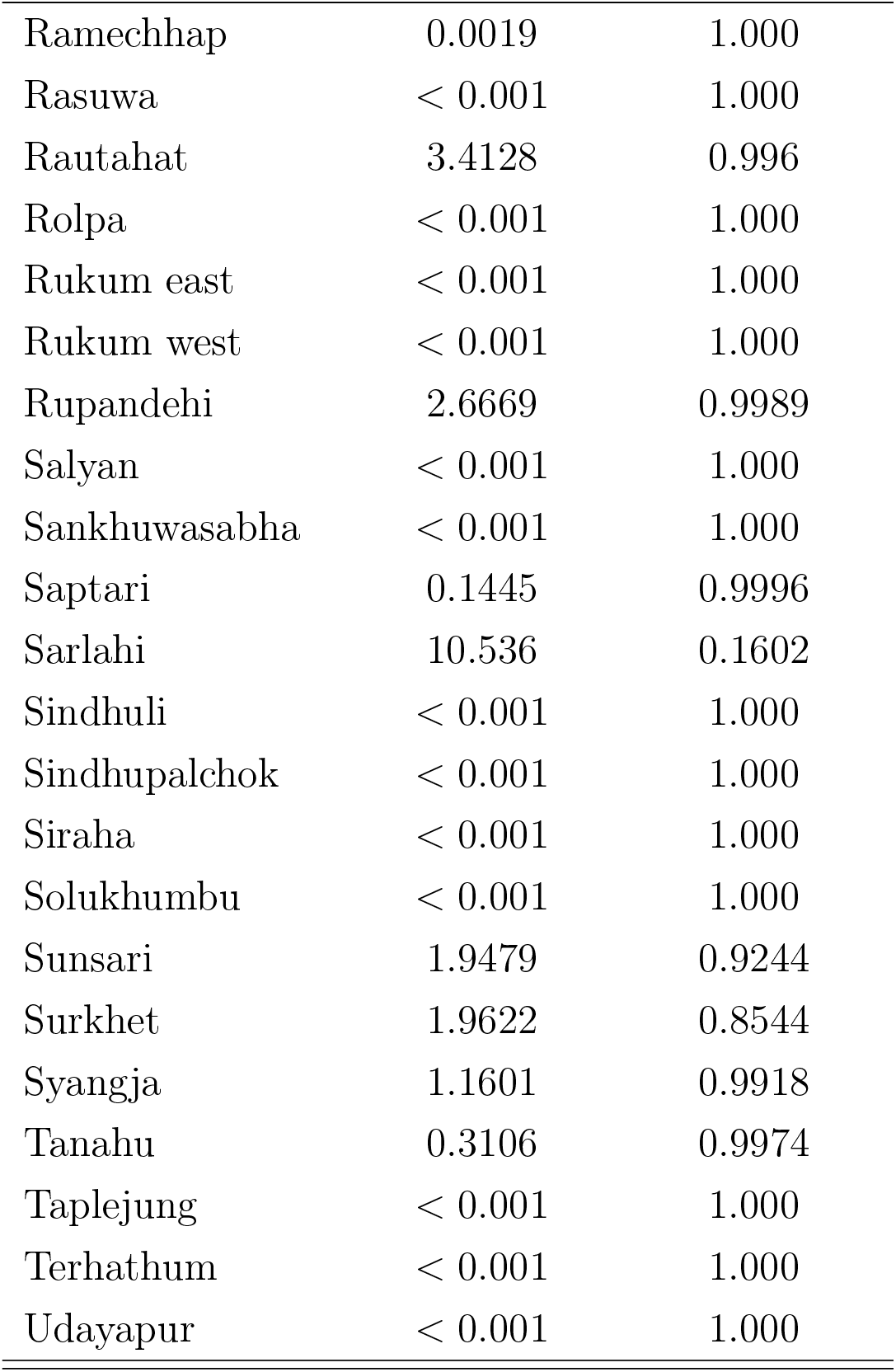
Summary of Chi-square (*χ*^2^) goodness of fit test results for the 77 Nepalese districts.

The figures below show the plots of the observed vs predicted counts of dengue cases in each district.

**Figure D.26:**
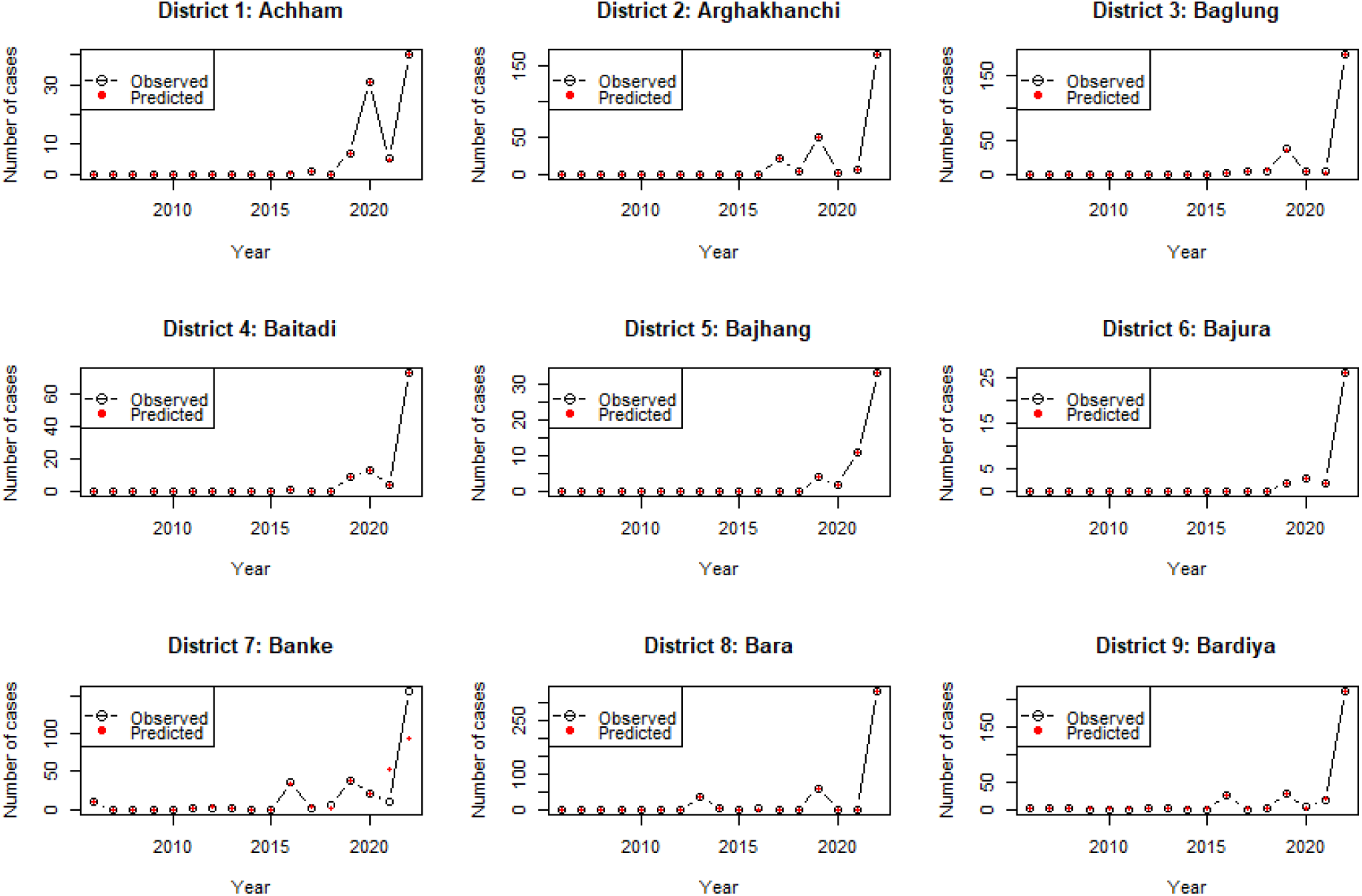
Plots of the observed vs predicted counts of dengue in districts 1 to 9

**Figure D.27:**
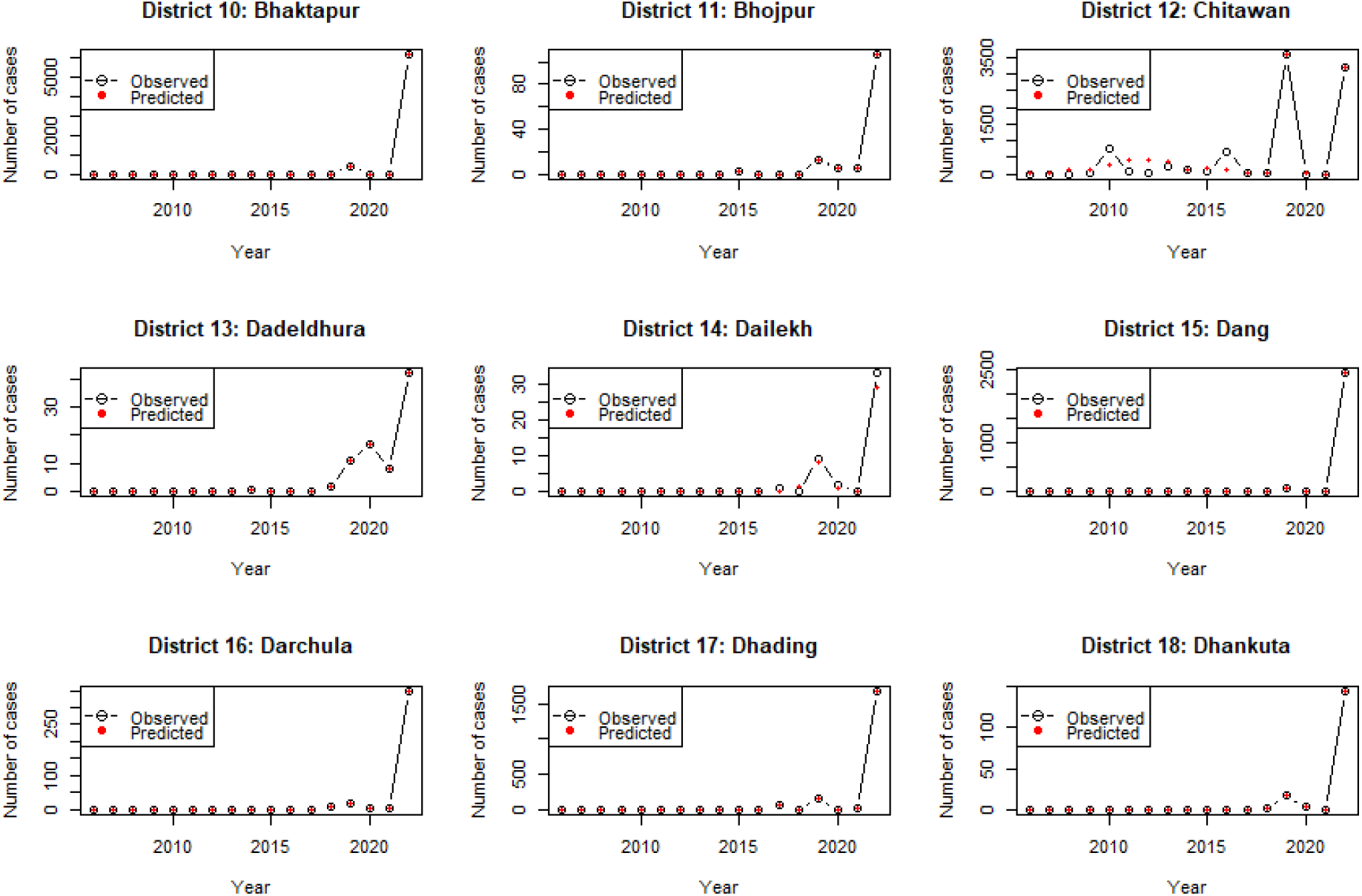
Plots of the observed vs predicted counts of dengue in districts 10 to 18

**Figure D.28:**
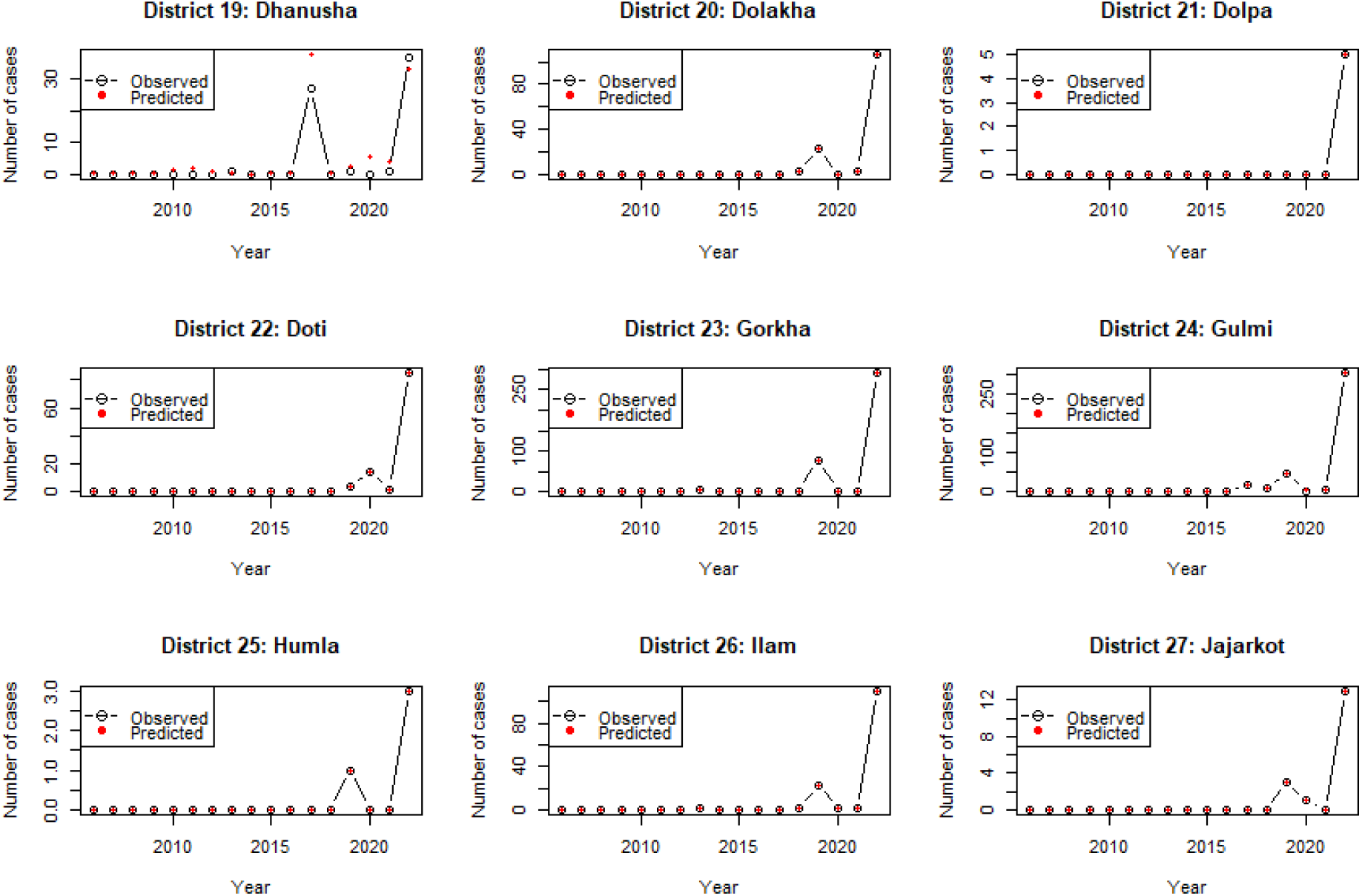
Plots of the observed vs predicted counts of dengue in districts 19 to 27

**Figure D.29:**
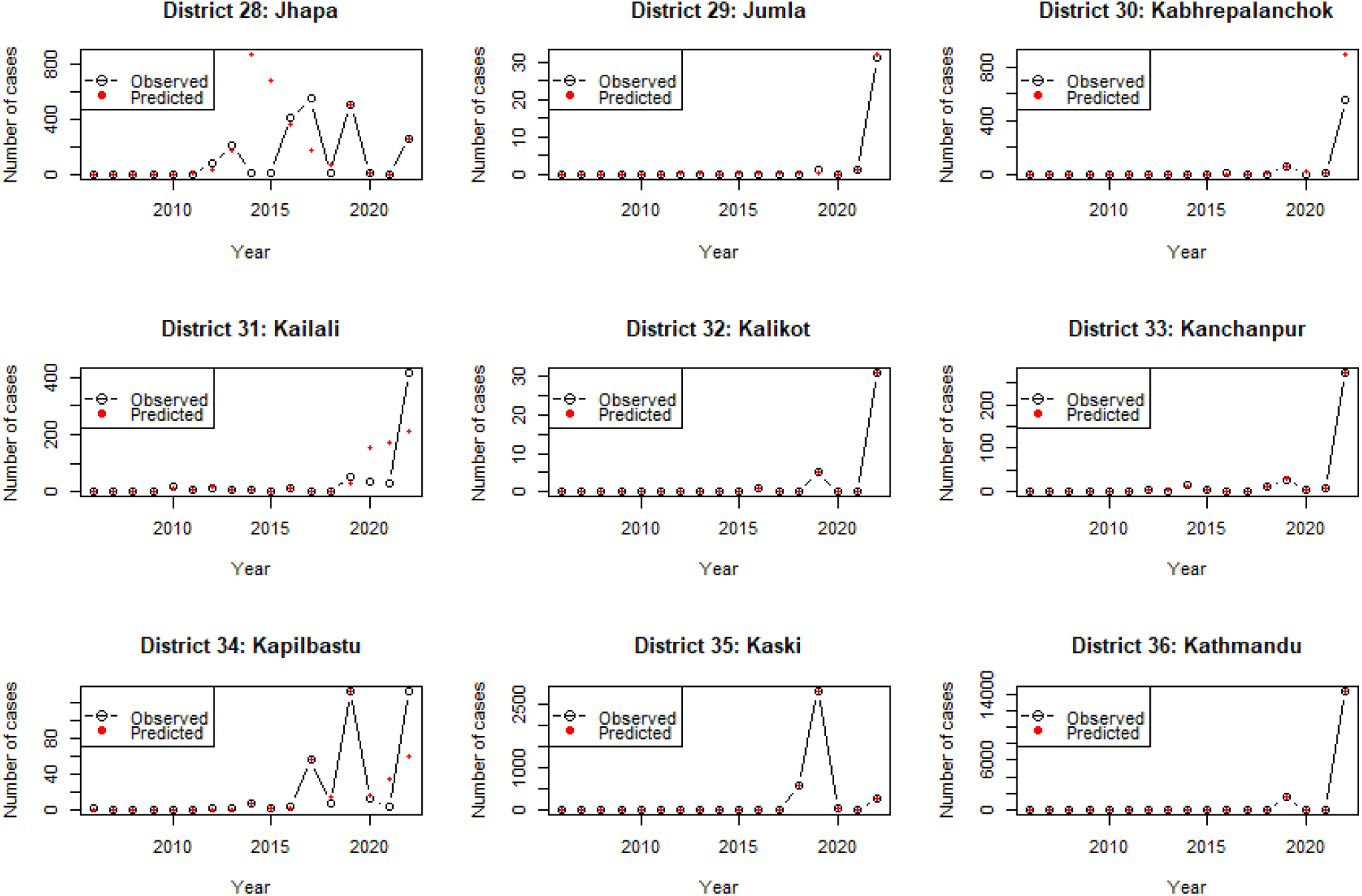
Plots of the observed vs predicted counts of dengue in districts 28 to 36

**Figure D.30:**
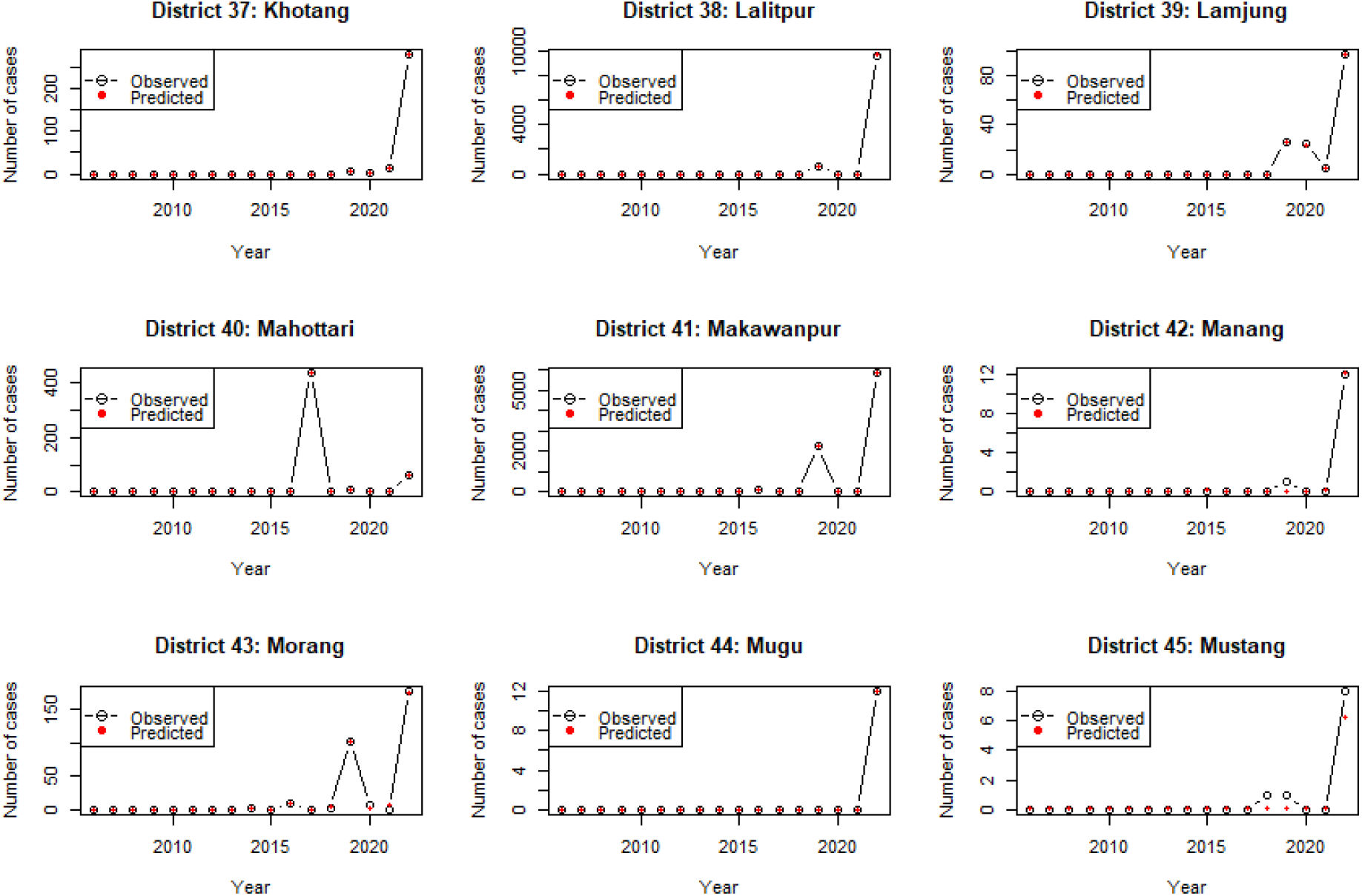
Plots of the observed vs predicted counts of dengue in districts 37 to 45

**Figure D.31:**
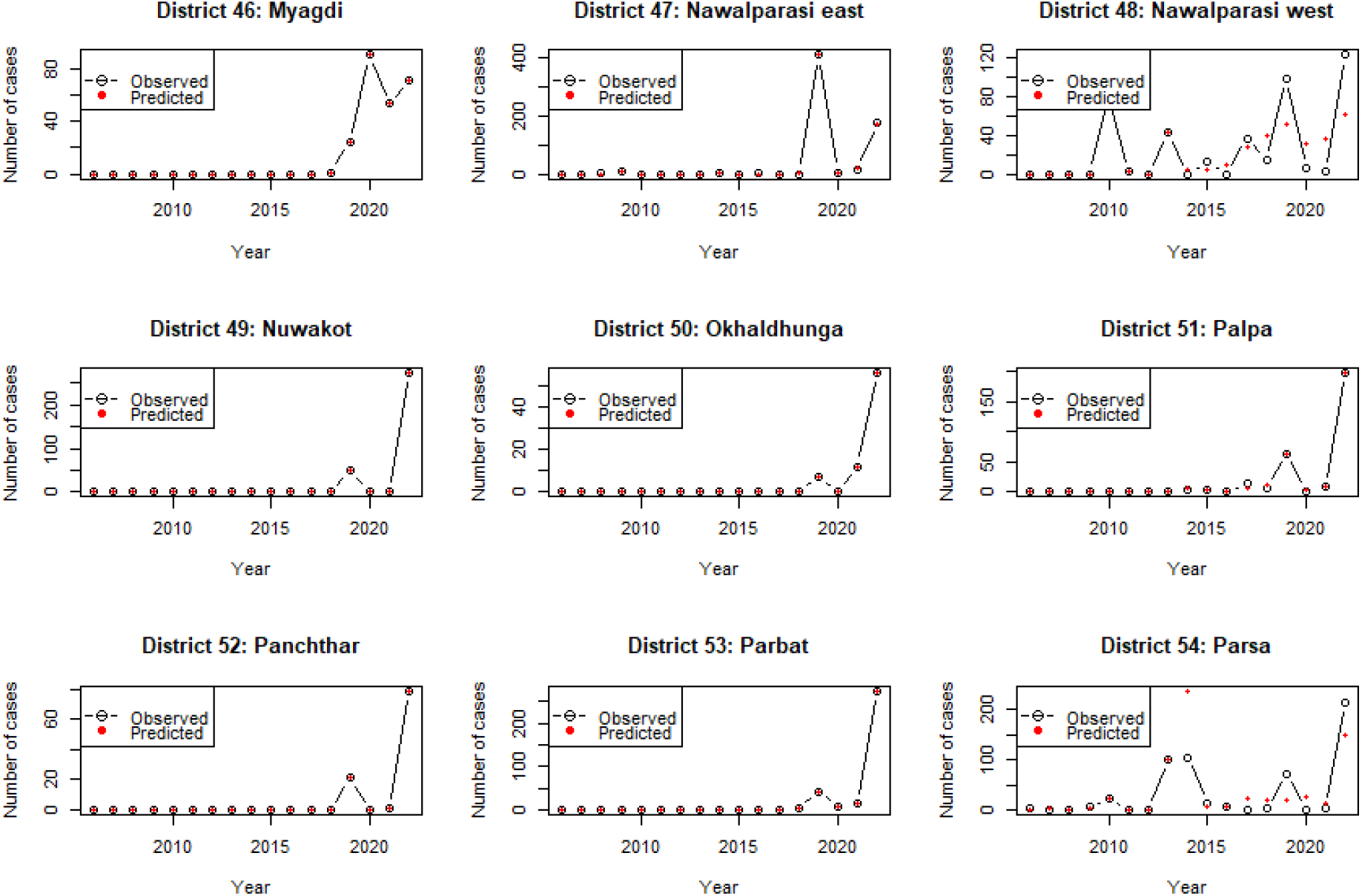
Plots of the observed vs predicted counts of dengue in districts 46 to 54

**Figure D.32:**
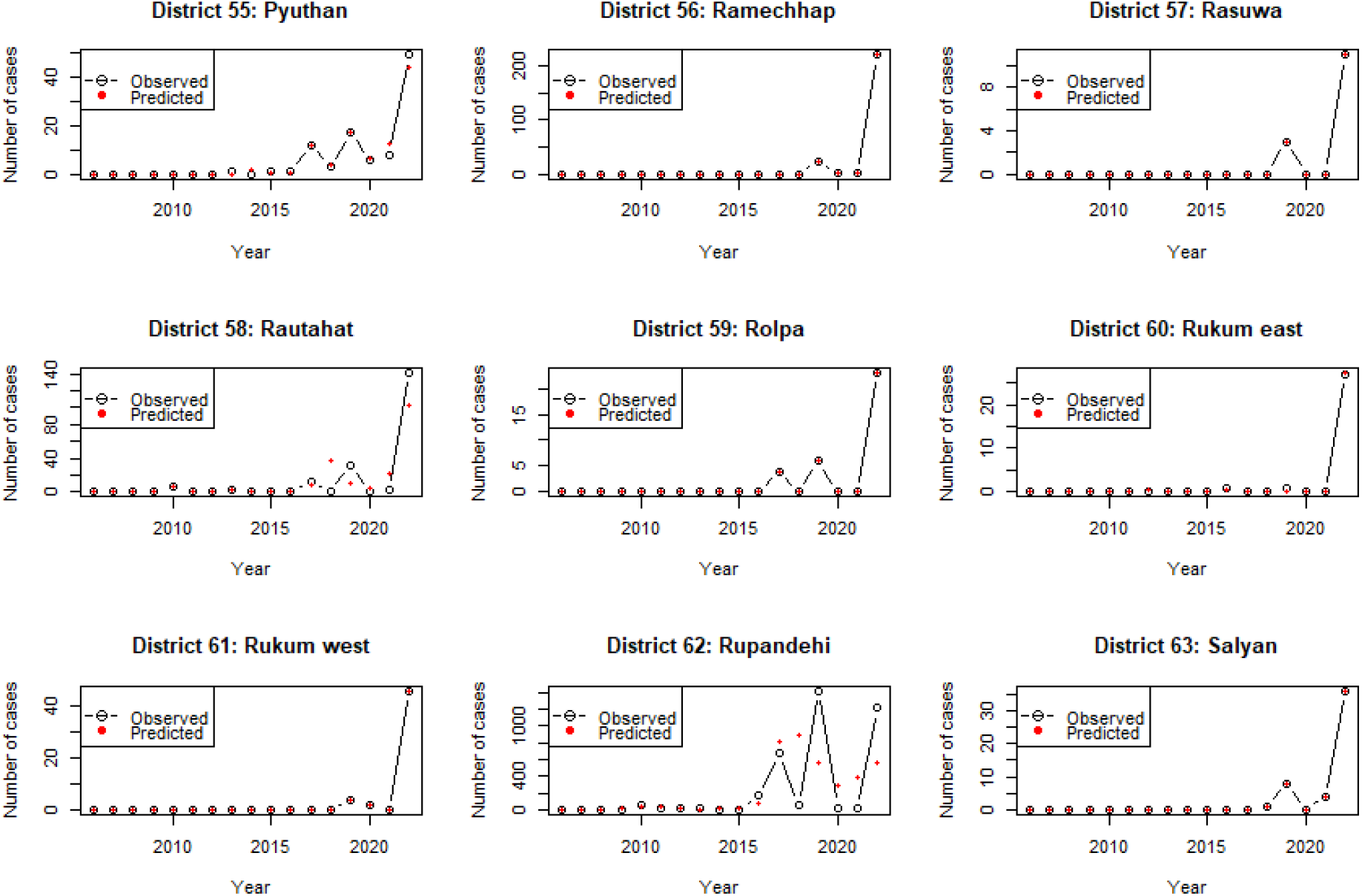
Plots of the observed vs predicted counts of dengue in districts 55 to 63

**Figure D.33:**
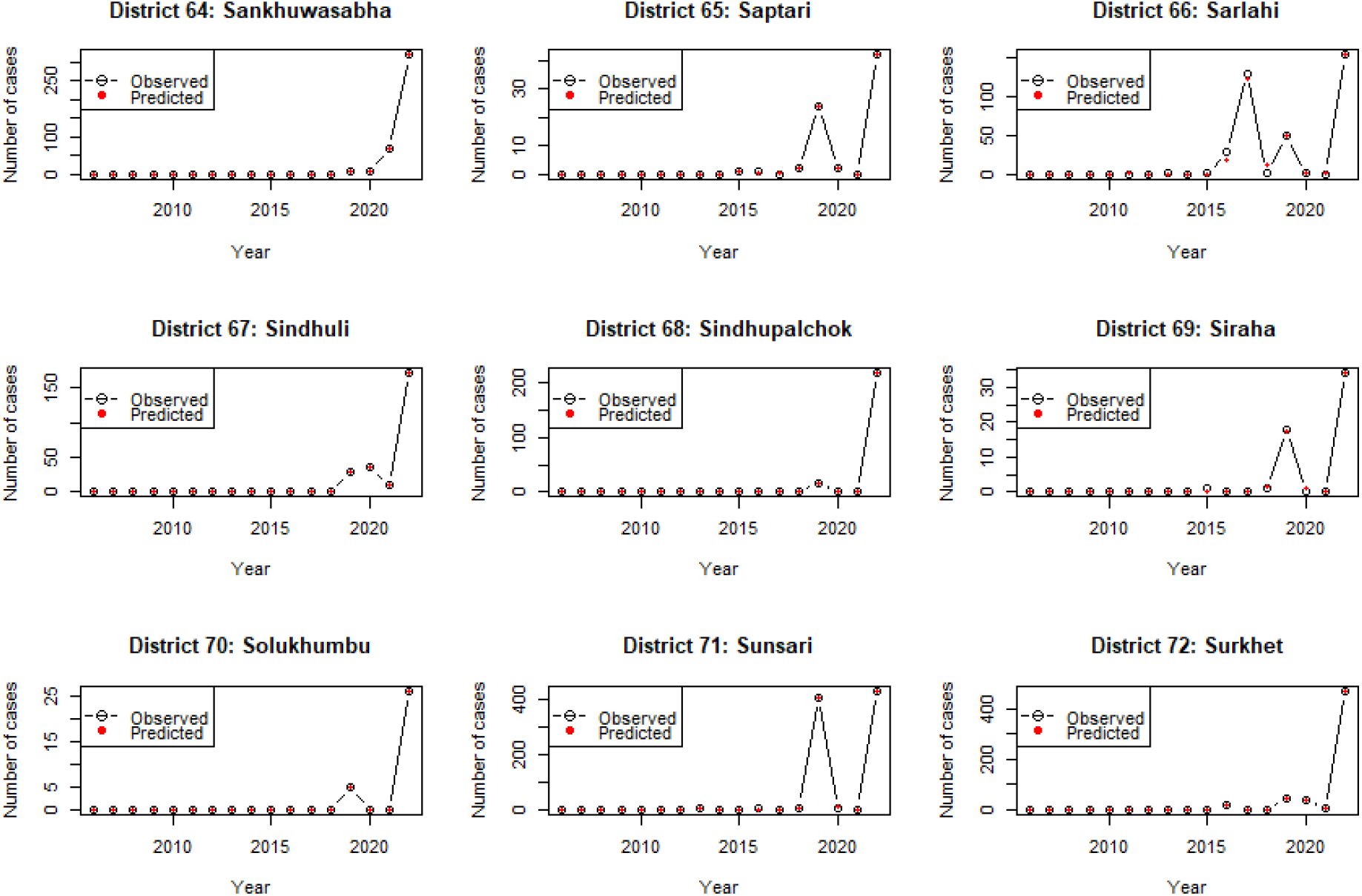
Plots of the observed vs predicted counts of dengue in districts 64 to 72

**Figure D.34:**
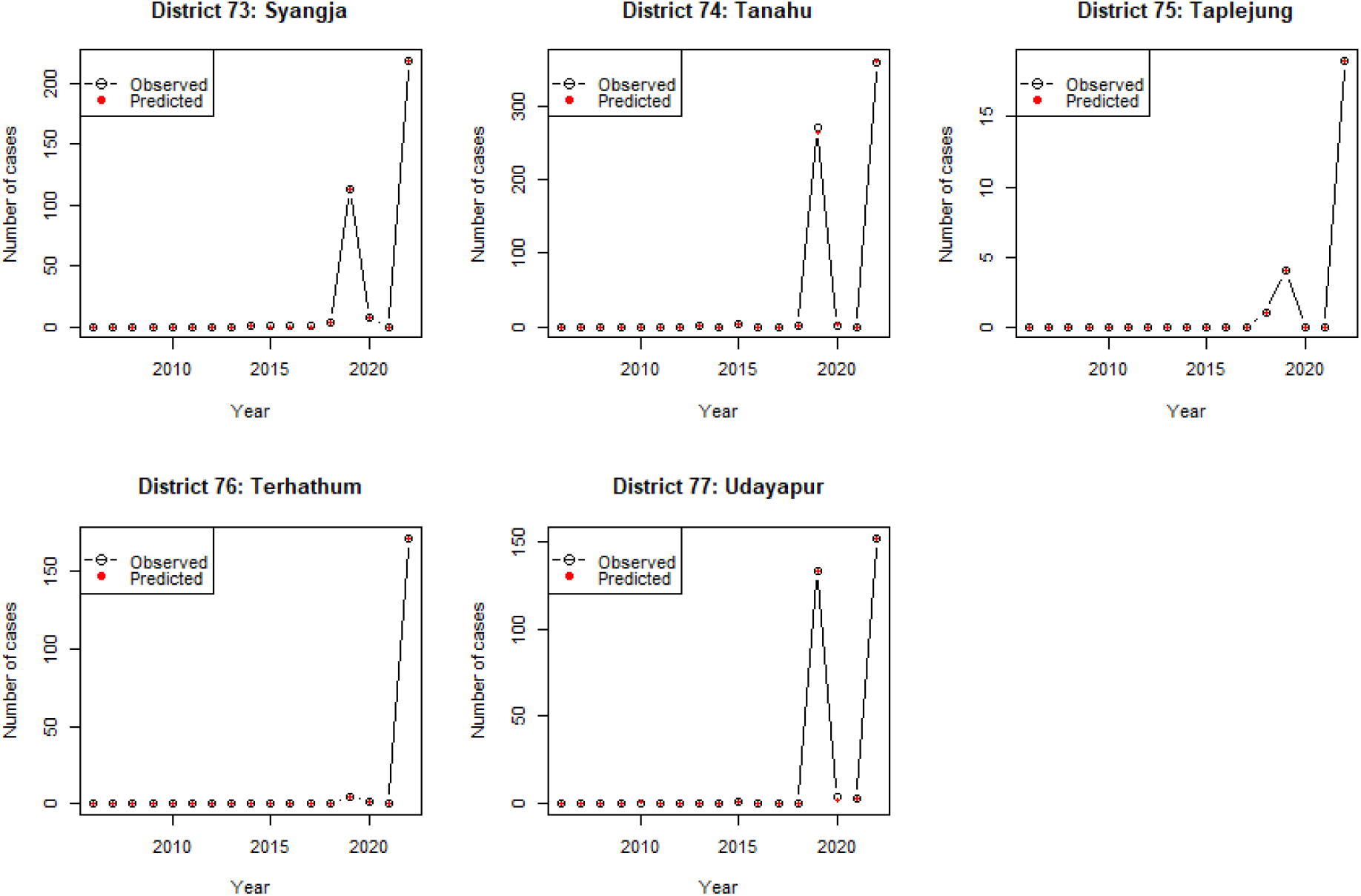
Plots of the observed vs predicted counts of dengue in districts 73 to 77

The figures below show that, in districts where model validation was unsatisfactory for a 3 OIF model, the differences between the observed and predicted dengue counts decreased as the number of OIFs in the models increased.

**Figure D.35:**
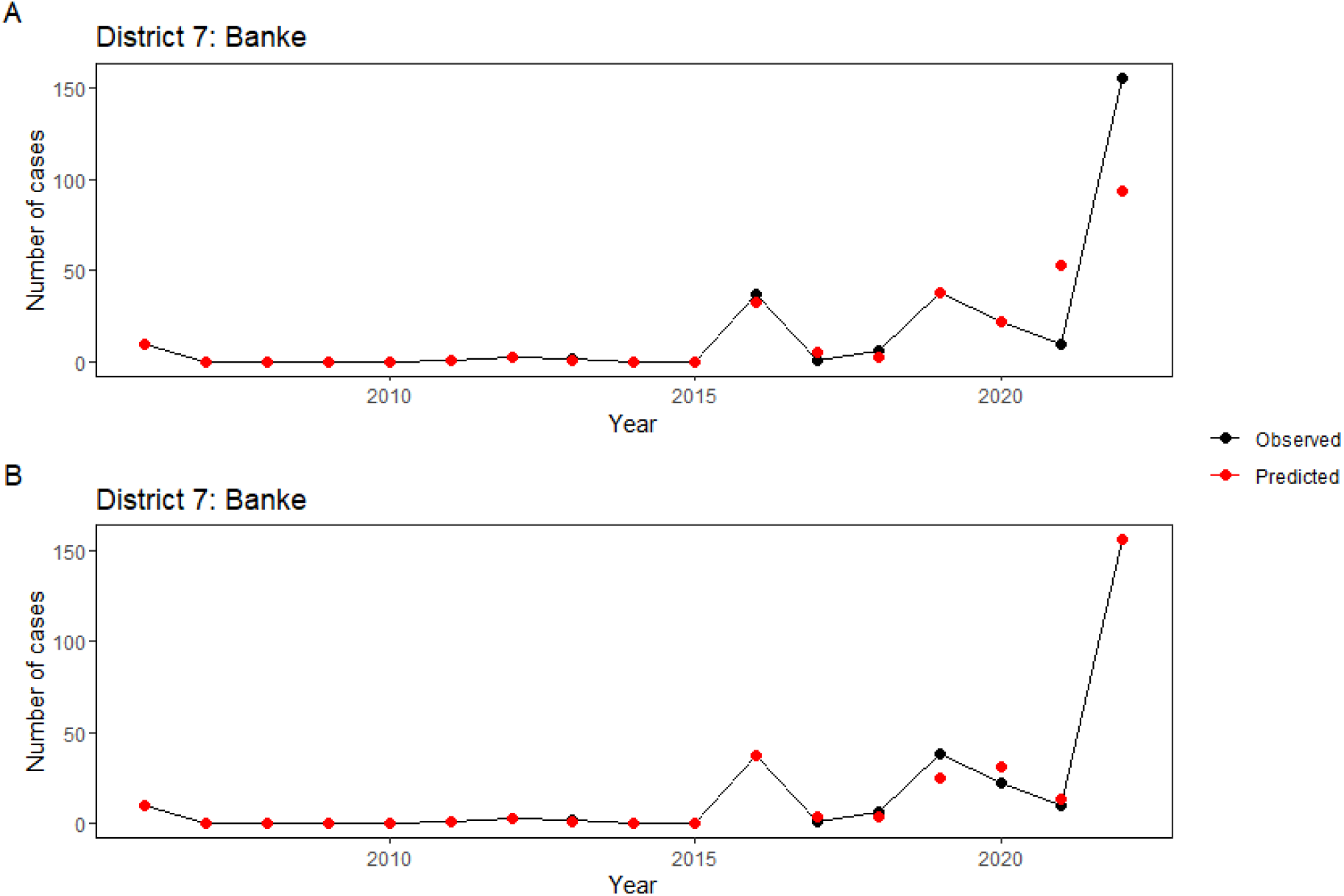
Plot of observed versus predicted dengue counts for district number 7 (Banke), illustrating models with three OIFs (Figure A) and four OIFs (Figure B).

**Figure D.36:**
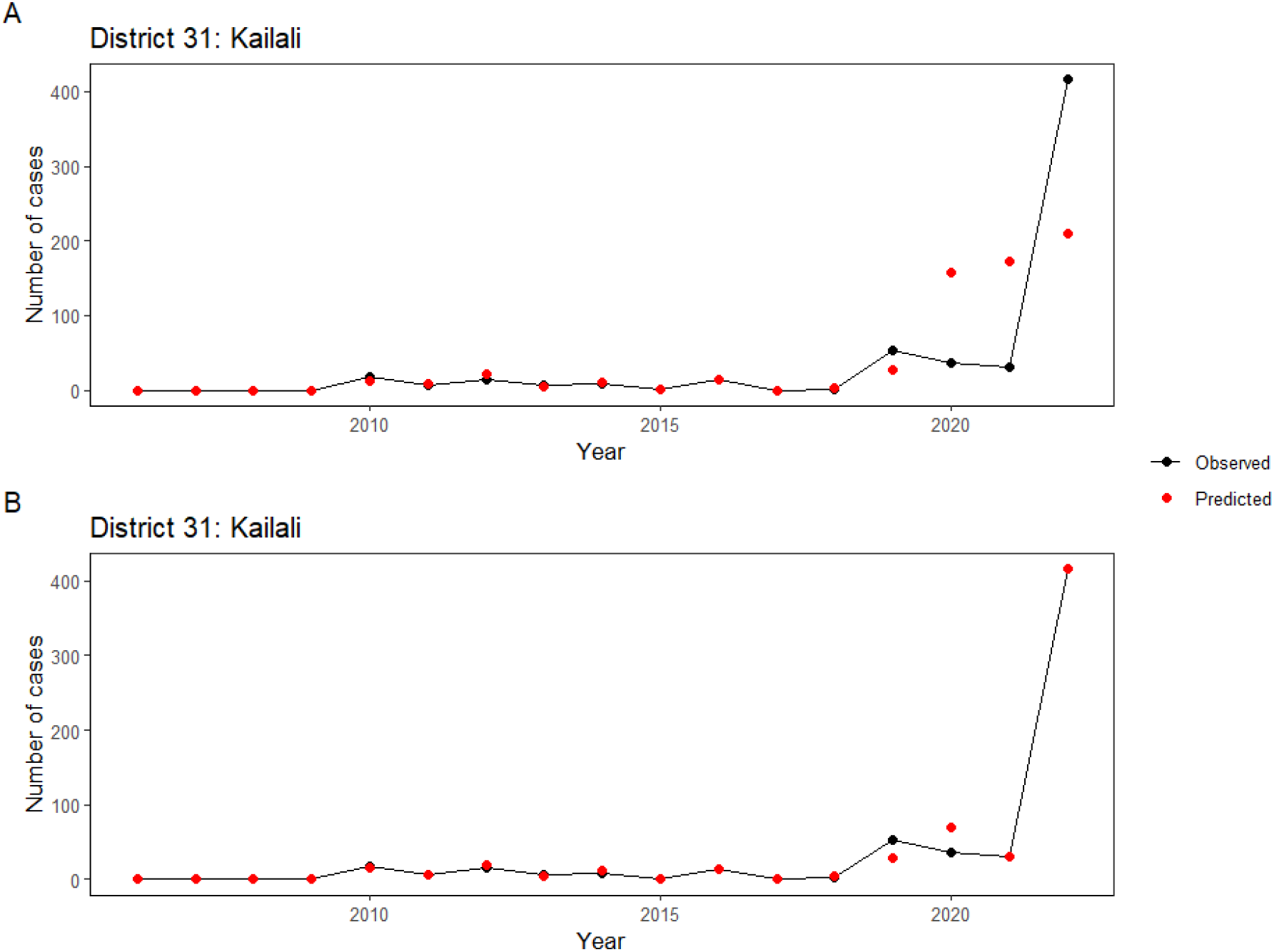
Plot of observed versus predicted dengue counts for district number 31 (Kailali), illustrating models with three OIFs (Figure A) and four OIFs (Figure B).

**Figure D.37:**
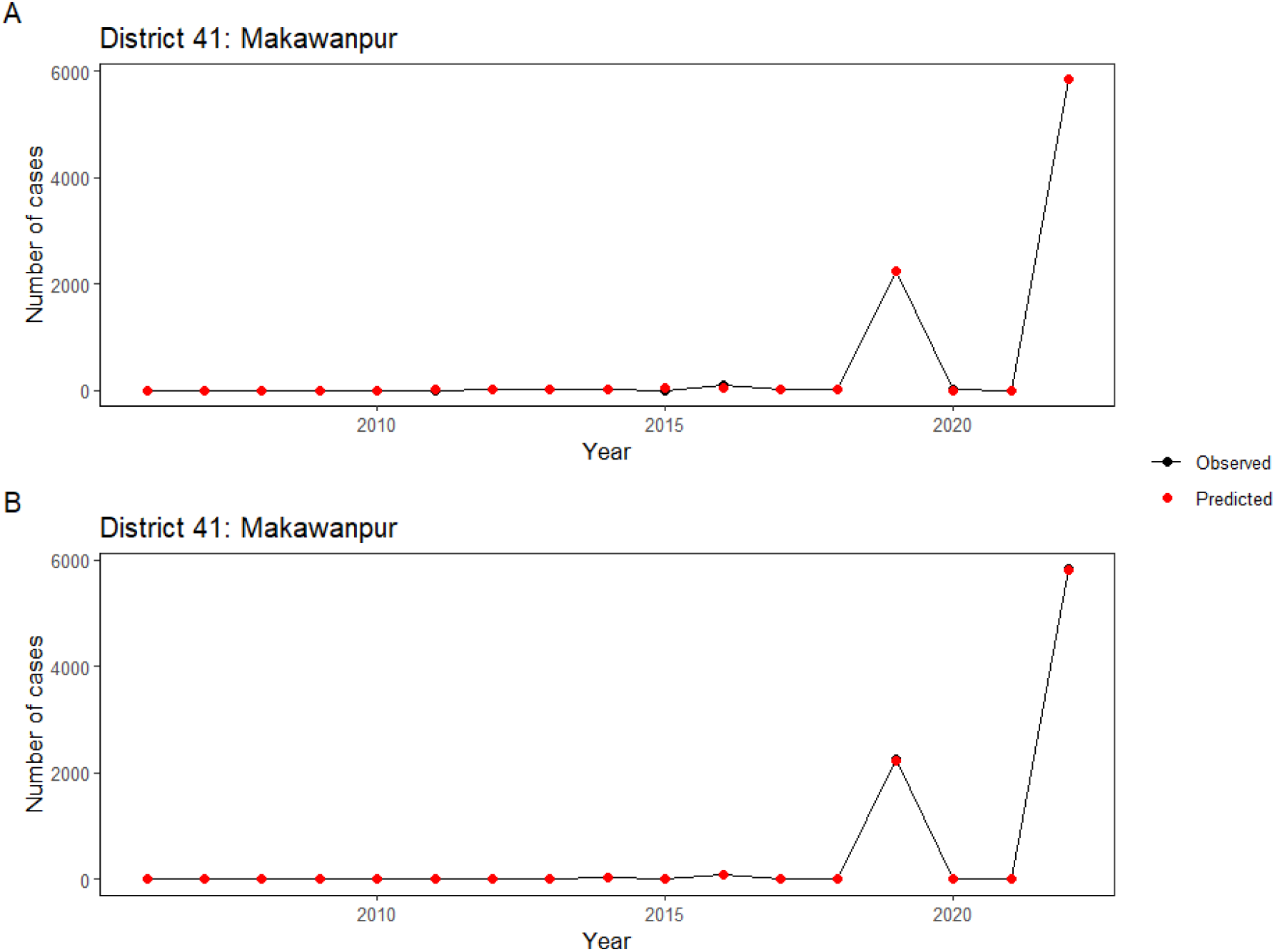
Plot of observed versus predicted dengue counts for district number 41 (Makawanpur), illustrating models with three OIFs (Figure A), and four OIFs (Figure B).

**Figure D.38:**
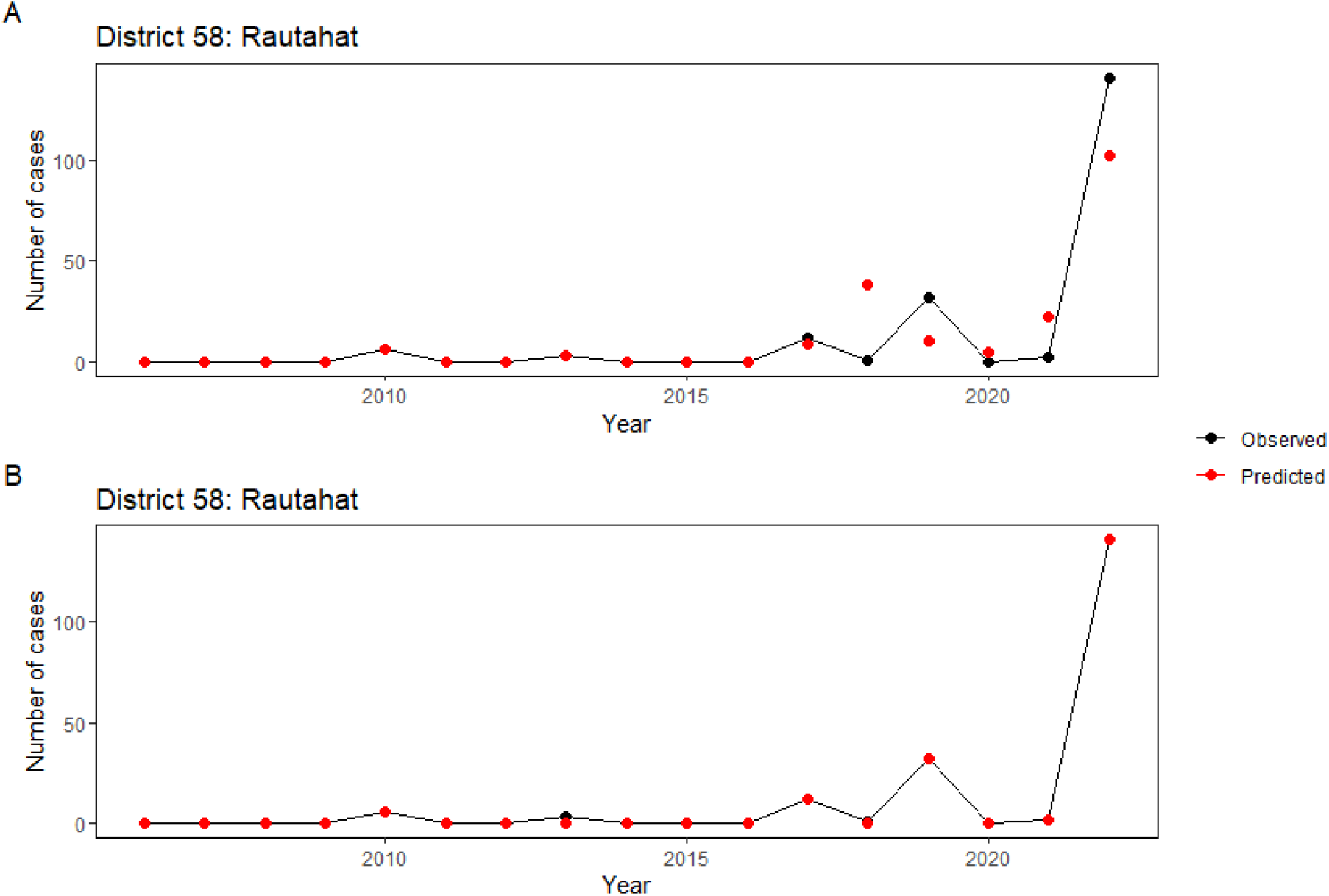
Plot of observed versus predicted dengue counts for district number 58 (Rautahat), illustrating models with three OIFs (Figure A), and four OIFs (Figure B).

**Figure D.39:**
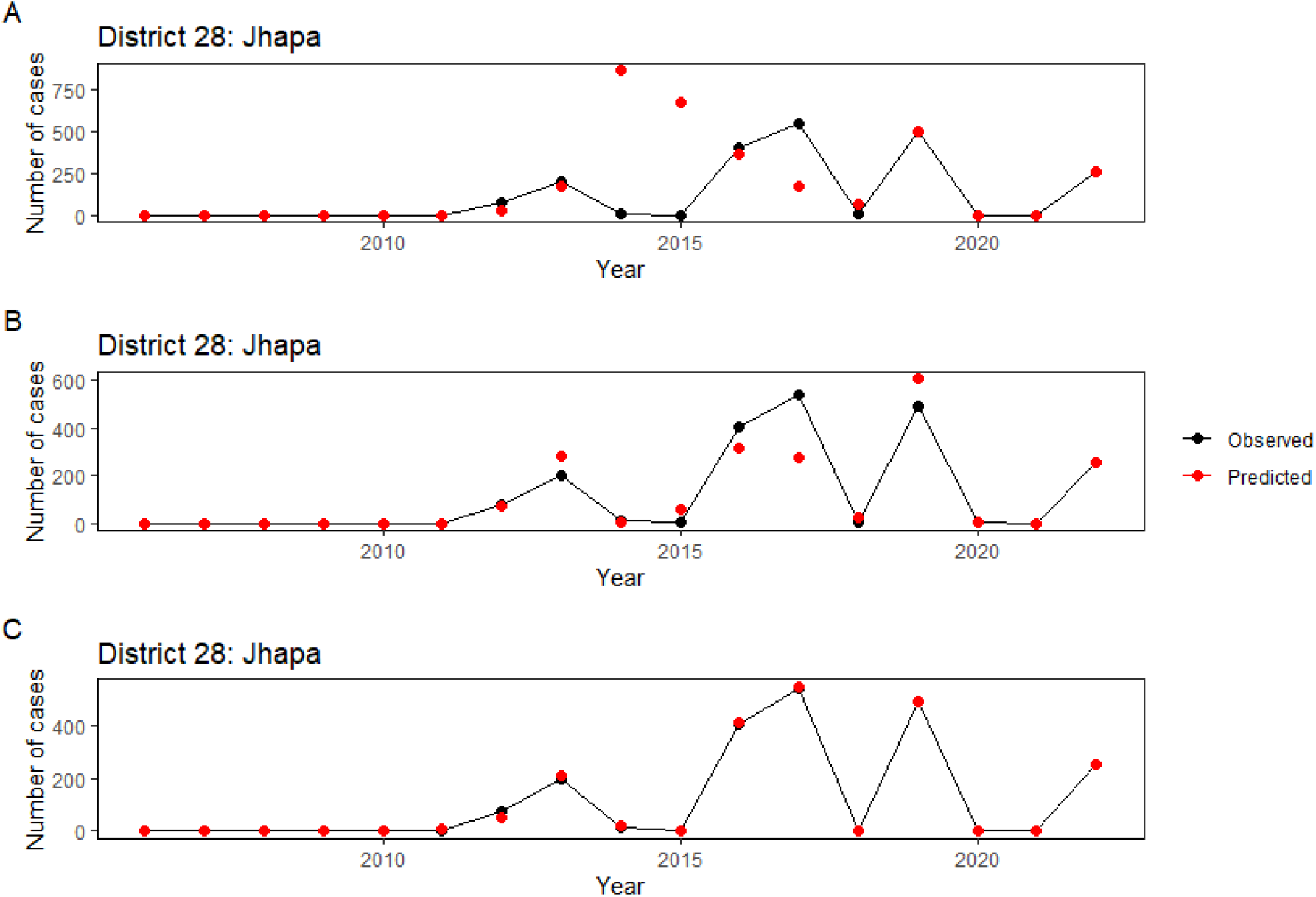
Plot of observed versus predicted dengue counts for district number 28 (Jhapa), illustrating models with three OIFs (Figure A), and four OIFs (Figure B).

**Figure D.40:**
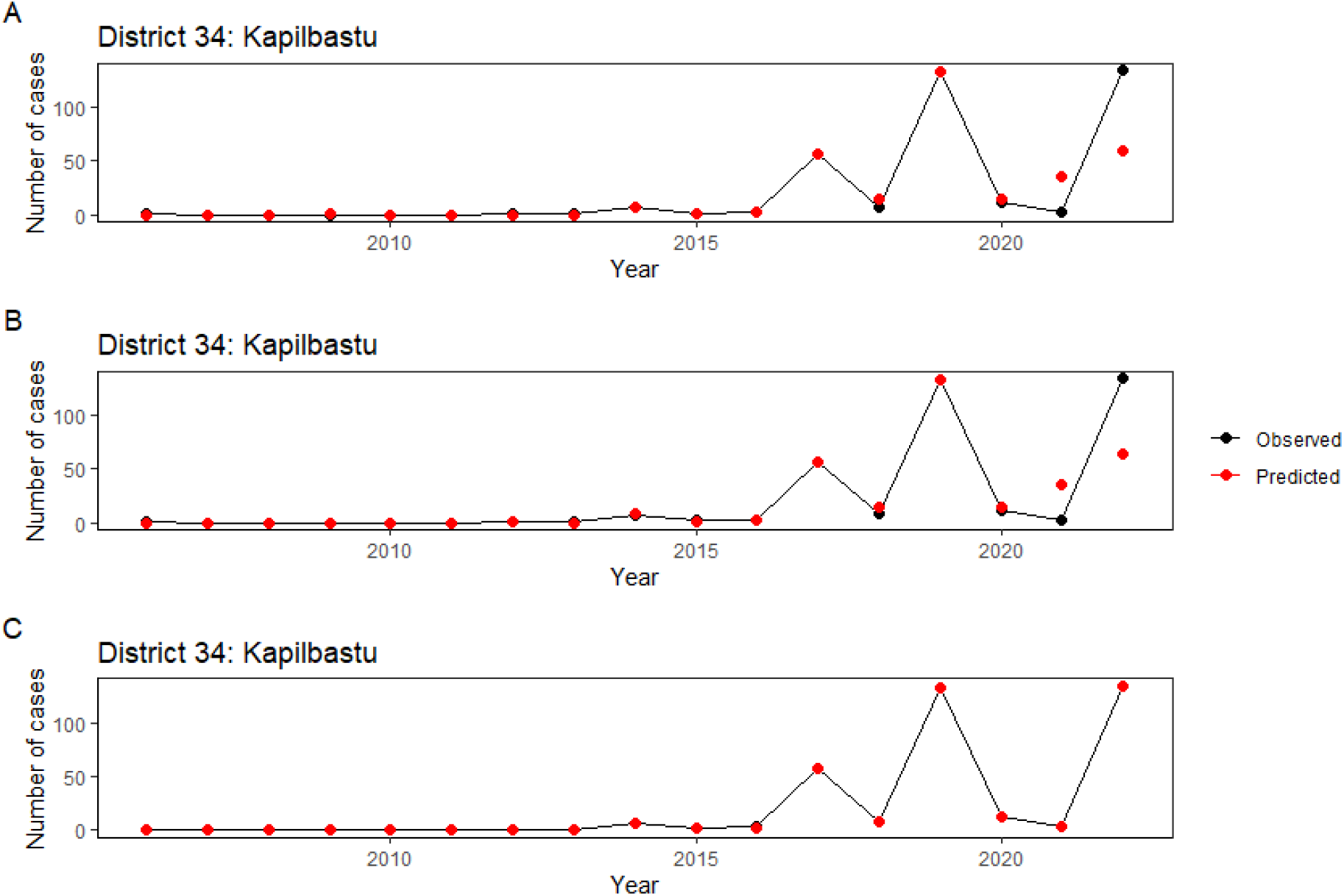
Plot of observed versus predicted dengue counts for district number 34 (Kapilbatsu), illustrating models with three OIFs (Figure A), four OIFs (Figure B), and five OIFs (Figure C).

**Figure D.41:**
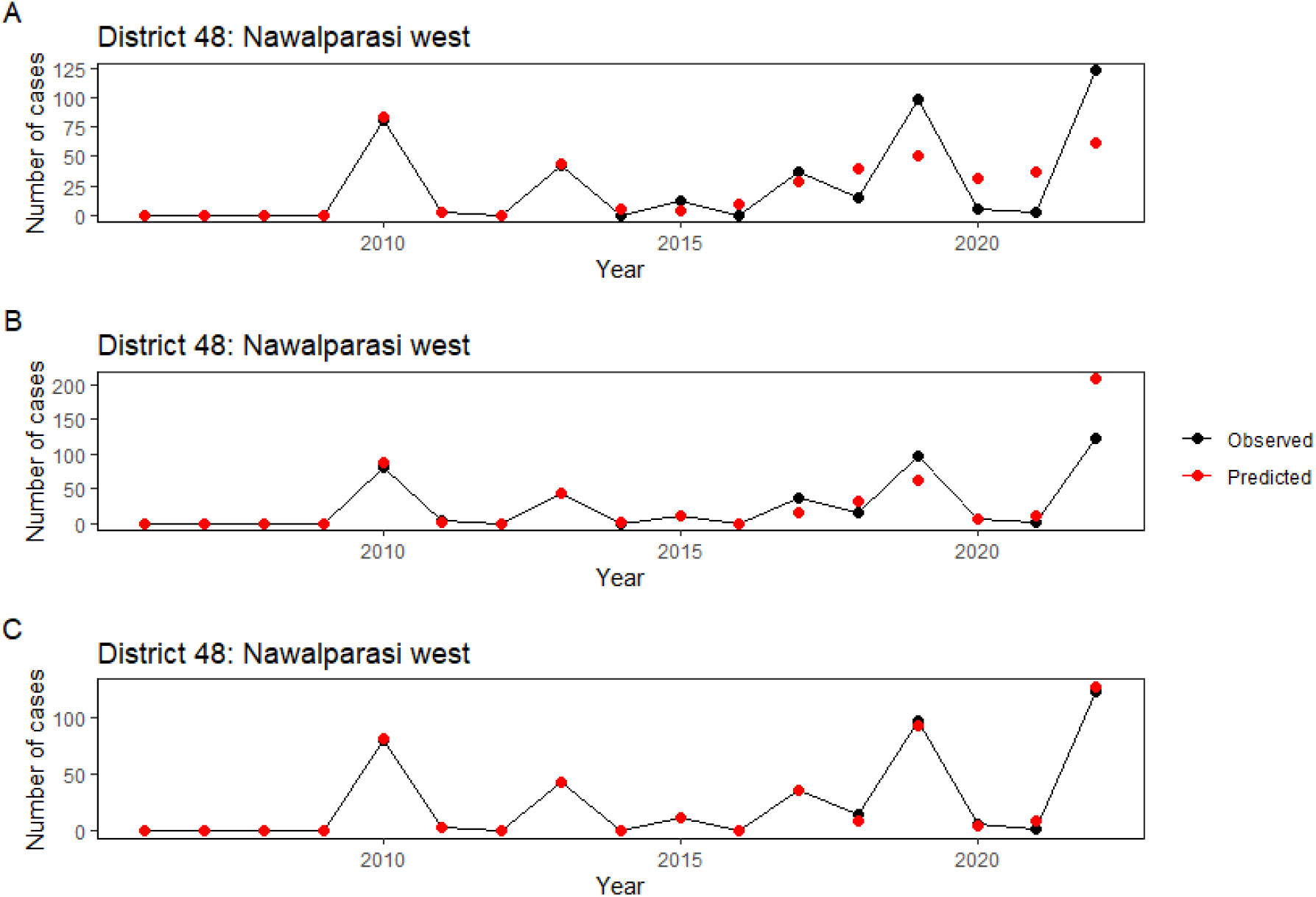
Plot of observed versus predicted dengue counts for district number 48 (Nawalparasi West), illustrating models with three OIFs (Figure A), four OIFs (Figure B), and five OIFs (Figure C).

**Figure D.42:**
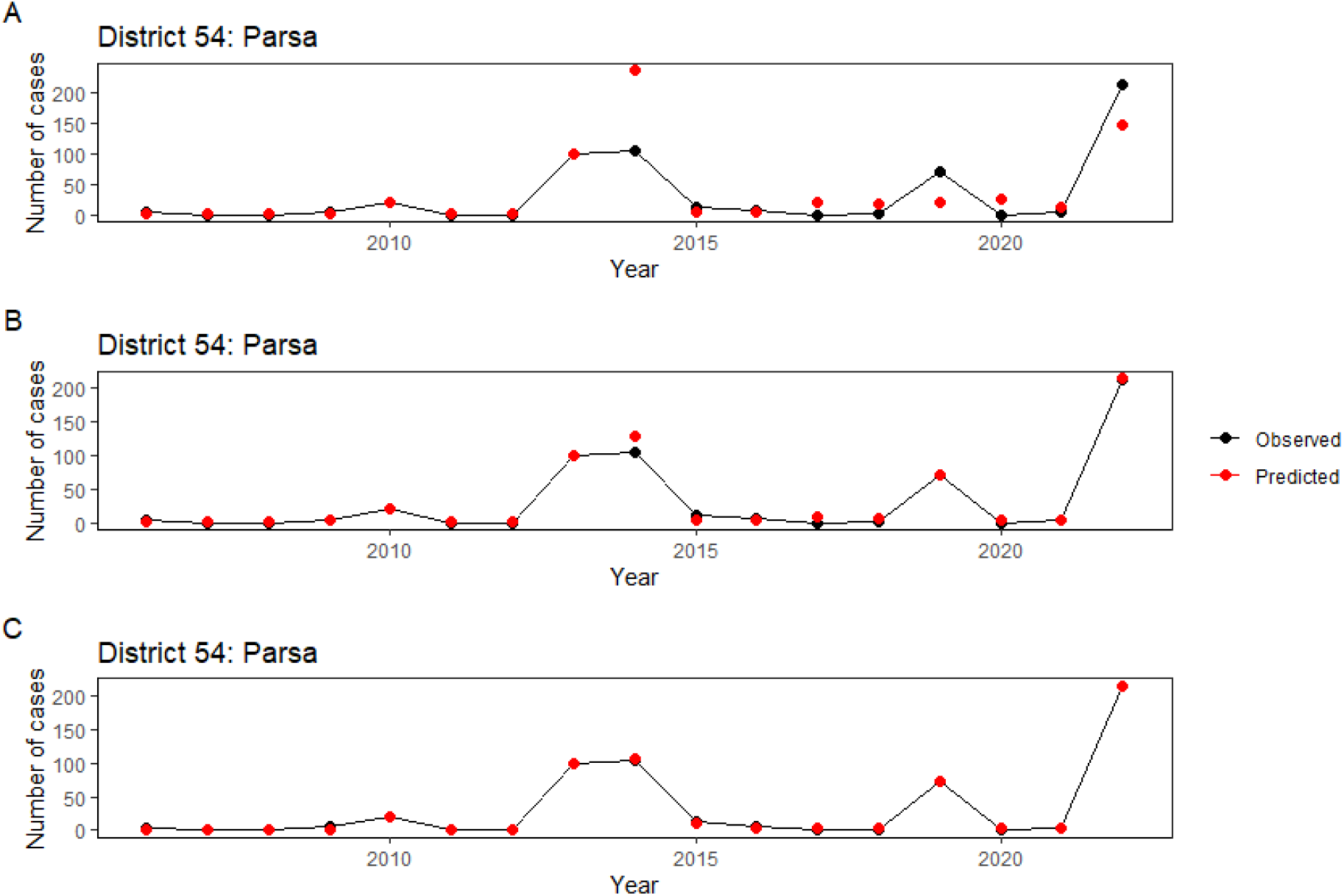
Plot of observed versus predicted dengue counts for district number 54 (Parsa), illustrating models with three OIFs (Figure A), four OIFs (Figure B), and five OIFs (Figure C).

**Figure D.43:**
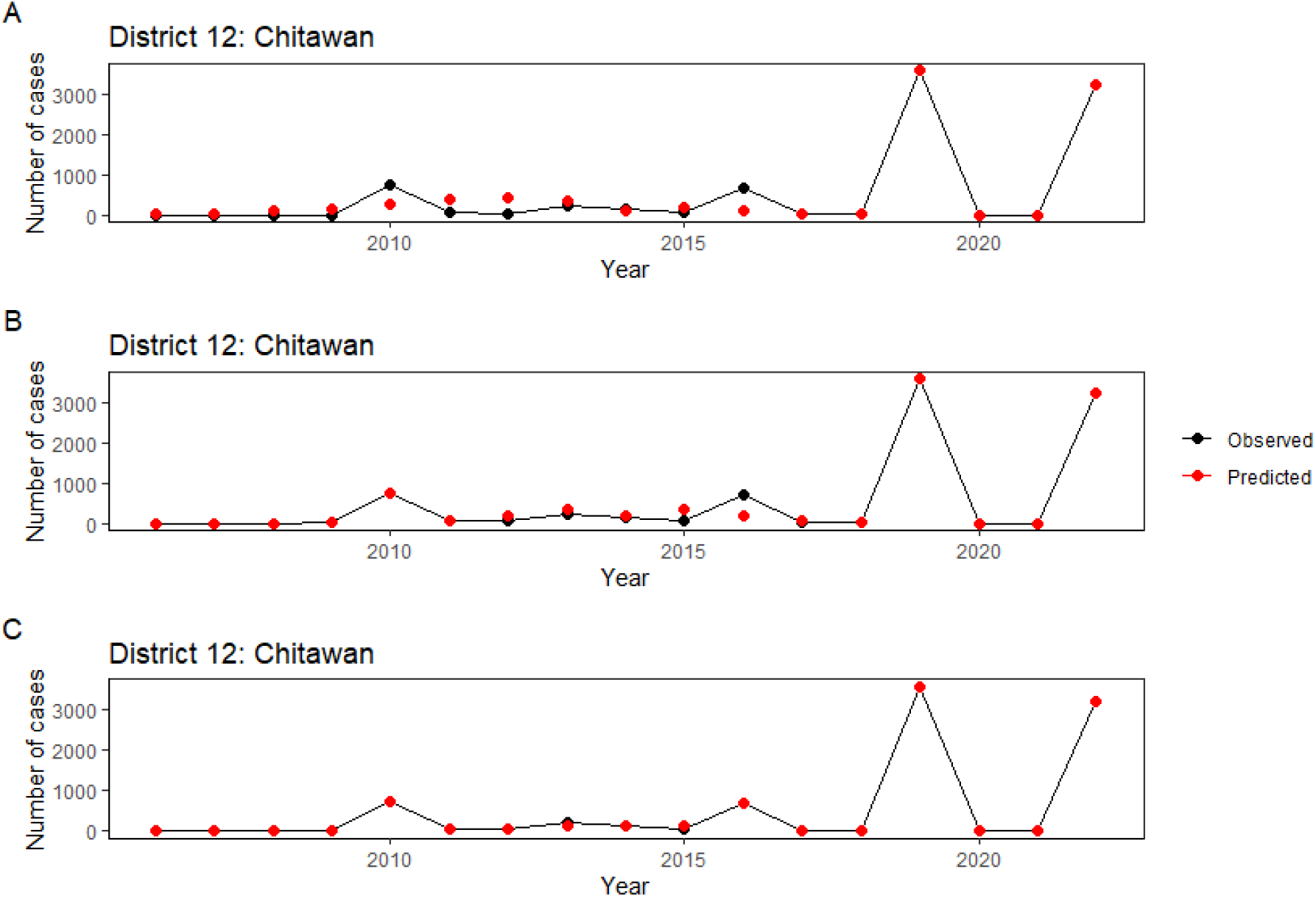
Plot of observed versus predicted dengue counts for district number 12 (Chitawan), illustrating models with three OIFs (Figure A), four OIFs (Figure B), and five OIFs (Figure C).

**Figure D.44:**
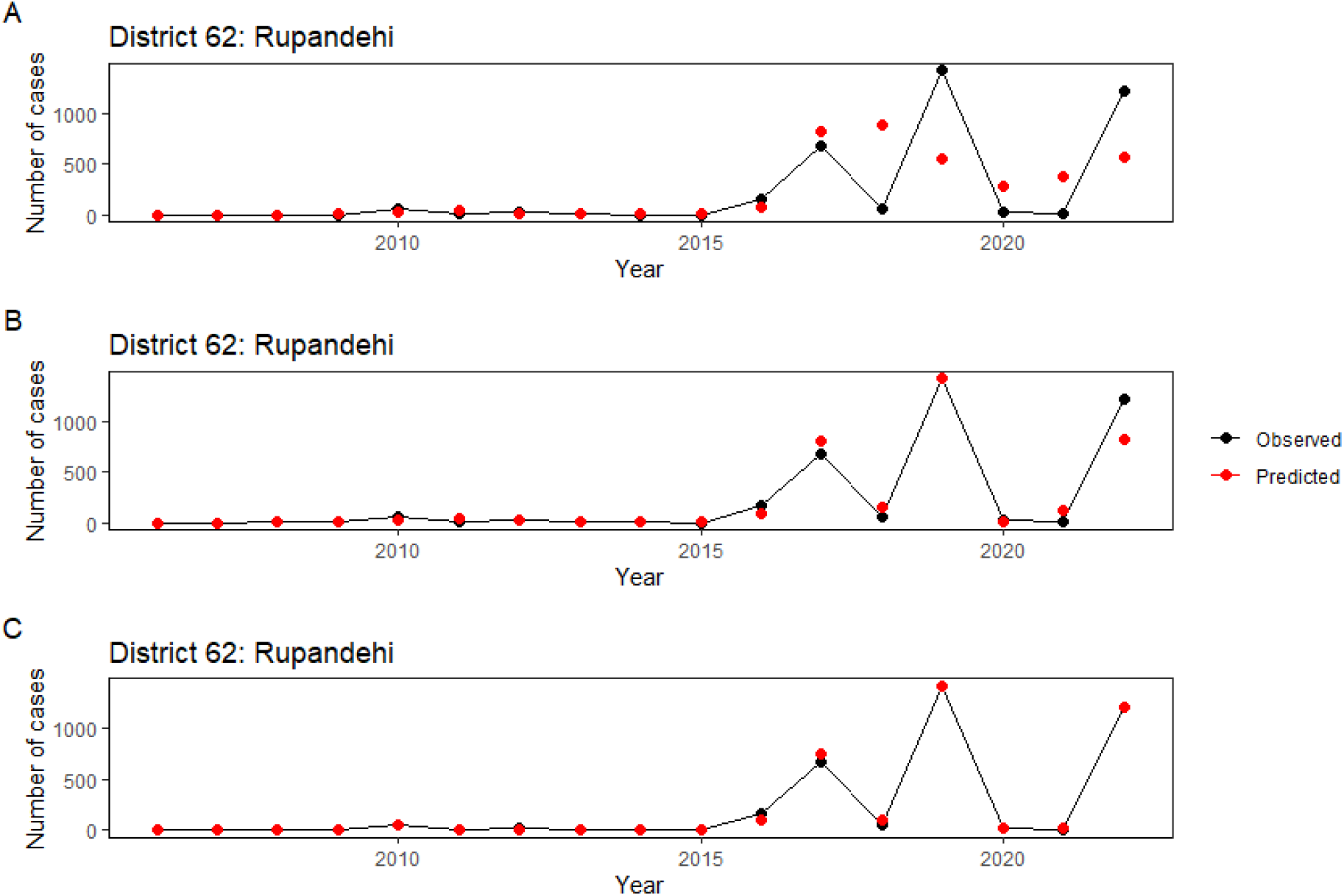
Plot of observed versus predicted dengue counts for district number 62 (Rupandehi), illustrating models with three OIFs (Figure A), four OIFs (Figure B), and five OIFs (Figure C).

## References

Abatzoglou, J.T., Dobrowski, S.Z., Parks, S.A., Hegewisch, K.C., 2018. Terraclimate, a high-resolution global dataset of monthly climate and climatic water balance from 1958–2015. Scientific data 5, 1–12.

Abdulsalam, F.I., Antunez, P., Yimthiang, S., Jawjit, W., 2022. Influence of climate variables on dengue fever occurrence in the southern region of thailand. PLOS Global Public Health 2, e0000188.

Acharya, B.K., Cao, C., Lakes, T., Chen, W., Naeem, S., 2016. Spatiotemporal analysis of dengue fever in nepal from 2010 to 2014. BMC Public Health 16, 1–10.

Acharya, B.K., Cao, C., Xu, M., Khanal, L., Naeem, S., Pandit, S., 2018. Present and future of dengue fever in nepal: mapping climatic suitability by ecological niche model. International journal of environmental research and public health 15, 187.

Acharya, B.K., Khanal, L., Dhimal, M., 2025. Increased thermal suitability elevates the risk of dengue transmission across the mid hills of nepal. PLoS One 20, e0322031.

Acharya, K.P., Chaulagain, B., Acharya, N., Shrestha, K., Subramanya, S.H., 2020. Establishment and recent surge in spatio-temporal spread of dengue in nepal. Emerging Microbes & Infections 9, 676–679.

Anderson, C., Lee, D., Dean, N., 2014. Identifying clusters in bayesian disease mapping. Biostatistics 15, 457–469.

Azzalini, A., 1985. A class of distributions which includes the normal ones. Scandinavian Journal of Statistics 12, 171–178.

Besag, J., York, J., Mollié, A., 1991. Bayesian image restoration, with two applications in spatial statistics. Annals of the Institute of Statistical Mathematics 43, 1–20.

Bhatt, S., Gething, P.W., Brady, O.J., Messina, J.P., Farlow, A.W., Moyes, C.L., Drake, J.M., Brownstein, J.S., Hoen, A.G., Sankoh, O., et al., 2013. The global distribution and burden of dengue. Nature 496, 504–507.

Bhuju, U.R., Shakya, P.R., Basnet, T.B., Shrestha, S., 2007. Nepal biodiversity resource book: protected areas, Ramsar sites, and World Heritage sites.

Campbell, K.M., Lin, C., Iamsirithaworn, S., Scott, T.W., 2013. The complex relationship between weather and dengue virus transmission in thailand. The American journal of tropical medicine and hygiene 89, 1066.

do Carmo, R.F., Silva Júnior, J.V.J., Pastor, A.F., de Souza, C.D.F., 2020. Spatiotemporal dynamics, risk areas and social determinants of dengue in northeastern brazil, 2014–2017: an ecological study. Infectious diseases of poverty 9, 1–16.

Chen, Y., Liu, T., Yu, X., Zeng, Q., Cai, Z., Wu, H., Zhang, Q., Xiao, J., Ma, W., Pei, S., et al., 2022. An ensemble forecast system for tracking dynamics of dengue outbreaks and its validation in china. PLoS computational biology 18, e1010218.

Colón-González, F.J., Sewe, M.O., Tompkins, A.M., Sjödin, H., Casallas, A., Rocklöv, J., Caminade, C., Lowe, R., 2021. Projecting the risk of mosquito-borne diseases in a warmer and more populated world: a multi-model, multi-scenario intercomparison modelling study. The Lancet Planetary Health 5, e404–e414.

Cooper, G.F., Villamarin, R., Tsui, F.C.R., Millett, N., Espino, J.U., Wagner, M.M., 2015. A method for detecting and characterizing outbreaks of infectious disease from clinical reports. Journal of biomedical informatics 53, 15–26.

Dhimal, M., Gautam, I., Kreß, A., Müller, R., Kuch, U., 2014. Spatio-temporal distribution of dengue and lymphatic filariasis vectors along an altitudinal transect in central nepal. PLoS Neglected Tropical Diseases 8, e3035.

Diggle, P.J., Giorgi, E., 2019. Model-based geostatistics for global public health: methods and applications. CRC Press.

Dumre, S.P., Bhandari, R., Shakya, G., Shrestha, S.K., Cherif, M.S., Ghimire, P., Klungthong, C., Yoon, I.K., Hirayama, K., Na-Bangchang, K., et al., 2017. Dengue virus serotypes 1 and 2 responsible for major dengue outbreaks in nepal: clinical, laboratory, and epidemiological features. The American journal of tropical medicine and hygiene 97, 1062.

Efron, B., 1987. Better bootstrap confidence intervals. Journal of the American statistical Association 82, 171–185.

Efron, B., 2000. The bootstrap and modern statistics. Journal of the American Statistical Association 95, 1293–1296.

Efron, B., Tibshirani, R.J., 1994. An introduction to the bootstrap. Chapman and Hall/CRC.

da Fonseca, B.A.L., Fonseca, S.N.S., 2002. Dengue virus infections. Current opinion in pediatrics 14, 67–71.

Franco, C., Ferreira, L.S., Sudbrack, V., Borges, M.E., Poloni, S., Prado, P.I., White, L.J., Águas, R., Kraenkel, R.A., Coutinho, R.M., 2022. Percolation across households in mechanistic models of non-pharmaceutical interventions in sars-cov-2 disease dynamics. Epidemics 39, 100551.

Gathsaurie, Neelika Malavige, P., Sjö, K., Singh, J.M., Piedagnel, C., Mowbray, S., Estani, Steven Chee Loon Lim, A.M., Siquierra, Graham S. and Ogg, L., Fraisse, I., Ribeiro, 2023. Facing the escalating burden of dengue: Challenges and perspectives. PLos Global Health 3, e0002598.

Giorgi, E., Fronterrè, C., Macharia, P.M., Alegana, V.A., Snow, R.W., Diggle, P.J., 2021. Model building and assessment of the impact of covariates for disease prevalence mapping in low-resource settings: to explain and to predict. Journal of The Royal Society Interface 18, 20210104.

Gupta, B.P., Haselbeck, A., Kim, J.H., Marks, F., Saluja, T., 2018. The dengue virus in nepal: gaps in diagnosis and surveillance. Annals of clinical microbiology and antimicrobials 17, 1–5.

Hjelm, L., Mathiassen, A., Miller, D., Wadhwa, A., 2017. Creation of a wealth index. United Nations World Food Programme .

Howe, L.D., Galobardes, B., Matijasevich, A., Gordon, D., Johnston, D., Onwujekwe, O., Patel, R., Webb, E.A., Lawlor, D.A., Hargreaves, J.R., 2012. Measuring socio-economic position for epidemiological studies in low-and middle-income countries: a methods of measurement in epidemiology paper. International journal of epidemiology 41, 871–886.

Imai, N., Dorigatti, I., Cauchemez, S., Ferguson, N.M., 2015. Estimating dengue transmission intensity from sero-prevalence surveys in multiple countries. PLoS neglected tropical diseases 9, e0003719.

Kolimenakis, A., Heinz, S., Wilson, M.L., Winkler, V., Yakob, L., Michaelakis, A., Papachristos, D., Richardson, C., Horstick, O., 2021. The role of urbanisation in the spread of aedes mosquitoes and the diseases they transmit—a systematic review. PLoS neglected tropical diseases 15, e0009631.

Kong, L., Xu, C., Mu, P., Li, J., Qiu, S., Wu, H., 2019. Risk factors spatial-temporal detection for dengue fever in guangzhou. Epidemiology & Infection 147.

Li, C., Lim, T., Han, L., Fang, R., 1985. Rainfall, abundance of aedes aegypti and dengue infection in selangor, malaysia. The Southeast Asian journal of tropical medicine and public health 16, 560–568.

Li, C., Liu, Z., Li, W., Lin, Y., Hou, L., Niu, S., Xing, Y., Huang, J., Chen, Y., Zhang, S., et al., 2023. Projecting future risk of dengue related to hydrometeorological conditions in mainland china under climate change scenarios: a modelling study. The Lancet Planetary Health 7, e397–e406.

Li, C., Wu, X., Sheridan, S., Lee, J., Wang, X., Yin, J., Han, J., 2021. Interaction of climate and socio-ecological environment drives the dengue outbreak in epidemic region of china. PLoS Neglected Tropical Diseases 15, e0009761.

Lu, J., Meyer, S., 2023. A zero-inflated endemic–epidemic model with an application to measles time series in germany. Biometrical Journal 65, 2100408.

Man, O., Kraay, A., Thomas, R., Trostle, J., Lee, G.O., Robbins, C., Morrison, A.C., Coloma, J., Eisenberg, J.N., 2023. Characterizing dengue transmission in rural areas: A systematic review. PLOS Neglected Tropical Diseases 17, e0011333.

Messina, J.P., Brady, O.J., Golding, N., Kraemer, M.U., Wint, G.W., Ray, S.E., Pigott, D.M., Shearer, F.M., Johnson, K., Earl, L., et al., 2019. The current and future global distribution and population at risk of dengue. Nature microbiology 4, 1508–1515.

Meyer, S., Held, L., Höhle, M., 2016. hhh4: Endemic-epidemic modeling of areal count time series. J Stat Softw 1, 1–55.

Nayava, J.L., 1980. Rainfall in nepal. Himalayan Review 12, 1–18.

Nepal National Statistics Office, N., 2021. 2021 nepal national population and housing census. URL: https://nepal.unfpa.org/en/publications/12th-national-population-and-housing-census-2021.

Ngwe Tun, M.M., Pandey, K., Nabeshima, T., Kyaw, A.K., Adhikari, M., Raini, S.K., Inoue, S., Dumre, S.P., Pandey, B.D., Morita, K., 2021. An outbreak of dengue virus serotype 2 cosmopolitan genotype in nepal, 2017. Viruses 13, 1444.

Organization, W.H., for Research, S.P., in Tropical Diseases, T., of Control of Neglected Tropical Diseases, W.H.O.D., Epidemic, W.H.O., Alert, P., 2019. Dengue: guidelines for diagnosis, treatment, prevention and control. World Health Organization.

Pandey, B.D., Costello, A., 2019. The dengue epidemic and climate change in nepal. The Lancet 394, 2150–2151.

Pandey, B.D., Nabeshima, T., Pandey, K., Rajendra, S.P., Shah, Y., Adhikari, B.R., Gupta, G., Gautam, I., Tun, M.M., Uchida, R., et al., 2013. First isolation of dengue virus from the 2010 epidemic in nepal. Tropical Medicine and Health 41, 103–111.

Pandey, B.D., Rai, S.K., Morita, K., Kurane, I., 2004. First case of dengue virus infection in nepal. Nepal Medical College Journal: NMCJ 6, 157–159.

Parpia, A.S., Skrip, L.A., Nsoesie, E.O., Ngwa, M.C., Abah, A.S.A., Galvani, A.P., Ndeffo-Mbah, M.L., 2020. Spatio-temporal dynamics of measles outbreaks in cameroon. Annals of epidemiology 42, 64–72.

Phanitchat, T., Zhao, B., Haque, U., Pientong, C., Ekalaksananan, T., Aromseree, S., Thaewnongiew, K., Fustec, B., Bangs, M.J., Alexander, N., et al., 2019. Spatial and temporal patterns of dengue incidence in northeastern thailand 2006–2016. BMC infectious diseases 19, 1–12.

Prajapati, S., Napit, R., Bastola, A., Rauniyar, R., Shrestha, S., Lamsal, M., Adhikari, A., Bhandari, P., Yadav, S.R., Manandhar, K.D., 2020. Molecular phylogeny and distribution of dengue virus serotypes circulating in nepal in 2017. PloS One 15, e0234929.

Rijal, K.R., Adhikari, B., Ghimire, B., Dhungel, B., Pyakurel, U.R., Shah, P., Bastola, A., Lekhak, B., Banjara, M.R., Pandey, B.D., et al., 2021. Epidemiology of dengue virus infections in nepal, 2006–2019. Infectious diseases of poverty 10, 1–10.

Rincón, L.M.G., 2020. Statistical Methods for Campylobacter Outbreak Detection using Genomics and Epidemiological Data. Phd thesis. University of Warwick.

Roback, P., Legler, J., 2021. Beyond multiple linear regression. Applied Generalized Linear Models and Multilevel Models in R, 436.

Rozilawati, H., Zairi, J., Adanan, C., et al., 2007. Seasonal abundance of aedes albopictus in selected urban and suburban areas in penang, malaysia. Trop Biomed 24, 83–94.

Semakula, M., Niragire, F., Nsanzimana, S., Remera, E., Faes, C., 2023. Spatio-temporal dynamic of the covid-19 epidemic and the impact of imported cases in rwanda. BMC Public Health 23, 1–13.

Sugeno, M., Kawazu, E.C., Kim, H., Banouvong, V., Pehlivan, N., Gilfillan, D., Kim, H., Kim, Y., 2023. Association between environmental factors and dengue incidence in lao people’s democratic republic: a nationwide time-series study. BMC Public Health 23, 2348.

Sun, W., Xue, L., Xie, X., 2017. Spatial-temporal distribution of dengue and climate characteristics for two clusters in sri lanka from 2012 to 2016. Scientific reports 7, 12884.

Tatem, A.J., 2017. Worldpop, open data for spatial demography. Scientific data 4, 1–4.

Tovissodé, C.F., Lokonon, B.E., Glèlè Kakaï, R., 2020. On the use of growth models to understand epidemic outbreaks with application to covid-19 data. Plos one 15, e0240578.

Utazi, C.E., Wagai, J., Pannell, O., Cutts, F.T., Rhoda, D.A., Ferrari, M.J., Dieng, B., Oteri, J., Danovaro-Holliday, M.C., Adeniran, A., et al., 2020. Geospatial variation in measles vaccine coverage through routine and campaign strategies in nigeria: Analysis of recent household surveys. Vaccine 38, 3062–3071.

Viboud, C., Simonsen, L., Chowell, G., 2016. A generalized-growth model to characterize the early ascending phase of infectious disease outbreaks. Epidemics 15, 27–37.

Wariri, O., Utazi, C.E., Okomo, U., Metcalf, C.J.E., Sogur, M., Fofana, S., Murray, K.A., Grundy, C., Kampmann, B., 2023. Mapping the timeliness of routine childhood vaccination in the gambia: A spatial modelling study. Vaccine 41, 5696–5705.

(WHO), W.H.O., 2019a. National guidelines on prevention, management and control of dengue in nepal. URL: https://www.edcd.gov.np/resource-detail/national-guidelines-of-prevention-control-and-management-of-dengue-in-nepal-2019-updated.

(WHO), W.H.O., 2019b. Ten threats to global health in 2019. URL: https://www.who.int/news-room/spotlight/ten-threats-to-global-health-in-2019.

Yang, X., Quam, M.B., Zhang, T., Sang, S., 2021. Global burden for dengue and the evolving pattern in the past 30 years. Journal of travel medicine 28, taab146.

Zeng, Z., Zhan, J., Chen, L., Chen, H., Cheng, S., 2021a. Global, regional, and national dengue burden from 1990 to 2017: A systematic analysis based on the global burden of disease study 2017. EClinicalMedicine 32.

Zeng, Z., Zhan, J., Chen, L., Chen, H., Cheng, S., 2021b. Global, regional, and national dengue burden from 1990 to 2017: A systematic analysis based on the global burden of disease study 2017. EClinicalMedicine 32, 100712.

Zwilling, M., 2013. Negative binomial regression. The Mathematica Journal 15.

